# Multi-task Bayesian Model Combining FDG-PET/CT Imaging and Clinical Data for Interpretable High-Grade Prostate Cancer Prognosis

**DOI:** 10.1101/2024.06.19.24308396

**Authors:** Maxence Larose, Louis Archambault, Nawar Touma, Raphaël Brodeur, Félix Desroches, Nicolas Raymond, Daphnée Bédard-Tremblay, Danahé LeBlanc, Fatemeh Rasekh, Hélène Hovington, Bertrand Neveu, Martin Vallières, Frédéric Pouliot

**Author notes:** Contributing authors.

## Abstract

We propose a fully automatic multi-task Bayesian model, named Bayesian Sequential Network (BSN), for predicting high-grade (Gleason **≥** 8) prostate cancer (PCa) prognosis using pre-prostatectomy FDG-PET/CT images and clinical data. BSN performs one classification task and five survival tasks: predicting lymph node invasion (LNI), biochemical recurrence-free survival (BCR-FS), metastasis-free survival, definitive androgen deprivation therapy-free survival, castration-resistant PCa-free survival, and PCa-specific survival (PCSS). Experiments are conducted using a dataset of 295 patients. BSN outperforms widely used nomograms on all tasks except PCSS, leveraging multi-task learning and imaging data. BSN also provides automated prostate segmentation, uncertainty quantification, personalized feature-based explanations, and introduces *dynamic predictions*, a novel approach that relies on short-term outcomes to refine long-term prognosis. Overall, BSN shows great promise in its ability to exploit imaging and clinico-pathological data to predict poor outcome patients that need treatment intensification with loco-regional or systemic adjuvant therapy for high-risk PCa.

## 1 Introduction

Prostate cancer (PCa) is the most frequent cancer and the second leading cause of cancer death among men in the United States [1]. Treatments for loco-regional non metastatic primary and recurrent PCa include radical prostatectomy, radiation therapy and/or hormonal therapy [2–4]. To improve patient’s oncological outcomes while limiting the morbidity of treatment overintensification, PCa treatment must be personalized based on an accurate quantitative prognostication. For this purpose, pretreatment data-based nomograms capable of predicting patient outcomes have been developed and validated [5–8], and their use is recommended by the latest National Comprehensive Cancer Network (NCCN) guidelines [9]. The most widely used and validated preoperative risk assessment tools for PCa are the Memorial Sloan Kettering Cancer Center (MSKCC) nomogram [10] and the Cancer of the Prostate Risk Assessment (CAPRA) score [11]. These nomograms rely solely on clinical data (CD) and do not incorporate any imaging information.

Prostate-Specific-Membrane-Antigen (PSMA) imaging using positron emission tomography (PET) and computed tomography (CT) is now recognized as the most accurate imaging staging modality for localized and recurrent PCa. However, the novelty of its use limits its radiomics data correlation with long-term outcomes such as metastatic recurrence or time to castration resistance. Moreover, despite showing improved accuracy compared to conventional imaging for metastasis detection, PSMA-PET/CT misses over 50% of lymph node (LN) metastases compared to lymphadenectomy histopathology at radical prostatectomy (RP) [12–14]. Recently, studies have shown that PSMA could improve nomograms’ performance to predict LN invasion, but these were not correlated with biochemical recurrence after surgery [15, 16]. Our group demonstrated that intraprostatic ^18^F-fluorodesoxyglucose (FDG) uptake on PET prior to RP is a significant prognostic marker for high-grade PCa at biopsy (Gleason ≥ 8) [17–19], independently predicting outcomes such as biochemical recurrence and time to castration-resistant prostate cancer [19].

The most common approach for incorporating PET/CT imaging data into PCa prognostic models is to reduce the PET information to a single metric such as maximum standardized uptake value (SUV) or metastatic LN status [20]. Alternatively, extraction of handcrafted radiomics (HCR), i.e., predefined quantitative attributes of an image [21], from a manually delineated intraprostatic region of CT and PET images is another standard method [22–24]. To obviate the need for manual prostate contouring and facilitate clinical translation, methods based on extraction of HCRs from an automatic segmentation map generated by a convolutional neural network (CNN) [25] have been proposed and validated [26–28]. These approaches have inspired the use of deep learning-based radiomics (DLR), i.e., latent vectors in deep layers of a CNN [29], for PCa prognosis [30]. Furthermore, a systematic review has recently suggested that combining clinical and imaging data in a single model could significantly enhance its performance [31]. For instance, a study recently showed that integrating CT-based HCR to CD leads to improved predictions of progression-free survival in high-grade PCa [32]. Nevertheless, there is no consensus on which prognostic tasks benefit from integrating FDG-PET/CT-based HCR or DLR with CD in comparison to using CD alone.

The current study aims to develop a fully automatic prognostic multi-task [33] model that takes as input both clinical and FDG-PET/CT imaging data without the need for manual segmentation (see Fig. 1a). The cohort consists of 295 patients diagnosed with high-grade PCa at biopsy who underwent RP (see Methods, section *Cohort description*). The global median follow-up time is 70 months (see Supplementary Table 8). We define six prognostic tasks associated with clinical outcomes: a binary classification task, which is the prediction of the probability of lymph node invasion (LNI); and five survival tasks, which include the prediction of biochemical recurrence-free survival (BCR-FS), metastasis-free survival (MFS), definitive androgen deprivation therapy-free survival (dADT-FS), castration-resistant prostate cancer-free survival (CRPC-FS), and prostate cancer-specific survival (PCSS). Occurrence of these events follows the natural history of high-grade PCa (see Fig. 1b). The distributions of survival time (see Fig. 1c) in the *learning set* (see Fig. 8a for a description of subsets) align with PCa progression (see Fig. 1b). This progression defines a natural sequence of event occurrences, providing a clinically coherent way of constructing a multi-task model. We propose the Sequential Network (SN) (see Fig. 2), a multi-task model comprised of several single-task models, which allows for *dynamic predictions*, i.e., predictions that are refined over time as outcomes from previous tasks unfold. For each single-task model within SN, we investigate the benefits of integrating HCR or DLR with CD using a series of experiments. We validate our approach by comparing SN’s performance with the MSKCC nomogram, CAPRA score, and single-task models.

**Fig. 1.**
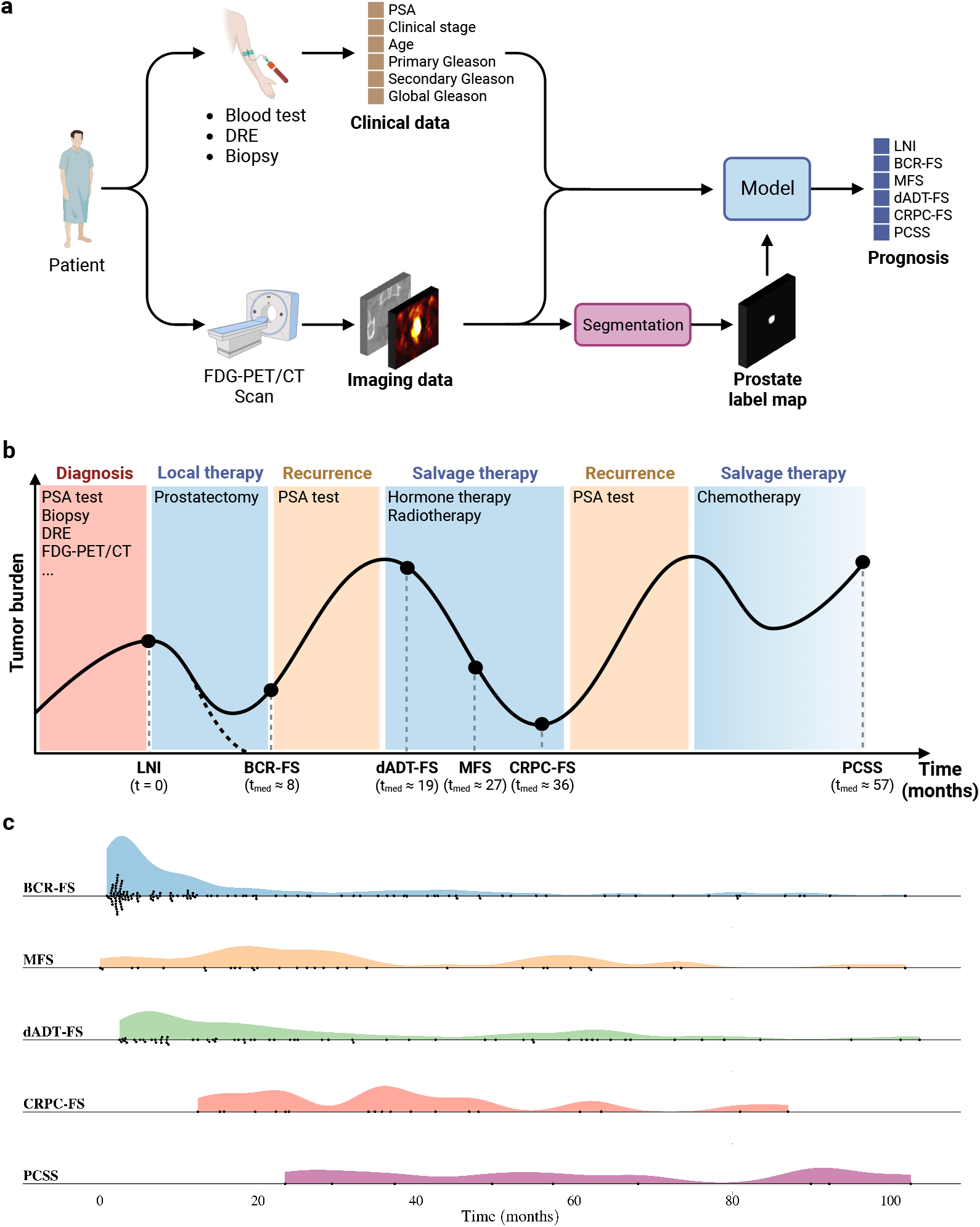
Study overview. (a) Schematic outline of a patient’s journey from diagnosis to prognosis. Following an FDG-PET/CT scan, a physician manually delineates the prostate on the CT image to create a segmentation map. This map is used to train the model and is not needed to infer the prognosis of a new patient, as the trained model automatically performs segmentation. Indeed, the study aims to develop a fully automatic prognostic model that takes as input both clinical and imaging data without requiring any manual steps. (b) Schematic representation of the natural history of high-grade PCa [44, 45]. All patients in the cohort underwent radical prostatectomy (RP), and therefore, event time is measured from the date of RP. Note that *t*_med_ is the median survival time, calculated based on data from the 250 patients in the *learning set*. See Supplementary Table 7 & 8 for in-depth analyses of survival and follow-up time distributions, respectively. (c) Distributions of survival time in the *learning set*. Each marker corresponds to a patient who suffered the corresponding event. Distributions are consistent with the natural progression of PCa. See Supplementary Fig. 2a, 5a, 8a, 11a & 14a for the Kaplan-Meier curves of each survival task on the *learning set* and Supplementary Fig. 18 for distributions of event indicators.

**Fig. 2.**
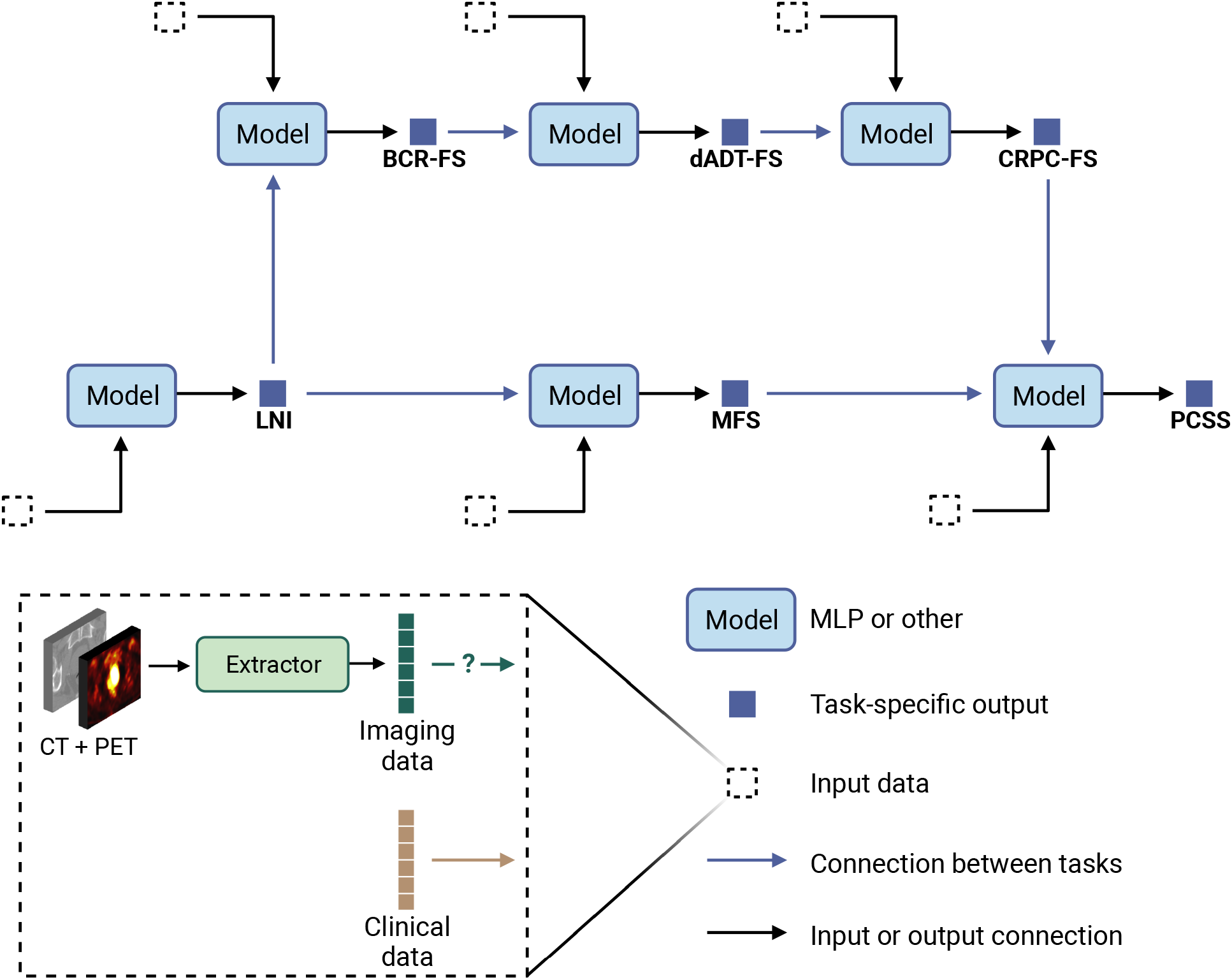
Framework of the Sequential Network (SN). SN is a multi-task model comprised of several single-task models. Single-task models can be any feed-forward neural network, such as a multi-layer perceptron (MLP). The output of each single-task model represents either positive class probability for classification tasks or event risk for survival tasks. Each model has 3 different input types: clinical data (mandatory), imaging data (task-specific), and predictions from previous models (task-specific), hence the terminology “sequential”. The sequence of tasks was determined based on the natural history of prostate cancer (see Fig. 1b). See Supplementary Fig. 1 for the correlation between each pair of tasks.

To enhance the model’s reliability and trustworthiness in a clinical context, it is important to provide clinicians with information about the systematic uncertainty of the model regarding an individual patient’s prediction [34–36]. To quantify the uncertainty, we created a Bayesian version of the SN using variational inference (VI) [37, 38]. We opted for VI over the commonly used Monte-Carlo Drop-Out (MCDO) in uncertainty-aware neural networks, as MCDO suffers from limitations such as unreliable uncertainty quantification, poor calibration, and weak out-of-distribution detection [39–41]. We validate our approach by comparing the performance of the Bayesian Sequential Network (BSN) with the deterministic SN. We also provide a feature-based explanation of model prediction using SHAP [42] and SurvSHAP(*t*) [43] frameworks.

In summary, the proposed model is a fully automatic BSN that combines FDG-PET/CT imaging data with CD to predict LNI, BCR-FS, MFS, dADT-FS, CRPC-FS, and PCSS in high-grade PCa. In the context of PCa prognosis and to the best of our knowledge, we are the first to systematically determine the contribution of FDG-PET/CT-based HCR and DLR on task-specific model performance, to use multi-task learning in a sequential approach, and to provide model’s uncertainty using VI.

## 2 Results

The BSN is developed using a constructive approach; each hypothesis is tested through a comparison experiment, with results of previous experiments guiding subsequent ones. Experiments are conducted using a *learning set* of 250 patients, with evaluation carried out (by 5-fold cross-validation) on 5 *test sets* of 50 patients each (see Methods, section *Experimental setup*). The *holdout set* is used exclusively for assessing the performance of the best model, selected among all models based on its average performance on the *test sets*. The workflow of the conducted experiments is outlined below, with hypotheses in italics.

1. **Single-task model (MLP vs. LR)**. *To assess whether a multi-layer perceptron* (*MLP*) *outperforms a linear regression* (*LR*) *and thereby justify transitioning to deep learning approaches*, the performance of MLPs and LRs, trained exclusively with CD, is compared.
2. **HCRs extraction (Automatic vs. manual segmentation)**. *To verify that models using HCRs extracted from an automatically segmented prostate region perform equally well compared to those using contours from a physician*, the performance of MLPs trained with HCRs obtained through either manual or automatic prostate segmentation is compared.
3. **DLRs extraction (U-NEXtractor vs. CNN)**. *To verify that simultaneously performing prostate segmentation and prognosis leads to the extraction of higher-quality image features*, the performance of models trained solely with images, specifically those that extract DLRs like U-NEXtractor and CNN, are compared.
4. **Imaging data integration (CD vs. CD+HCR vs. CD+DLR)**. *To assess whether imaging data improve the prognosis of some outcomes*, the performance of MLPs trained with both CD and either HCR or DLR is compared with MLPs trained solely with CD.
5. **Multi-task model (SN vs. MLP)**. *To assess whether multi-task learning enhances the performance for some tasks*, SN is evaluated and compared to MLPs.
6. **Uncertainty quantification (BSN vs. SN)**. *To validate that probabilistic models perform just as well as their deterministic counterparts, while also providing an uncertainty quantification of individual predictions*, the performance of SN is compared to its Bayesian counterpart, BSN.
7. **Dynamic predictions (BSN**_*t*=0_ **vs. BSN**_*t*>0_). *To assess whether dynamic predictions improve the prognosis of long-term outcomes*, accuracy of dynamic and static predictions are compared.
8. **Baseline models (MSKCC & CAPRA vs. BSN)**. *To determine whether the developed model outperforms baseline models*, the MSKCC nomogram and CAPRA score are evaluated and compared to BSN. Note that the MSKCC nomogram is trained with over 10,000 patients, so its impressive performance may stem from the sheer number of training cases rather than from its underlying model, i.e. a LR that exclusively uses CD.

In the following sections, the results of the aforementioned experiments are presented, culminating in the selection of the best-performing model. This model is then evaluated on the *holdout set*, with visuals depicting its performance on both *test sets* and *holdout set*. Furthermore, a global interpretation of predictions is provided, along with a patient prognosis example to illustrate how the model can support clinical decisions.

### 2.1 Model comparison and selection

#### Performance metrics

The performance metrics used for the classification task are the area under the receiver operating characteristic (ROC) curve (AUC) and binary balanced accuracy (BA); survival tasks rely on concordance index (CI), CI based on inverse probability of censoring weights (CIWC), and cumulative/dynamic AUC (CDA); whereas segmentation tasks are evaluated solely based on Dice similarity coefficient (DSC) (see Methods, section *Tasks implementation*).

#### Single-task model

For all experiments, the selected single-task model used to predict each clinical outcome is an MLP (see Methods, section *Models*), as MLPs show a substantial and consistent increase in performance compared to LRs for all tasks (see Supplementary Table 24, section A).

#### HCRs extraction

The U-Net (see Fig. 9a), trained with manual contours, generates a prostate segmentation map on the CT. The segmentation map is then used to extract HCRs on both CT and PET images in the intraprostatic region (see Fig. 3a for HCR extraction pipeline). Opting for automatic segmentation maps instead of manual ones does lead to a minor performance decrease (see Supplementary Table 24, sections B & C), but we argue that the advantages of automation in a clinical setting outweigh this drawback. Of 200 HCRs initially extracted (see Methods, section *Handcrafted radiomic features*), the 6 selected HCRs with the highest Gini importance [46], obtained from a random forest classifier trained to predict LNI, are all first-order intensity-based features extracted from PET images (see Supplementary Fig. 19). Note that the number of selected HCRs (6 features) is arbitrarily chosen to be equal to the number of CD. The automatically selected HCRs are the same with U-Net and Bayesian U-Net, even though U-Net exhibited slightly higher average DSCs, i.e., 0.845 compared to 0.834 on *test sets* (see Supplementary Table 25).

**Fig. 3.**
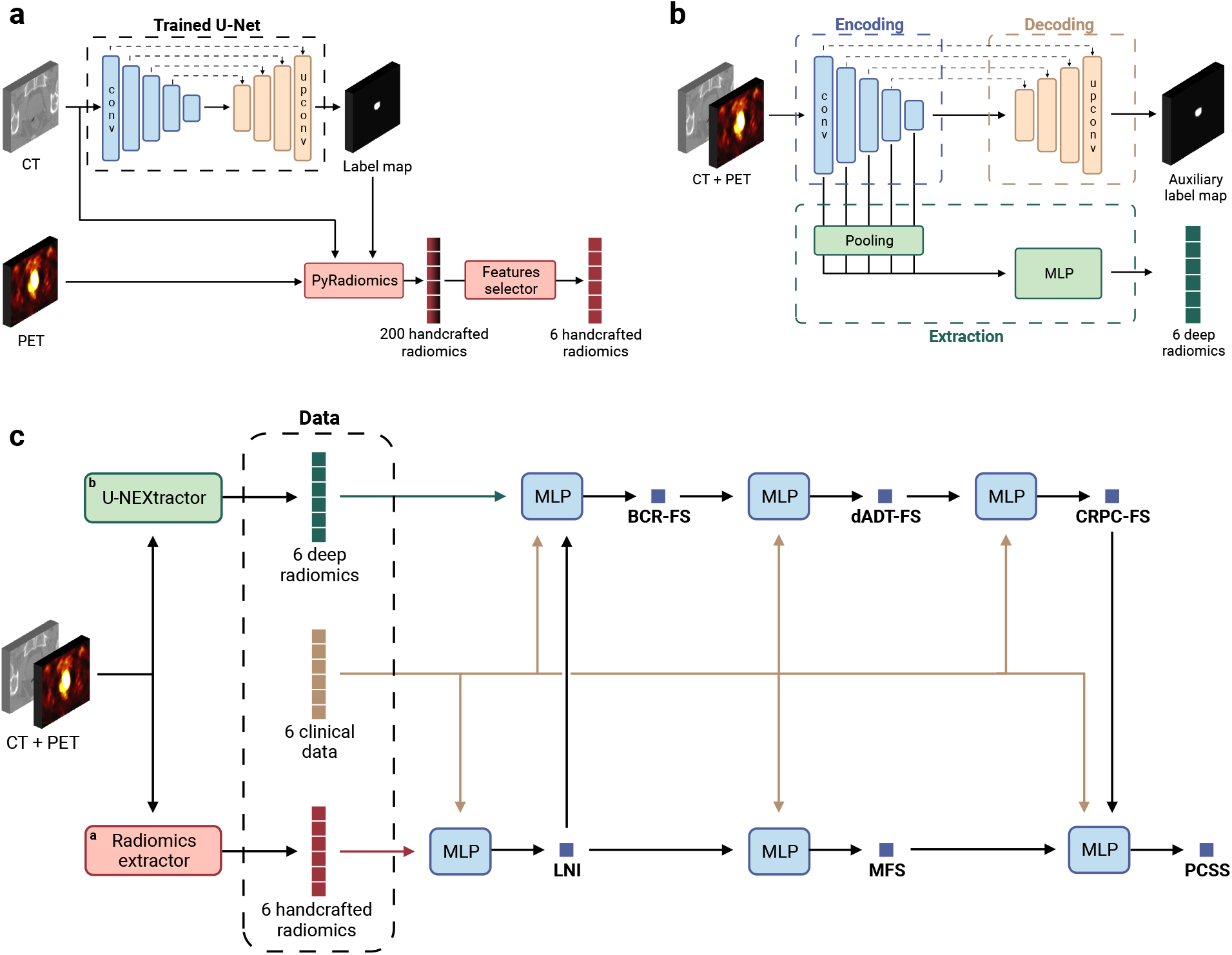
Sequential Network (SN). (a) Handcrafted radiomic features extraction pipeline. The U-Net, trained with manual contours, automatically generates a prostate segmentation map from the CT image (see Fig. 9a for the visualization of feature maps in different layers of the U-Net). A total of 200 radiomic features are computed with the `pyradiomics` [47] Python library (see Supplementary Table 21 & 22 for extraction parameters on CT and PET images, respectively), using voxels from both CT and PET images within the segmented region. The Gini importance [46] of each feature is then determined using a random forest classifier with 10,000 trees implemented with the `scikit-learn` [48] Python library and trained to predict a single task using 200 extracted radiomics. The 6 features with highest Gini importance are selected (see Supplementary Fig. 19 for selected features). (b) Deep radiomic features extraction pipeline. The model, named U-NEXtractor, segments the prostate and extracts deep radiomic features simultaneously. The idea is that the auxiliary segmentation task is expected to spatially guide the network to extract prognostically relevant features in the prostate region [49]. See Fig. 9b for the U-NEXtractor’s detailed architecture. (c) Architecture of the final SN (see Fig. 2 for the conceptual framework). The input data for each single-task model is based on the data that yields the highest scores for MLP on the *test sets*. This refers to the best data (see Table 1, section B), i.e., clinical data and handcrafted radiomics for LNI, clinical data and deep radiomics for BCR-FS, and clinical data for MFS, dADT-FS, CRPC-FS, and PCSS.

#### DLRs extraction

The U-NEXtractor (see Fig. 9b) simultaneously performs the segmentation of the prostate and the extraction of DLRs (see Fig. 3b for DLR extraction pipeline). This approach aims to use segmentation as an auxiliary task to guide the network in capturing clinically significant features in the prostate region, without however being limited to the ground truth region [49]. This method demonstrates comparable prognosis performances to a conventional CNN, i.e., U-NEXtractor without the decoding branch (sand-colored branch in Fig. 9b), but it achieves highest overall performance scores on survival tasks (see Supplementary Table 24, section D). U-NEXtractor and Bayesian U-NEXtractor produce poor prostate segmentation maps, with average DSCs of 0.26 and 0.05. The DSC itself is not of particular interest, as the auxiliary segmentation task is only used to enhance predictive performance. Nonetheless, the fact that the segmentation map is not random suggests that the network is prioritizing a specific region to enhance performance. For instance, the segmentation map overlaid on the PET image reveals that Bayesian U-NEXtractor avoids bones and regions of high FDG uptake by the bladder, and segments everything else (see Supplementary Fig. 22).

**Table 1.**
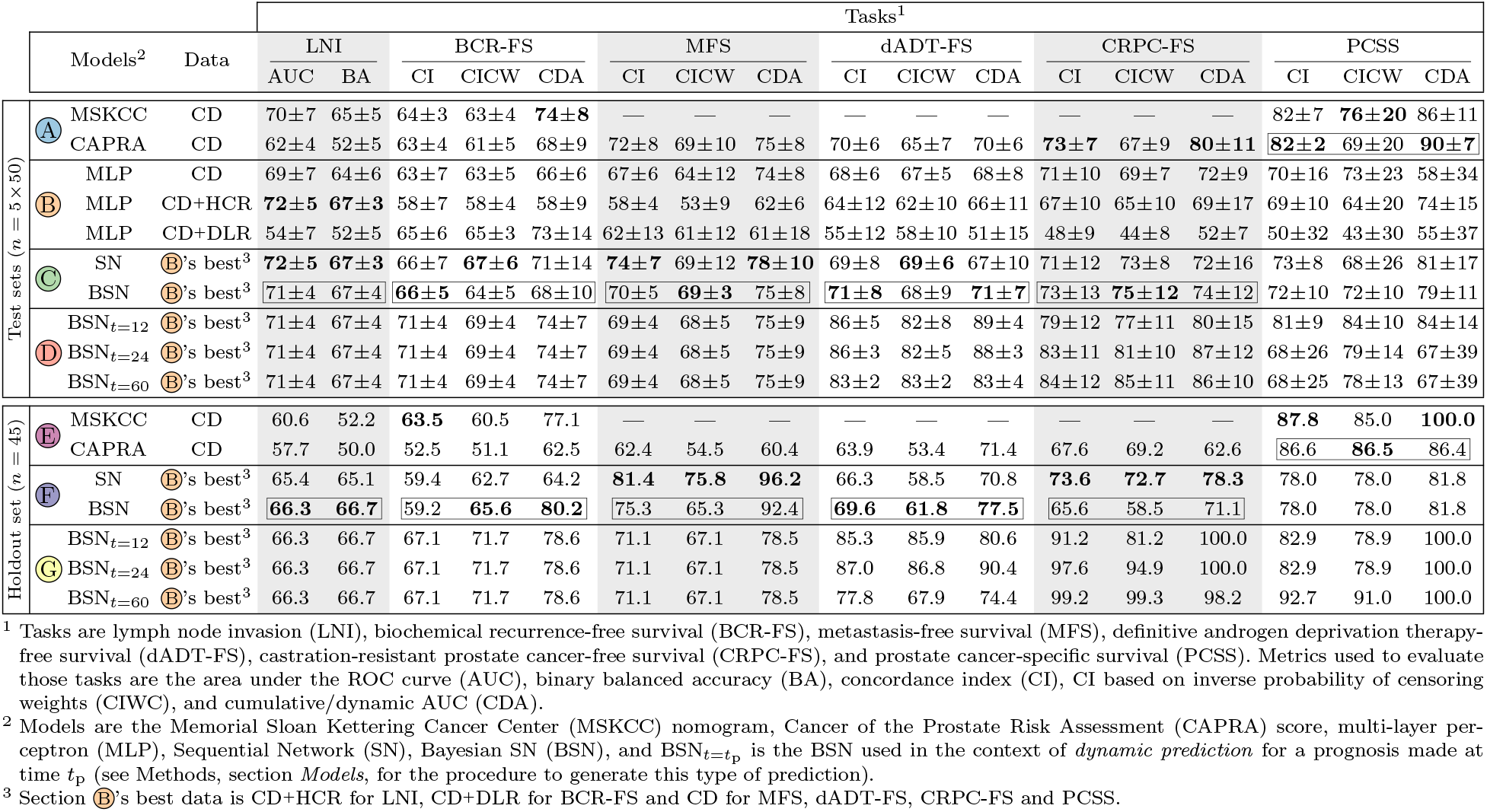
Performance comparison of the models. Performance on the *test sets* and the *holdout set* of (A, E) baseline nomograms, (B) single-task models, (C, F) multi-task sequential models, and (D, G) dynamic multi-task sequential models. Possible input data to models are clinical data (CD), clinical data and handcrafted radiomics (CD+HCR), and clinical data and deep radiomics (CD+DLR). Scores on *test sets* correspond to the *mean ± standard deviation* of the scores on the 5 *test sets* of 50 patients each. The highest score achieved by the models in sections A, B, and C (*test sets*), and in sections E and F (*holdout set*) are highlighted in bold (see Supplementary Table 23 for additional metric scores on the LNI task and Supplementary Table 26 & 27 for calculated *p*-values). Note that dynamic models in sections D and G are excluded from the process of determining the highest score to ensure fairness, as they have the advantage of having access to target data as model inputs. The selected models, chosen for each task through experiments conducted on *test sets* (see Fig. 8), have their metric scores enclosed in a box. The prostate segmentation maps generated by U-Net and Bayesian U-Net, which are used to compute handcrafted radiomic features, respectively achieved average DSC of 0.842±0.004 and 0.821±0.004 on the *test sets* compared to manual contours delineated by a physician. Likewise, these models respectively achieved average DSC of 0.845 and 0.834 on the *holdout set* of 45 patients. See Supplementary Table 25 for additional information on DSC.

#### Imaging data integration

The *best data*, i.e., data that yields the highest score on the *test sets*, corresponds to CD+HCR for LNI, CD+DLR for BCR-FS, and CD for MFS, dADT-FS, CRPC-FS and PCSS (see Table 1, section B, for scores, and Supplementary Table 27 for *p*-values). These results show that only LNI and BCR-FS benefit from the integration of imaging data.

#### Multi-task model

The selected input data of each MLP comprised in SN (see Fig. 3c for SN architecture) is based on the *best data* of each task. SN outperforms (for BCR-FS, MFS, dADT-FS, CRPC-FS, and PCSS) or matches (for LNI) MLPs, though not significantly (see Table 1, sections B & C, and Supplementary Table 27).

#### Uncertainty quantification

Performance comparison between BSN and SN shows no significant difference, except in the case of CDA for BCR-FS, where SN significantly outperforms BSN (see Table 1, section C, and Supplementary Table 26).

#### Dynamic predictions

*Dynamic predictions* are predictions that are refined over time according to the clinical results of previous tasks (see Methods, section *Models*, for a description of *dynamic predictions*). In a clinical setting, *dynamic predictions* are expected to be employed following the occurrence of an event to acquire fresh and improved predictions for subsequent events. These predictions show interesting potential (see Table 1, section D & G). Indeed, BSN’s performance at *t* = {12, 24, 60} months is considerably higher than at diagnosis time. Note that these time points were chosen to align with standard follow-up intervals.

#### Baseline models

Statistical analysis (see Table 1, sections A & C, and Supplementary Table 26) of scores obtained on *test sets* shows that the performance of BSN is significantly higher than that of CAPRA for LNI and dADT-FS. For PCSS, the performance of MSKCC and CAPRA is significantly higher than that of BSN.

#### Model selection

The selected models (one choice of model per task, hence the plural) are BSN for all tasks except PCSS, which uses CAPRA score (see Table 1, sections A, B & C, boxed model’s scores). Selection is based on the model’s performance on *test sets* and on the fact that Bayesian models offer the advantage of uncertainty quantification.

### 2.2 Performance of the selected model

#### Performance on *holdout set*

On the *holdout set*, selected models show higher performance than others for LNI, BCR-FS, and dADT-FS, but lower performance for MFS, CRPC-FS, and PCSS (see Table 1, sections E & F; selected models are boxed). Statistical analysis (see Supplementary Table 26) shows no significant difference, except in the case of CDA for MFS, where BSN significantly outperforms CAPRA, and for PCSS, where, as in the *test sets*, the performance of baseline models significantly outperforms ours. *Dynamic predictions* (see Table 1, section G) show considerable improvement for most long-term tasks compared to prognosis made at diagnosis time.

#### Risk stratification performance

Stratification performance of selected models is evaluated on the *test sets* and *holdout set* (see Fig. 4). Log-rank tests [50, 51] performed on Kaplan-Meier curves show that high- and low-risk groups, established based on inferred risks (see Methods, section *Risk groups*, for a description of risk thresholds), separate well (*p* < 0.05) on the *test sets* for all survival tasks, except CRPC-FS where groups tend towards (but did not attain) significantly different separation with *p* = 0.09 (see Fig. 4). On the *holdout set*, only CRPC-FS and PCSS show a significant group difference, but BCR-FS and MFS also tend towards a significant group separation with *p* = 0.13 and *p* = 0.09, respectively. For dADT-FS, the two risk groups are confounded (*p* = 0.6).

**Fig. 4.**
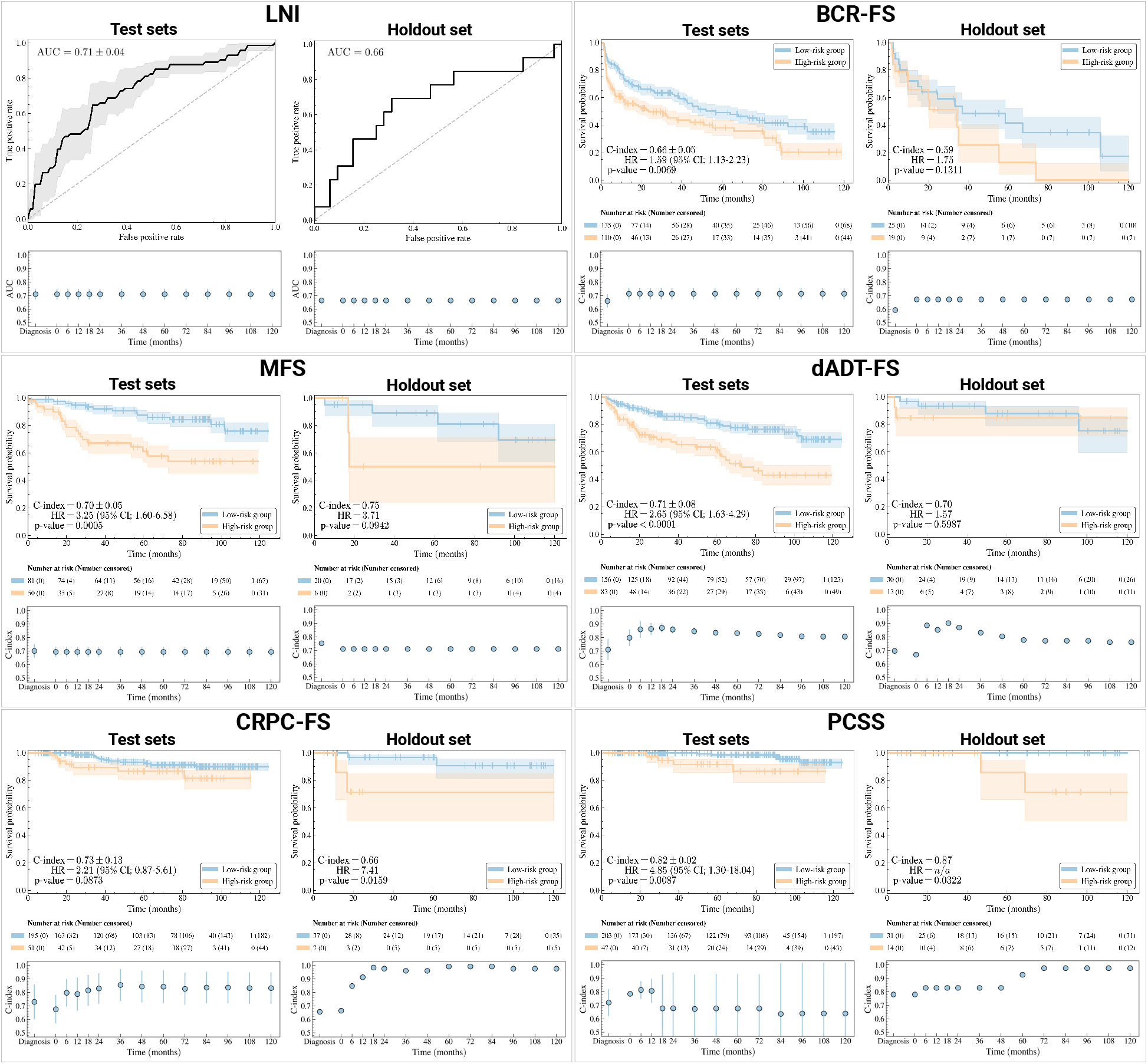
Visualization of the performance of selected models. Selected models are Bayesian Sequential Network (BSN) for all tasks except PCSS, which uses CAPRA score. For the classification task (i.e. LNI), receiver operating characteristic (ROC) curves [52] on the *test sets* (top left) and *holdout set* (top right) are shown. The ROC curve on *test sets* corresponds to the mean (line) and standard deviation (shade) of the ROC curves on the 5 *test sets*. For each survival task, the model’s ability to stratify patients into clinically significant risk groups is illustrated by Kaplan-Meier curves [53] of *test sets* (top left) and *holdout set* (top right) using stratification based on predicted risk (see Methods, section *Risk groups*, for a description of the risk threshold computation method). The 95% confidence interval (95% CI, shade) of the Kaplan-Meier curve (line) is estimated using log hazard [54]. The *p*-value is computed using a log-rank test [50, 51], which also provides statistics to calculate the hazard ratio (HR) and its 95% confidence interval [55]. For each task, the performance of BSN’s *dynamic predictions* is shown both on *test sets* (bottom left) and *holdout set* (bottom right). Results show that *dynamic predictions* are refined over time as events unfold. Scores on the *test sets* correspond to the mean (marker) and standard deviation (error bar) of scores on the 5 *test sets*.

#### Single-feature-based risk stratification

The clinical features that yield the best single-feature-based risk stratification on the *test sets*, i.e., obtained the lowest *p*-value, are the global Gleason score for BCR-FS and MFS, and secondary Gleason score for other tasks (see Supplementary Fig. 3c, 6e, 9c, 12e & 15e). All of these *p*-values are lower than those obtained with risk groups inferred by models shown in 4, showing that selected models improve risk stratification of high-grade PCa compared to a single-feature-based approach. However, on the *holdout set*, the global Gleason score stratifies better than selected models for BCR-FS, MFS, and dADT-FS (see Fig. 4 & Supplementary Fig. 4c, 7c, 10c).

#### Time-dependence of *dynamic predictions*

*Dynamic predictions*’ performance for LNI remains constant over time (see Fig. 4, bottom figures), which is expected since this is the first task in the sequence defined by SN (see Fig. 3a), and therefore no additional information is provided as input to LNI’s single-task model following diagnosis. Similarly, performance remains constant for BCR-FS and MFS tasks once LNI is assessed at RP, i.e. at *t* = 0; however, BCR-FS shows a slight increase in performance, while MFS experiences a minor decrease. For subsequent tasks, namely dADT-FS, CRPC-FS, and PCSS, *dynamic predictions* substantially improve performance compared to predictions made at diagnosis time on both *test sets* and *holdout set*, except for PCSS on *test sets*, which exhibits a decrease after 12 months. However, performance variations for PCSS are quite large, given the low number of events (*n* = 9) in the *test sets*. Finally, both dADT-FS and CRPC-FS tend to reach a plateau or even decline in performance after a certain duration.

### 2.3 Global interpretability analysis

#### Importance of HCRs

Explanations for predictions of selected models are provided for the 45 patients in the *holdout set* (see Fig. 5). SHapley Additive exPlanations (SHAP) [56] values (see Fig. 5a) show that the importance of the mean PET image intensity in the intraprostatic region is the feature with the most influence when discriminating patients with high and low probability of LNI, thus supporting the selection of HCRs as additional input data for this task. Furthermore, four of six HCRs are among the nine most influential features for this task.

**Fig. 5.**
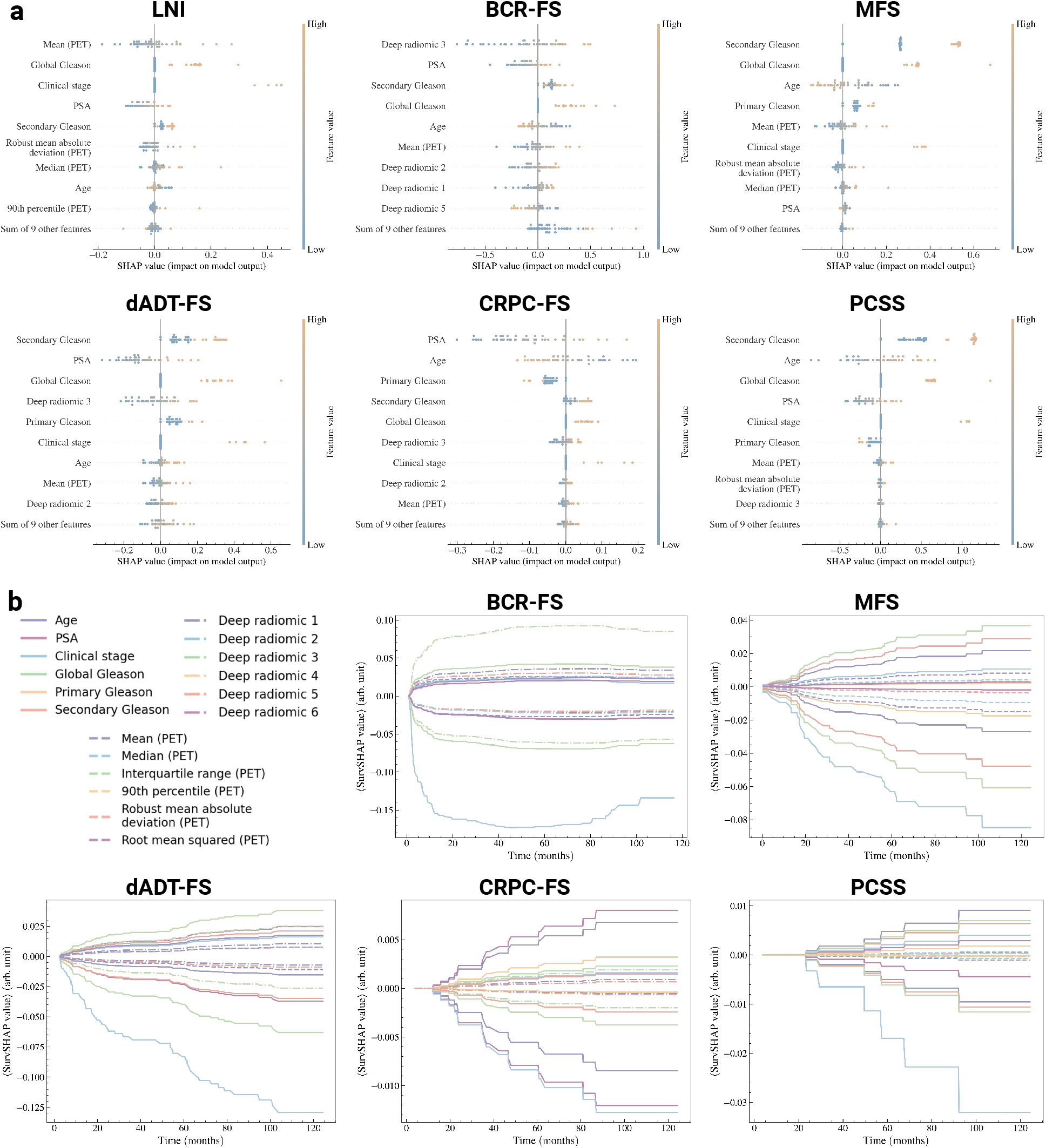
Explanation of the predictions of selected models. Selected models correspond to the boxed models in Table 1, sections A, B & C, and predictions are made on the 45 patients in the *holdout set*. (a) SHapley Additive exPlanation (SHAP) [56] values of every patient in the *holdout set* for each task. Features are ranked in order of mean absolute importance, with the most important at the top. For each feature, a marker is associated to each patient. (b) Time-dependent SHAP (SurvSHAP(*t*)) [57] curves of the top 9 most important features shown in *panel A* for each survival task. For each figure, 2 curves are shown per feature: one is the mean of strictly positive SurvSHAP(*t*) curves, and the other is the mean of strictly negative SurvSHAP(*t*) curves, both obtained from each patient in the *holdout set*.

#### Importance of DLRs

A similar observation holds for BCR-FS with DLRs, as the feature *deep radiomics 3* emerges as the most discriminating feature. Once again, four of six DLRs are in the top nine most important features. For other tasks, the model’s predictions are mainly explained by secondary Gleason score, except for CRPC-FS, where prostate-specific antigen (PSA) dominates.

#### Feature importance propagation

Due to the SN’s architecture, some imaging features appear as the most important features for tasks that rely solely on CD as inputs. For example, *mean* (*PET*) ranks fifth in importance for predicting MFS, despite MFS’s single-task model (within SN) not directly using any HCRs as an input. This finding aligns with SN’s sequential architecture; as MFS’s model uses both CD and LNI prediction as inputs, and LNI’s model takes HCRs as input, then HCRs’ influence propagates through the network. In the explanatory figure of MFS (see Fig. 5a), the prediction of LNI therefore emerges as a linear combination of the features that contributed to its prediction.

#### Time-dependence of feature importance

Time-dependent SHAP (SurvSHAP(*t*)) [57] curves (see Fig. 5b) show that clinical stage is of considerable importance in predicting low-risk patients for all tasks. For BCR-FS, we notice a decrease in the importance of all features for times *t* > 60 months due to the low number of events occurring with such durations (see Fig. 1c for distribution of survival time).

### 2.4 Illustration of clinical application

The prognosis of an arbitrarily selected patient from the *holdout set* is provided (see Fig. 6 to illustrate the typical journey of a patient (see Fig. 1a), from diagnosis to prognosis, using the selected model as a clinical decision support tool. Initially, the patient’s 6 CD (see Fig. 6a) are collected through tests in regular clinical practice, followed by a FDG-PET/CT scan. The Bayesian U-Net generates a prostate segmentation map from the CT and computes uncertainty (standard deviation) (see Fig. 6b). Using this map, 6 HCRs are extracted from both CT and PET images (see Fig. 3a). In parallel, the U-NEXtractor directly uses FDG-PET/CT images to extract 6 DLRs (see Fig. 3b). All 18 features serve as input data for SN (see Fig. 3c) to predict PCa progression (see Fig. 6c), providing prognosis. For LNI, the prediction corresponds to the probability of a positive outcome and its uncertainty is a standard deviation, while for survival tasks, the prediction is provided as a survival curve and its uncertainty as a confidence interval (see Fig. 6d). For each task, the patient is classified into either high- or low-risk group (see Methods, section *Risk groups*). SHAP and SurvSHAP(*t*) values offer feature-based explanation of the prognosis (see Fig. 6e & 6f).

**Fig. 6.**
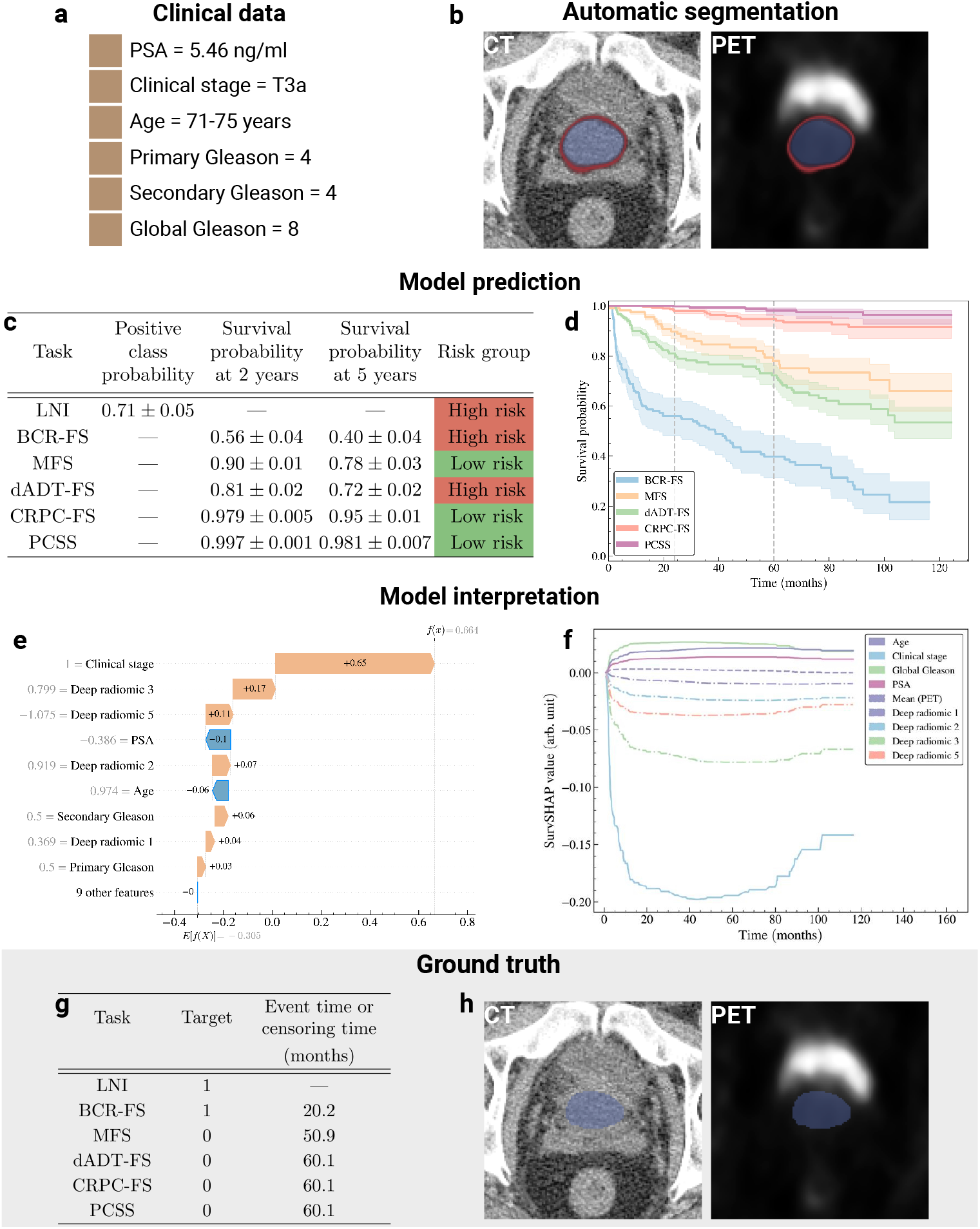
Illustration of a clinical application of selected models. The application involves establishing the prognosis of an arbitrarily selected patient from the *holdout set*. See Supplementary Fig. 20 & 21 for other examples of patient prognosis. (a) Patient’s clinical data. (b) Segmentation map of the prostate obtained from Bayesian U-Net trained on the *learning set*. The segmentation map is overlaid on CT and PET images to illustrate that the region of high FDG uptake by the bladder lies outside the segmentation map’s boundaries. The segmentation map is used to extract handcrafted radiomic features. Note that the DSC between automatic and manual segmentation (ground truth) is 0.910. Color code: average prostate segmentation map (blue) and standard deviation (red) over 100 inferences. See Supplementary Fig. 22a for the segmentation map obtained by Bayesian U-NEXtractor. (c) Average prediction and standard deviation of the model over 100 inferences. (d) Average survival curves predicted by the model (line) and 95% confidence interval (shade) over 100 inferences (e) Shapley additive explanation (SHAP) [56] of the predicted risk of BCR-FS. (f) Time-dependent SHAP (SurvSHAP(*t*)) [57] of the predicted risk of BCR-FS. (g) Ground truth progression of the patient’s cancer. Time represents the survival time when the target value is 1 and acts as a censoring time otherwise. (h) Ground truth prostate segmentation map obtained from manual contouring by a physician.

## 3 Discussion

High-grade PCa treatment must be personalized based on an accurate quantitative prognosis. Current models lack integration of imaging data, and cannot predict multiple outcomes simultaneously. Hence, a fully automatic multi-task model that uses both clinical and FDG-PET/CT imaging data is developed. The model is selected and optimized based on experiments conducted on the *learning set*, and the best-performing model (BSN for all tasks except PCSS, where CAPRA is used) is evaluated on the *holdout set*.

### Model comparison and selection

The integration of FDG-PET/CT as input data improves performance for LNI and BCR-FS (see Table 1, section B). Of the six studied outcomes, these two are the closest in time to diagnosis (see Fig. 1b & 1c). This observation highlights the temporal limitation of prognostic information derived from FDG-PET/CT images; the prognostic relevance of images decreases with the time elapsed between the event and the image acquisition, due to PCa progression. Also, the limited number of late events in the dataset (see Fig. 1c) likely contributes to suboptimal predictions for late outcomes. Furthermore, multi-task models exhibit superior performance compared to single-task models (see Table 1, sections B & C), though not significantly (see Supplementary Table 27). This improvement demonstrates the utility of sequential networks in propagating inter-task information, supporting the relevance of multi-task learning [33] in PCa prognosis. Moreover, SN and BSN exhibit comparable performance (see Supplementary Table 26), suggesting that VI exerts negligible influence on performance while enabling uncertainty quantification.

### Performance of selected model

The selected model effectively stratified patients in the *test sets* but faced challenges with the *holdout set* (see Fig.4), potentially due to over-fitting of the model’s parameters or of the risk stratification threshold. However, the limited size of the *holdout set* (*n* = 45) may not provide enough statistical power for decisive assessment of the measured statistical difference via log-rank test, as evidenced by overlapping confidence intervals of Kaplan-Meier curves of different risk groups. Nevertheless, maintaining a *holdout set* set for final model evaluation is crucial, especially considering all design choices were made on the *learning set*. Moreover, a substantial improvement of the model’s performance through *dynamic predictions* is observed (see Fig. 4), which confirms the relevance of the model’s sequential architecture, as *dynamic predictions* are only feasible due to this sequential framework. The observed performance plateau in both dADT-FS and CRPC-FS may result from the increasing unpredictability of the relationships between tasks over time. Since the model maintains the same assumption about these relationships across all time frames, it eventually leads to incorrect predictions when the assumption becomes unreliable.

### Global interpretability analysis

The FDG-PET signal is the most important variable in predicting LNI (see Fig. 5), with higher average intensity indicating a greater likelihood of LNI. In line with previous research, high intraprostatic uptake of FDG on PET-CT images correlates with higher Gleason grades [58]. The accuracy of the model with an AUC of 0.71 for LNI prediction is slightly lower than nomograms combining PSMA-PET/CT LN nodes status with clinical data [15, 16]. However, our cohort includes only high-grade patients at biopsy which decreases greatly the discriminative potential of our models because higher grade at biopsy has been shown to be a major factor discriminating pN0 from pN1 in clinical nomograms [16, 59, 60]. For BCR-FS, DLRs are highly valuable predictors, as *deep radiomics 3* is the most significant feature and persists over time (see Fig. 5b). Although not the most important, radiomic features play a significant role in predicting other outcomes, made possible by SN’s architecture propagating feature importance across tasks. As for clinical features, Gleason grades, clinical stage, and PSA are crucial variables for very high-risk PCa [61], aligning with the importance obtained for these features across all outcomes (see Fig. 5). It has been found that intrinsic tumor biology, as determined by Gleason grade, is the most significant predictor of very high-risk disease for metastases [61]. Our results confirm these findings and show that Gleason 9 and 10 predict poorer outcomes, just as advanced clinical stage (T3a) does (see Fig. 5). The predicted risk is also found to be proportional to PSA level (see Fig. 5a). Furthermore, a younger age at diagnosis may reflect a more aggressive disease, and therefore higher risk prediction (see Fig. 5a). Indeed, there is a high likelihood that PCa in younger individuals will have a genetic component, and disruption of key genes can also lead to more aggressive behavior [62, 63]. In the same way, younger age at diagnosis may be associated with cancers that exhibit hereditary genetic characteristics that make them more likely to progress and metastasize. Finally, the observed higher risk for developing CRPC at a younger age may be due to lead time bias, with individuals surviving long enough on dADT to develop CRPC.

### Illustration of clinical application

The illustration of a patient’s prognosis (see Fig. 6) allows to compare the practical benefits of the proposed approach to existing solutions. The proposed AI system, i.e., the model and accompanying tools, provides clinicians with additional benefits: automatic prostate segmentation, uncertainty estimation for individual predictions, personalized feature-based explanations, and *dynamic predictions*. The generated segmentation map has no prognostic value on its own; nonetheless, it validates the model’s focus on relevant anatomical regions. Conversely, the capability to express uncertainty by providing the standard deviation of a prediction is crucial for the safe clinical deployment of a prognosis-oriented AI system [36]. Indeed, quantifying uncertainty in individual predictions fosters trust with healthcare workers by enabling principled decision-making and serves as a protective measure against unsafe prediction failures [34]. Also, feature-based explanations could aid clinicians in understanding and validating predictions, informing decision-making. Lastly, during follow-up, clinicians could employ *dynamic predictions* to refine prognosis of long-term outcomes following occurrences of short-term outcomes. For instance, the model could be employed at diagnosis and, upon observation of BCR, reran to generate more accurate predictions for ensuing outcomes, namely dADT-FS, CRPC-FS, and PCSS, outperforming those made without prior knowledge at diagnosis.

### Limitations and future work

A limitation of the proposed approach lies in the imputation method for missing CD; since the imputation is not bayesian (see Methods, section *Preprocessing*), the estimated uncertainty fails to account for potential uncertainty stemming from the imputation process. However, a recent study has introduced ways to incorporate imputation uncertainty into a Bayesian model using a deep latent generative model [64], an avenue we aim to explore. Additionally, we intend to investigate activation maps as a tool to uncover the regions and patterns used by U-NEXtractor, which could elucidate the nature of extracted DLRs [25]. Furthermore, we aim to integrate example-based interpretability methods. Rather than explaining the model through input feature contributions, these methods interpret the model’s behavior through influential training data points, i.e., patients from the *learning set* that were important for the model prediction [65]. Finally, while FDG imaging has proven valuable in metastatic PCa, its role in high-risk prostate cancer requires refinement. Hence, validating our approach with PSMA-PET/CT patients could facilitate its translation into clinical practice.

### Conclusion

Current high-grade PCa prognostic models rely solely on CD, lacking integration of imaging data. The proposed model, BSN, outperforms the MSKCC nomogram and CAPRA score in 5 out of 6 tasks, owing to multi-task learning and integration of FDG-PET/CT imaging data. Additionnaly, the proposed AI system, centered around BSN, provides automated prostate segmentation, uncertainty quantification, personalized feature-based explanations, and *dynamic predictions*.

## 4 Methods

### 4.1 Cohort description

The study cohort consists of 295 patients diagnosed with high-grade PCa at biopsy, with high-grade defined by a pre-treatment global Gleason score ≥ 8. These individuals underwent RP along with pelvic lymph node dissection (PLND) between 2011 and 2020 at our tertiary care center in Quebec City, Canada, and received follow-up care at the Urology department of this hospital (see Supplementary Table 8 for median follow-up times). Patients’ descriptive measurements include pre-operative FDG-PET/CT images, a ground truth annotation of the prostate on the CT, clinical variables, and outcomes information. FDG-PET/CT was performed approximately 75 minutes after the administration of 300–500 MBq FDG, with oral contrast, from base of skull to upper thighs, on a Biograph 6 PET/CT system (Siemens Healthcare, Erlangen, Germany) [58]. The PET/CT scan served the dual purpose of offering a detailed anatomical frame of reference and visualizing the distribution of FDG [66]. The clinical variables include age, pre-operative PSA level, primary and secondary Gleason scores, global Gleason score, and clinical stage (see Supplementary Table 1–6 for a detailed descriptive analysis of clinical features for each task). The available outcomes are LNI, BCRFS, MFS, dADT-FS, CRPC-FS, and PCSS following RP (see Supplementary Table 7, 8 for survival time analysis, Supplementary Fig. 1 for correlation between tasks & Supplementary Fig. 2, 5, 8, 11, 14 for descriptive analysis of the outcomes). Note that BCR is defined as a rising PSA of > 0.2 ng/ml or the initiation of a secondary treatment in response to elevated PSA levels following RP.

### 4.2 Preprocessing

The preprocessing of clinical features and images is conducted with the `DELIA`^1^ (an in-house developed package) and `MONAI` [67] Python libraries (see Fig. 7a). The clinical stage is missing in 34 cases and the PSA level in 1 case. Both are imputed using an iterative imputer with a random forest estimator implemented with the `scikit-learn` [48] Python library. Approximately 20% of all cases exhibit some missing PET attributes, necessitating the following imputations for converting PET volume into SUV [20]: body weight is assumed to be 75 kg, scan injection delay is set to 105 minutes (a conservative value), and the total injected dose is assumed to be 420 MBq. The clipping thresholds on the CT image in Hounsfield unit (HU) are determined by analyzing the HU range for prostate voxels across the images in the *learning set* and removing the 0.2% outliers [68]. The resulting HU values range is [−178, 244], so the [−200, 250] range is arbitrarily selected as a slightly larger range. The PET image clipping threshold (25) is determined based on the maximum observed SUV value (24.9) in the images of the *learning set*, as assessed by a nuclear medicine physician. This capping of SUV values at 25 helps to reduce the impact of any variability in tracer uptake and ensures a more reliable and reproducible assessment of metabolic activity. To enhance the model’s ability to generalize, data augmentation [69] is applied during the training process, meaning that each batch of data is augmented before being fed into the model. The augmentation process includes Gaussian noise, axial flipping, and axial rotation, each with a 50% probability of application (see Fig. 7b).

**Fig. 7.**
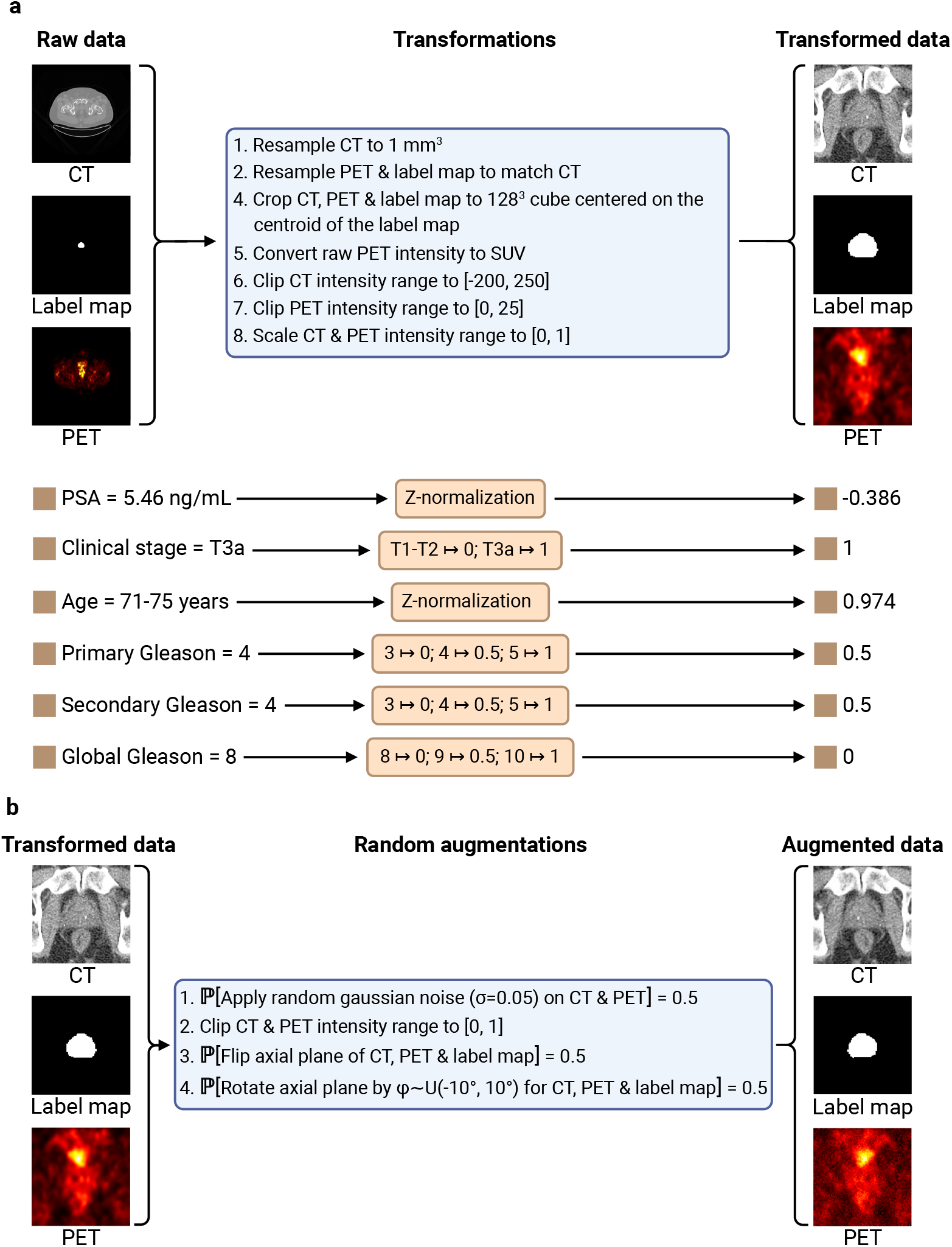
Data preprocessing pipeline. (a) Data transformation pipeline. Following a 1 mm^3^ resampling of the CT image, the PET image and the manual prostate segmentation map (label map) are resampled to align with the voxels of the CT image. The position of the centroid of the label map, which corresponds to the centroid of the prostate, is computed and images are cropped to a 128 mm^3^ cube centered on this position. The PET volume is converted into standardized uptake value (SUV) [20] image. The intensity of the CT and PET images are clipped to [-200, 250] and [0, 25] respectively (see Methods, section *Preprocessing* for the clipping range selection methodology), and then mapped to [0, 1]. Continuous clinical features are standardized using z-normalization, while categorical features are mapped to numerical values using ordinal encoding. (b) Imaging data augmentation pipeline. After applying a small amount of random noise with 50% probability, the images are clipped to [0, 1] to ensure that the intensity of the transformed images remains in the same range as the untransformed images. Flipping and rotation are then applied with 50% probability.

### 4.3 Experimental setup

The *learning set*, generated by extracting 85% of the patients in the *full dataset*, is used to run experiments to search for the best model (see Fig. 8a for experimental framework). The *holdout set*, generated from the remaining 15% of the patients, is kept hidden until the final (best) model is selected and ready to be evaluated. The *learning set* and *holdout set* are compared thoroughly by performing: a comparison between the Kaplan-Meier curves for each task (see Supplementary Fig. 2b, 5b, 8b, 11b & 14b), a comparison between the distribution of clinical features and event indicators (see Supplementary Fig. 17 & 18), and statistical analyses of features and outcomes (see Supplementary Table 9 & 10).

**Fig. 8.**
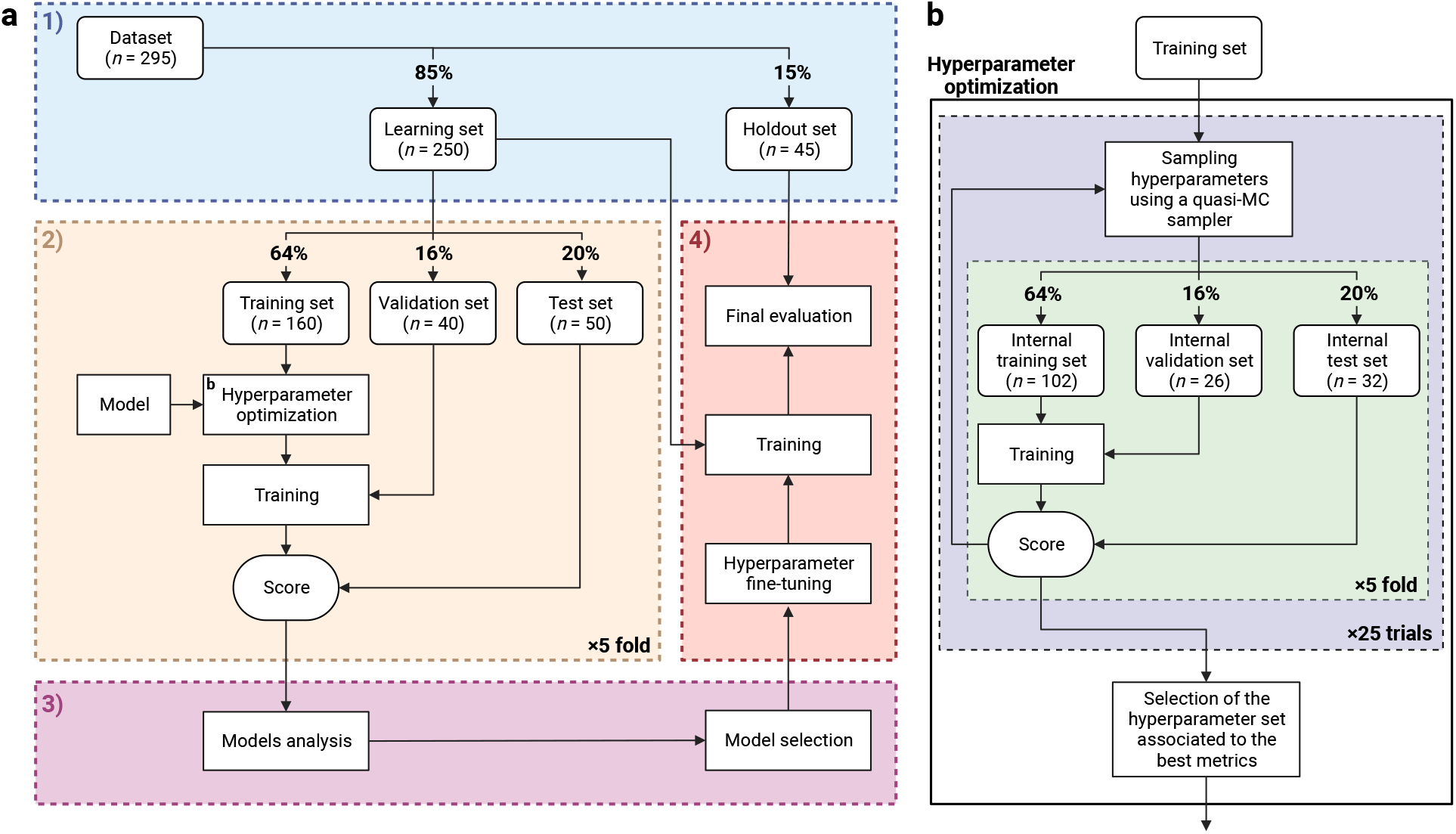
Experimental setup [71]. (a) Overview of the model selection process. 1) Stratified division of the *dataset* into a *learning set* and a *holdout set*. See Supplementary Fig. 17 & 18 and Supplementary Table 9 & 10 for visual and statistical comparison between the two generated sets. Stratification is based on LNI class labels and BCR-FS event indicators. 2) Evaluation of the models on 5 *test sets* using stratified 5-fold cross-validation. 3) Comparison and selection of the models based on performance and interpretability. The performance is measured using the mean and standard deviation of the scores on the 5 *test sets*. 4) Final evaluation of the selected model on the *holdout set*. (b) Detailed diagram of the *Hyperparameter optimization* box shown in section 2 of *panel a*. Hyperparameter optimization is performed automatically using the quasi-Monte Carlo (MC) sampler from the `BoTorch` [72] Python library, which is used under the framework of the `Optuna` [73] Python library. A total of 25 sets of hyperparameter values are sequentially sampled using the quasi-MC sampler and evaluated on the same 5 *internal test sets*. The performance is measured using the mean of the scores on the 5 *internal test sets*. The first 5 sets of hyperparameter values are randomly generated, while the subsequent ones are determined based on the performance score of the preceding sets. The set of hyperparameter values associated to the highest AUC (for classification tasks) or CI (for survival tasks) is selected. See Supplementary Table 11–17 for hyperparameter search spaces and Supplementary Table 18–20 for selected hyperparameter values.

The models are compared using stratified 5-fold cross-validation on the *learning set*, divided into *training set* (64%), *validation set* (16%), and *test set* (20%). These three sets are respectively used to optimize the models’ parameters, select the set of parameters that minimizes overfitting, and evaluate the trained models’ performance. Furthermore, the application of 5-fold cross-validation enhances the robustness of model evaluation by mitigating dependence on the training-validation-test split. For an unbiased comparison of models [70], the models’ hyperparameters are automatically optimized on the *training set* (see Fig. 8b for the hyperparameter optimization process, Supplementary Table 11–17 for hyperparameter search spaces, and Supplementary Table 18–20 for selected hyperparameter values). The model selection is based on the models’ interpretability and performance, measured using the empirical mean and standard deviation of the scores on the 5 *test sets*. The selected model is finally trained with the *learning set* and evaluated on the *holdout set*. Note that the models are trained on multiple NVIDIA A100 Tensor Core GPUs (80 GB).

### 4.4 Tasks implementation

The mathematical notation in this section adheres to that presented in the Deep Learning book [74], with the exception that we employ 2D matrix notation for 3D tensors as well. Following the book’s notation, the correspondence between italicized symbols (e.g. *x*, ***x, X***, …) and their non-italicized counterparts (e.g. x, **x, X**, …) is that a given italicized variable *x* represents an observation of the non-italicized random variable x, i.e., *x* ∈ supp(x) where supp(x) denotes the set of values that the random variable x can take. Note that italicized subscripts (e.g. *x*_*i*_, *x*_*j*_, *x*_*k*_, …) serve as indices, while non-italicized subscripts (e.g. *x*_a_, *x*_b_, *x*_c_, …) are used to supplement the definition of the given mathematical object, particularly to specify the context or concept to which that object refers.

#### 4.4.1 Binary classification

The binary classification data of a patient *i* corresponds to the pair 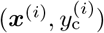 where ***x***^(*i*)^ is a vector containing the values of the patient’s risk factors (features), and 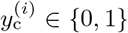 is the observed class. The binary classification task consists in modeling the probability distribution *p*(y_c_ = 1 | **x**) as a neural network *f*_c_, i.e., *f*_c_(***x***) ≈ *p*(y_c_ = 1 | **x** = ***x***). The loss function used to train this neural network is the weighted binary cross-entropy (BCE) [75]

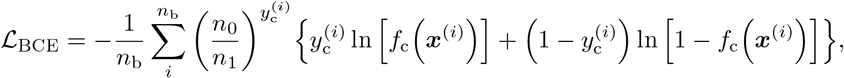

where *n*_b_ denotes the number of patients contained in the batch on which the loss is calculated, and *n*_0_ and *n*_1_ are respectively the number of patients with y_c_ = 0 and y_c_ = 1 in the set used to train the model. This scaling factor helps to provide more balanced predictions. The performance metrics used for the classification task are the area under the ROC curve (AUC) [52] and the binary balanced accuracy (BA) [76].

#### 4.4.2 Survival analysis

The time-to-event data (or survival data) of a patient *i* corresponds to the triplet 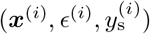 where ***x***^(*i*)^ is a vector containing the values of the patient’s risk factors (features), *ϵ*^(*i*)^ is the event indicator (or censoring indicator) so that

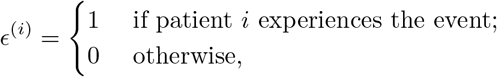

and 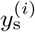 is the observed time, which represents either the survival time 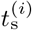 or the censoring time 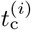, i.e.,

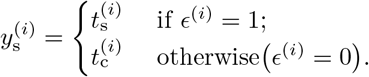

The survival analysis task consists in modeling the survival function *s*(*t*, ***x***) = *p*(t_s_ ≥ *t* | **x** = ***x***), where *t* is a duration of time (in months). The most common approach is to use the Cox proportional hazards model [77], which provides a semi-parametric specification of the hazard rate

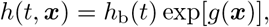

where *h*_b_(*t*) is a non-parametric baseline hazard and *g*(***x***) is the risk function. The survival function can then be retrieved by

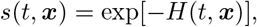

where *H*(*t*, ***x***) is the cumulative hazard defined as

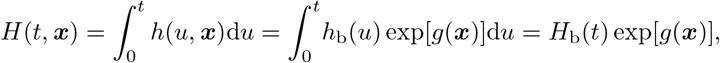

where *H*_b_(*t*) is the cumulative baseline hazard. To incorporate deep learning methods into survival analysis, the risk function *g* is modeled with a neural network *f*_s_ as *f*_s_(***x***) ≈ *g*(***x***). Indeed, the properties of the survival function, namely *s*(*t* = 0, ***x***) = 1, *s*(*t* → ∞, ***x***) = 0, and *s*(*t*, ***x***) monotonically decreasing, make it difficult to model directly as a neural network, so the risk function *g* is modeled instead [78]. The loss function used to train the neural network *f*_s_ is the negative partial log-likelihood (NPLL)

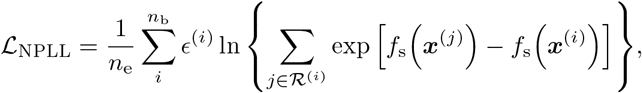

where *n*_b_ denotes the number of patients contained in the batch on which the loss is calculated, *n*_e_ is the number of events (i.e. patients with *ϵ* = 1) in the batch, and ℛ^(*i*)^ denotes the set of all individuals in the batch who are at risk at time 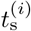 (i.e. not censored and have not experienced the event before time 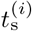 [79].

The cumulative baseline hazard can be estimated by the Breslow estimator [80]

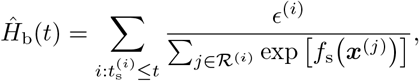

using the implementation from the `scikit-survival` [81] Python library. Note that Breslow’s method is used for handling tied event times. Three performance metrics are used to evaluate survival tasks: the concordance index (CI) [82], the CI based on inverse probability of censoring weights (CICW) [83], and the cumulative/dynamic AUC (CDA) [84]. CI is defined as the proportion of all pairs of individuals for which predictions and outcomes agree; CICW is an alternative to the CI estimator that is unbiased with respect to the distribution of censoring times in the test data; and CDA quantifies a model’s ability to distinguish subjects who fail before a given time from subjects who fail after that time [81].

#### 4.4.3 Segmentation

The segmentation data of a patient *i* corresponds to the pair (***X***^(*i*)^, ***Y*** ^(*i*)^) where ***X***^(*i*)^ is a 3D image (a tensor) and ***Y*** ^(*i*)^ is a 3D ground truth segmentation map where the voxel (*j, k, ℓ*) is denoted by 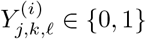 with extraprostatic and intraprostatic voxels taking values of 0 and 1, respectively. The segmentation task consists in modeling the probability distribution *p*(Y_*j,k,ℓ*_ = 1 | **X**) of all voxels. The prediction 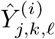 for the voxel (*j, k, ℓ*) of a patient *i* is given by

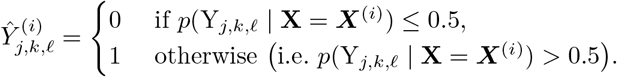

The probability distribution *p* is modeled as a neural network *f*_seg_ with *f*_seg_(***X***) ≈ *p*(Y_*j,k,ℓ*_ = 1 | **X** = ***X***), and the loss function used to train the neural network *f*_seg_ is the Dice loss

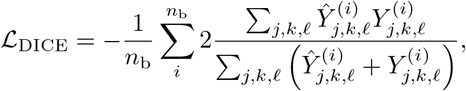

where *n*_b_ denotes the number of patients contained in the batch on which the loss is calculated. The performance metric used for the segmentation task is the Dice similarity coefficient (DSC) [85] implemented by the `MONAI` [67] Python library.

### 4.5 Risk groups

For the classification task, the risk group *g*_c_ ∈ {0, 1} of a patient *i* is determined based on the predicted positive class probability *f*_c_ (***x***^(*i*)^) as follows

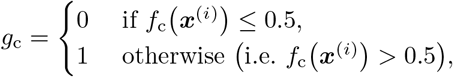

where low- and high-risk correspond to *g*_c_ = 0 and *g*_c_ = 1, respectively. For survival tasks, the risk group *g*_s_ is determined based on the predicted risk *f*_s_ (***x***^(*i*)^), with the risk threshold computed on the *learning set* using percentile thresholds [86]. This process is performed individually for each survival task. The lower percentile threshold is

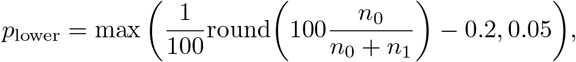

where *n*_0_ an *n*_1_ are the number of patients in the *learning set* with *ϵ*^(*i*)^ = 0 and *ϵ*^(*i*)^ = 1, respectively; the upper threshold is

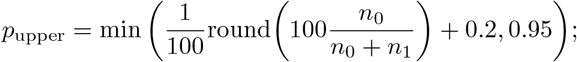

and the following percentile thresholds are therefore tested

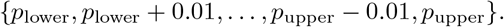

The selected risk threshold *r* is the one associated with the percentile threshold *p* that maximized the significance of the separation in the *learning set* by the log-rank test [50, 51], i.e., the threshold that minimizes the *p*-value between the two risk groups. The risk group is therefore defined as

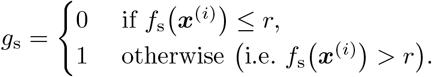

### 4.6 Handcrafted radiomic features

The U-Net, trained with a physician’s manual contours, automatically generates a segmentation map of the prostate from a CT image (see Fig. 9a for the visualization of the feature maps in the different layers of the U-Net). Using voxels from both CT and PET images within the segmented region, 200 radiomic features are extracted (see Fig. 3a for HCR extraction pipeline) with the `pyradiomics` [47] Python library (see Supplementary Table 21 & 22 for the extraction parameters on the CT and PET images, respectively). The extracted features are compliant with the Image Biomarker Standardization Initiative (IBSI) [87] except for the computed kurtosis of the distribution of voxels [47]. The Gini importance [46] of each feature is then determined using a random forest classifier with 10,000 trees implemented with the `scikit-learn` [48] Python library and trained to predict a single task using the 200 extracted radiomic features. The 6 features with the highest Gini importance are selected (see Supplementary Fig. 19 for selected features), six corresponding to the number of clinical features.

**Fig. 9.**
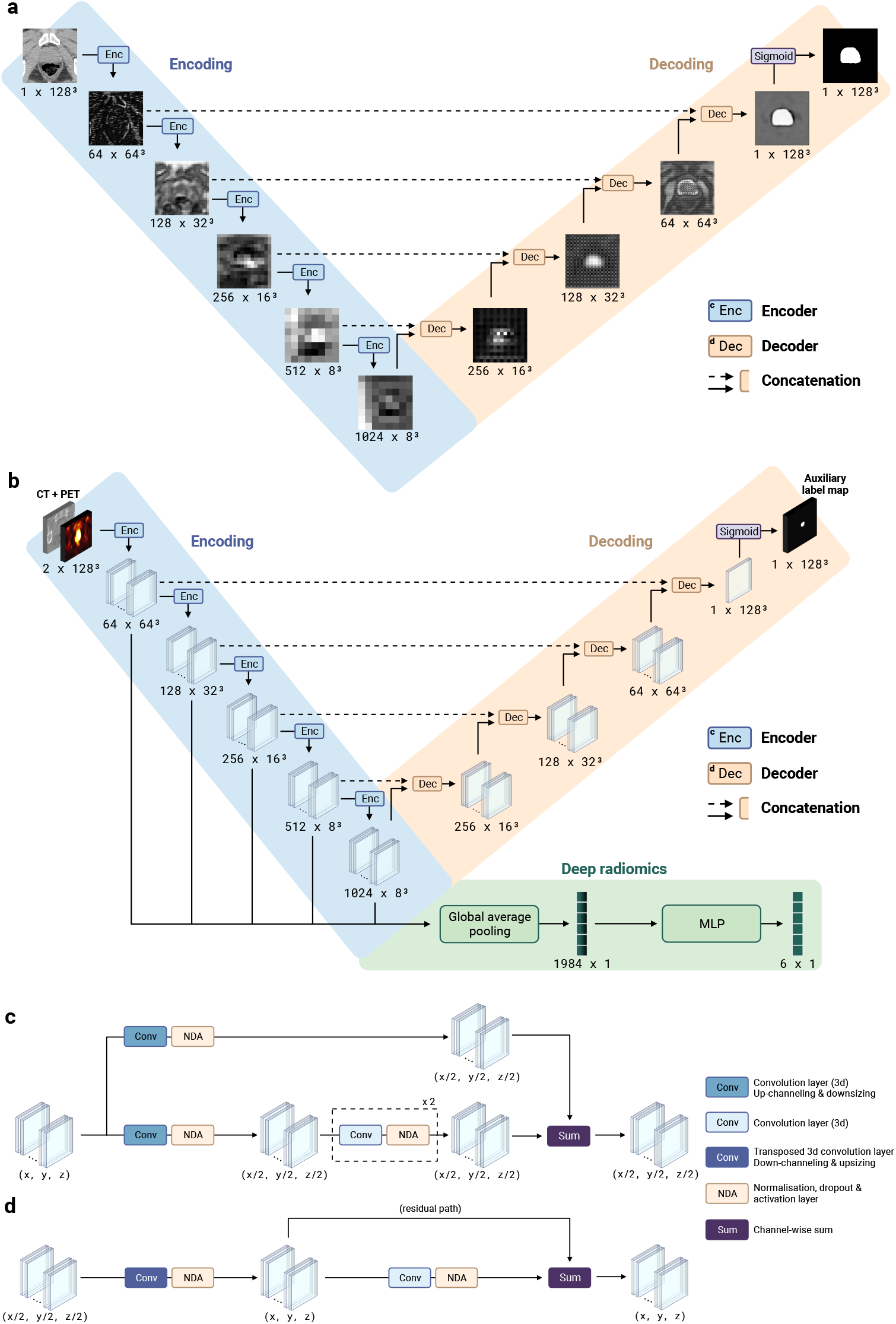
Architecture of the convolutional neural networks. (a) 3D U-Net [91] with residual units [92]. Size refers to *channels × spatial dimensions*. A grid pattern in the feature maps of the decoder appears due to padding in transposed convolution operations. See Supplementary Table 11 & 20 for hyperparameters. (b) Architecture of the 3D U-NEXtractor [49] with residual units [92]. The model is trained to segment the prostate and extract deep radiomic features simultaneously. The segmentation task guides the extraction of prognostically significant features within the prostate region. See Supplementary Table 11 & 19 for hyperparameters. (c) Encoder block. This block represents a function that reduces the size of each dimension of the input tensor by a factor of 2 using a strided convolution. (d) Decoder block. This block increases the size of each dimension of the input tensor by a factor of 2 using a strided transposed convolution.

### 4.7 Models

#### 4.7.1 Baseline models

The two baseline models are the Memorial Sloan Kettering Cancer Center (MSKCC) nomogram [10] and the Cancer of the Prostate Risk Assessment (CAPRA) score [11]. Both of these nomograms rely solely on CD and do not incorporate any imaging information. The mathematical model used by the MSKCC nomogram is a survival logistic regression while the CAPRA score is a straightforward 0 to 10 value, i.e., a linear map between the clinical features and a single number. Both models are implemented as web-based applications, and so the `prostate-nomograms`^2^ Python library was developed and used to speed up the prediction process for a very large number of patients. Indeed, the statistical models of the nomograms are reproduced in Python which allows one to calculate in a few seconds the probabilities and the scores of thousands of patients. The coefficients of the models are read from the websites and then used for the calculations.

#### 4.7.2 Deep neural networks

The neural networks are implemented and trained with the `PyTorch` [88], `MONAI` [67], and `Bayesian-Torch` [89] Python libraries. The architecture of the feed-forward neural networks, used to map a vector of features to a clinical outcome, is a multi-layer perceptron (MLP) [90]. To map a 3D image to a 3D segmentation map of the prostate, a 3D U-Net [91] with residual units [92] is employed (see Fig. 9a). The encoder and decoder architectures both use strided convolution (see Fig. 9c & 9d). Lastly, to simultaneously map a 3D image to a clinical outcome and a 3D segmentation map, the 3D U-NEXtractor [49] with residual units [92] is used (see Fig. 9b). This method builds on multi-task learning [33], which refers to sharing representations between related tasks to enable a model to generalize better. Indeed, this approach aims to use segmentation to guide the network in capturing clinically significant features in the prostatic region. The multi-task architecture consists of a 3D U-Net with an additional radiomic branch (see green-colored branch in Fig. 9b). This radiomic branch is connected to several layers of the encoding path to gather information at several levels of complexity. The connection consists of a global averaging layer to aggregate spatial information into a vector of 1984 DLRs, which is then reduced to 6 DLRs by a trainable MLP. For the hyperparameters of the selected models, see Supplementary Table 11 & 18–20.

#### 4.7.3 Bayesian neural networks

Bayesian neural networks (BNN) expand upon traditional neural networks by incorporating principles from Bayesian probability theory into the neural network framework. Let ***θ*** be the vector of model parameters, Θ the space of said parameters and 𝕏 the *learning set*. In the context of traditional neural networks, model parameters ***θ*** are typically treated as fixed yet unknown values, and training aims to recover their true underlying values. However, in the context of BNNs, these parameters are most often viewed as random variables characterized by unknown probability distributions. This approach better accounts for model uncertainty. Therefore, by recovering the posterior distribution over the model parameters *p*(***θ*** | 𝕏), one could subsequently derive the posterior predictive distribution over the model’s prediction **ŷ** given the input features ***x***^(*i*)^ of a new patient *i*, i.e., (***x***^(*i*)^, ***y***^(*i*)^) ∉ 𝕏:

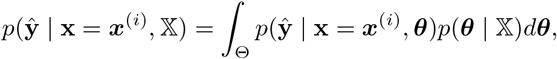

thus allowing for probabilistic prediction and knowledge of the uncertainty of a given prediction of the model [38]. But for the complex models used in deep learning, the sheer size of the parameter space Θ would render this integral intractable. Besides, the posterior distribution over the weights *p*(***θ*** | 𝕏) typically cannot be analytically recovered [93]. In practice, a single parameter value is used upon inference.

The Bayes-by-backpropagation method [94] proposes a practical implementation of variational inference to sidestep this problem and approximate the underlying probabilistic process of model prediction. The idea of this method is to assign to each weight an arbitrary variational approximation *q*(*θ*) of its posterior distribution *p*(*θ* | 𝕏). Upon training, the distributions *q*(***θ***) are learned and an exact parameter value is sampled from these learned distribution upon inference. A simple known form for *q*(***θ***), i.e., a Gaussian distribution 𝒩 [37], is chosen to serve as a tractable approximation of the true posterior distribution *p*(***θ*** | 𝕏). That is, *q*(***θ***) = 𝒩 (***θ***; *µ, σ*), where *µ* is the mean of the distribution and *σ* is the standard deviation. As a result, learning the distribution *q*(*θ*) of each weight *θ* is equivalent to learning its corresponding pair of parameters (*µ, σ*), which effectively doubles the number of parameters to learn. In order not to have gradient descent go through internal random nodes, the reparametrization trick [95] is applied so that randomness is introduced as an external input of the latent space of ***θ*** instead. Because the parameters ***θ*** are drawn from *q*(***θ***), every forward pass of the model is different, even if the input ***x*** is constant. Taking statistics such as the mean or the variance of the predictions collected from the multiple forward passes gives a better insight into the underlying probabilistic process. This ensemble of predictions can be used to quantify the uncertainty of the model for a given prediction in situations where uncertainty is critical, such as in most medical settings.

#### 4.7.4 Sequential Network

The model that groups all single-task models together in a multi-task framework is the SN (or its Bayesian counterpart BSN) (see Fig. 3). Each single-task model has 3 different inputs: clinical data (mandatory), imaging data (task-specific), and predictions from previous single-task models (task-specific), giving rise to the term *sequential*. The sequence of tasks was determined based on the natural progression of PCa (see Fig. 1b). This approach aims to use the prediction of short-term outcomes to improve the prediction of long-term outcomes, as they are correlated (see Supplementary Fig. 1). The architecture of the SN naturally gives rise to *dynamic predictions*, i.e., predictions that are refined over time according to the clinical results of previous tasks. The formal definition of *dynamic predictions* follows. Let {*k*_1_, …, *k*_*n*_} be the set of tasks in the SN. For each of these tasks *k*_*i*_, the single-task neural network (contained in the SN) that performs the task *k*_*i*_ is denoted by *f*_*i*_, the output value of *f*_*i*_ by *o*_*i*_, the input data features by ***x***_*i*_, and the set of previous connected single-task models by *S*_*i*_ = {*f*_1_, …, *f*_*m*_} ⊆ {*f*_1_, …, *f*_*i*−1_}. For a prognosis made at time *t*_p_, the vector function ***a***_*i*_(*t*_p_) that defines the additional input to *f*_*i*_ that comes from the output of the models in *S*_*i*_ is ***a***_*i*_(*t*_p_) = (*a*_1_(*t*_p_), …, *a*_*m*_(*t*_p_))^⊤^, where *a*_*j*_(*t*_p_) is defined as

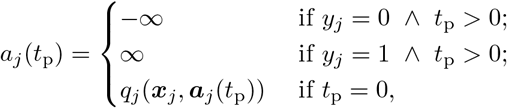

for binary classification tasks, where *y*_*j*_ ∈ {0, 1} i s the observed class for the task *k*_*j*_, and *q*_*j*_ corresponds to the neural network *f*_*j*_ without the activation function in the output layer, i.e., *f*_*j*_ = *σ* ◯ *q*_*j*_ where *σ* is the sigmoid function. For survival tasks, *a*_*j*_ (*t*_p_) is defined as

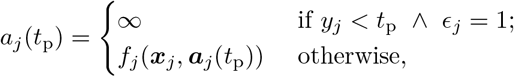

where *ϵ*_*j*_ is the event indicator and *y*_*j*_ is the observed time in this case. For reasons of computational stability, infinities are replaced by 5 and 10 in the code for classification and survival tasks, respectively. The *dynamic prediction* of the outcome *o*_*i*_ made at time *t*_p_ is then

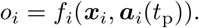

In a clinical context, *dynamic predictions* are likely to be used following the occurrence of an event to obtain new and, ideally, better predictions of subsequent events.

### 4.8 Interpretability

As confusion is easily created when discussing interpretability and explainability, it is important to define both prior to their discussion. In this case, interpretability refers to the analysis and understanding of a model’s decision-making process and explainability refers to the presentation of such a process in a manner that allows for a clear understanding of the link between input and output data [96]. As such, SHAP and SurvSHAP(*t*) values are meant to improve explainability. However, the portrayal of these values correlated with the input data and compared with clinical observations can improve interpretability. In the following, SHAP [56] values are discussed as the main source of interpretability and explainability. However, SurvSHAP(*t*) [57] values serve a similar purpose and are not discussed as thoroughly to reduce redundancy.

Originating from the Shapley [97] values used in game theory, SHAP values allow for intuitive analysis of the importance attributed to each input features *x*_*i*_ by a neural network *f* to predict the output *f* (*x*_1_, …, *x*_*d*_). Whereas Shapley values are determined by removing an input feature to determine its contribution to the output, SHAP values replace the said input feature by sampling its value from the probability distribution of that feature on the *learning set* 𝕏, thus achieving a similar analysis to the Shapley method over many samples. For a given patient, the sum of all its SHAP values added to the expected value of the prediction on the whole dataset gives the predicted value for that patient, i.e. given that the SHAP value of the feature *x*_*i*_ is *ϕ*_*i*_, the prediction is given by

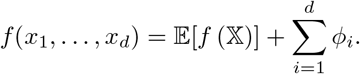

In order to compute the desired SHAP values, the `CAPTUM` [98] Python library is employed. Once SHAP values are computed, the `SHAP` [56] Python library is utilized to create appropriate and readable graphs. It is through this step that the model becomes more interpretable, by comparing and analyzing importance values in relation to input data. SurvSHAP(*t*) values are computed using the `SurvSHAP`(*t*) [57] Python library and the related graphs are created using in-house code. The graphs created from either SHAP or the in-house code serve one of three purposes:

- **Patient-wise explanation**. Graphing a patient’s SHAP values (e.g. through waterfall plots such as Fig. 6e) allows for a straightforward explanation of the model’s prediction in a specific case. This is the most efficient method to comprehend how a specific output is obtained and can serve as a foothold to begin the analysis of a prediction.
- **Dataset-wise interpretation**. Graphing the SHAP values associated with the dataset (e.g. through beeswarm plots such as Fig. 5a) can show tendencies within the importance values. These tendencies can be compared with clinical observations to verify the model’s quality or analyze its decision-making process.
- **Sequence-specific analysis**. Since multiple neural networks were used in succession to compute multiple tasks, it is possible to compute SHAP values at different points within the sequential neural network. The succession of SHAP values makes it possible to interpret the sequential architecture and see if each network is affected by the preceding ones. For example, the presence of HCR in a task that does not receive it as an input indicates that the permeation of input data has an effect on the network’s decisions, albeit one whose quality is difficult to determine.

## Data Availability

Due to participant confidentiality and privacy concerns, data are available upon reasonable written request to.

https://github.com/MedPhysUL/ProstateCancerPrognosisAI

https://github.com/MedPhysUL/delia

https://github.com/MedPhysUL/prostate-nomograms

## 6 Code availability

All the codes and guidance can be found on our organization’s GitHub at https://github.com/MedPhysUL. More specifically, the main code is available at https://github.com/MedPhysUL/ProstateCancerPrognosisAI, DELIA at https://github.com/MedPhysUL/delia, and the implementation of the baseline nomograms at https://github.com/MedPhysUL/prostate-nomograms.

## 7 Acknowledgements

This work was supported by the Natural Sciences and Engineering Research Council of Canada (NSERC), Fonds de recherche en Santé du Québec, the Fonds de recherche du Québec (FRQNT) and Fondation du CHU de Québec. We would like to thank VALERIA Université Laval for access to their GPU cluster. We would also like to acknowledge the use of Biorender for the creation of most figures.

## 8 Author information

F.P. designed the clinical protocol and objectives (with L.A.), collected the clinical data, and carried out patient follow-up and ethical approval. N.T., D.B.T., H.H., and B.N. performed data curation. N.T., D.L., and F.R. contoured the prostate on the CT images and revised the manuscript. M.L., L.A., and M.V. designed the approach. M.L. developed the `DELIA` and `prostate-nomograms` Python libraries, implemented the preprocessing, training, and hyperparameters optimization schemes, developed the underlying framework of the models, performed all experiments, and analyzed the results. R.B. implemented the segmentation neural networks, constructed the Bayesian counterpart of all models, and developed visualization tools. F.D. implemented the explanation methods and analyzed their results. N.R. provided the idea for the experimental setup (model selection process) and helped build the software architecture. M.V. proposed the architecture of the sequential network. L.A., M.V., and F.P. supervised the project. M.L., N.T., R.B., and F.D. wrote the manuscript. All authors reviewed the manuscript.

## 9 Ethics declarations

The research protocol was approved by the Ethical committee of CHU de Québec-Université Laval Hospital, Quebec City, Quebec, Canada (IRB # 2018-3667).

## Footnotes

1 https://github.com/MedPhysUL/delia

2 https://github.com/MedPhysUL/prostate-nomograms

## Notes

### Competing Interest Statement

The authors have declared no competing interest.

### Author Declarations

The research protocol was approved by the Ethical committee of CHU de Quebec-Universite Laval Hospital, Quebec City, Quebec, Canada (IRB # 2018-3667).

