## Supplementary Material for "Multi-task Bayesian Model Combining FDG-PET/CT Imaging and Clinical Data for Interpretable High-Grade Prostate Cancer Prognosis"

#### 1 Cohort description

##### 1.1 Descriptive analysis of clinical features

**Supplementary Table 1** Descriptive analysis of the clinical features for the LNI task on the *full dataset*, the *learning set*, and the *holdout set*. The $p$-values are computed using the Mann-Whitney U test [1] for continuous clinical features (age and psa) and the $\chi^2$ test [2] for categorical features (clinical stage, global Gleason score, primary Gleason score, and secondary Gleason score) using `scipy` [3] Python library.

| | Full dataset | | | | Learning set | | | | Holdout set | | | |
|---|---|---|---|---|---|---|---|---|---|---|---|---|
| Feature | All | Negative | Positive | $p$-value | All | Negative | Positive | $p$-value | All | Negative | Positive | $p$-value |
| | $n = 295$ | $n = 209$ | $n = 86$ | | $n = 250$ | $n = 177$ | $n = 73$ | | $n = 45$ | $n = 32$ | $n = 13$ | |
| | 100.0 % | 70.8 % | 29.2 % | | 100.0 % | 70.8 % | 29.2 % | | 100.0 % | 71.1 % | 28.9 % | |
| Age [years] | | | | 0.8035 | | | | 0.7697 | | | | 0.9600 |
| Mean (Median) | 65.4 (66.0) | 65.4 (66.0) | 65.3 (66.0) | | 65.6 (66.0) | 65.6 (66.0) | 65.4 (66.0) | | 64.4 (66.0) | 64.4 (65.5) | 64.5 (66.0) | |
| Min - Max | 48.0 - 80.0 | 48.0 - 80.0 | 48.0 - 78.0 | | 48.0 - 80.0 | 48.0 - 80.0 | 48.0 - 78.0 | | 50.0 - 76.0 | 50.0 - 76.0 | 53.0 - 75.0 | |
| PSA [ng/ml] | | | | 0.0572 | | | | 0.0854 | | | | 0.5903 |
| Mean (Median) | 11.0 (7.4) | 9.9 (7.1) | 13.5 (8.1) | | 11.5 (7.4) | 10.3 (7.1) | 14.4 (8.2) | | 8.0 (7.4) | 7.9 (7.0) | 8.3 (7.4) | |
| Min - Max | 1.1 - 155.3 | 1.1 - 155.3 | 1.1 - 110.0 | | 1.1 - 155.3 | 1.4 - 155.3 | 1.1 - 110.0 | | 1.1 - 17.7 | 1.1 - 17.7 | 5.5 - 16.7 | |
| Clinical stage | | | | $< 0.0001$ | | | | $< 0.0001$ | | | | 0.5913 |
| T1-T2 | 230 (88.1) | 176 (95.1) | 54 (71.1) | | 196 (87.9) | 150 (95.5) | 46 (69.7) | | 34 (89.5) | 26 (92.9) | 8 (80.0) | |
| T3a | 31 (11.9) | 9 (4.9) | 22 (28.9) | | 27 (12.1) | 7 (4.5) | 20 (30.3) | | 4 (10.5) | 2 (7.1) | 2 (20.0) | |
| Global Gleason | | | | 0.0020 | | | | 0.0031 | | | | 0.2363 |
| 8 | 188 (63.7) | 146 (69.9) | 42 (48.8) | | 160 (64.0) | 124 (70.1) | 36 (49.3) | | 28 (62.2) | 22 (68.8) | 6 (46.2) | |
| 9 | 106 (35.9) | 62 (29.7) | 44 (51.2) | | 90 (36.0) | 53 (29.9) | 37 (50.7) | | 16 (35.6) | 9 (28.1) | 7 (53.8) | |
| 10 | 1 (0.3) | 1 (0.5) | 0 (0.0) | | 0 (0.0) | 0 (0.0) | 0 (0.0) | | 1 (2.2) | 1 (3.1) | 0 (0.0) | |
| Primary Gleason | | | | 0.3240 | | | | 0.4349 | | | | 0.5732 |
| 3 | 16 (5.4) | 13 (6.2) | 3 (3.5) | | 14 (5.6) | 11 (6.2) | 3 (4.1) | | 2 (4.4) | 2 (6.2) | 0 (0.0) | |
| 4 | 268 (90.8) | 190 (90.9) | 78 (90.7) | | 230 (92.0) | 163 (92.1) | 67 (91.8) | | 38 (84.4) | 27 (84.4) | 11 (84.6) | |
| 5 | 11 (3.7) | 6 (2.9) | 5 (5.8) | | 6 (2.4) | 3 (1.7) | 3 (4.1) | | 5 (11.1) | 3 (9.4) | 2 (15.4) | |
| Secondary Gleason | | | | 0.0324 | | | | 0.0247 | | | | 0.5486 |
| 3 | 2 (0.7) | 1 (0.5) | 1 (1.2) | | 0 (0.0) | 0 (0.0) | 0 (0.0) | | 2 (4.4) | 1 (3.1) | 1 (7.7) | |
| 4 | 178 (60.3) | 136 (65.1) | 42 (48.8) | | 152 (60.8) | 116 (65.5) | 36 (49.3) | | 26 (57.8) | 20 (62.5) | 6 (46.2) | |
| 5 | 115 (39.0) | 72 (34.4) | 43 (50.0) | | 98 (39.2) | 61 (34.5) | 37 (50.7) | | 17 (37.8) | 11 (34.4) | 6 (46.2) | |

**Supplementary Table 2** Descriptive analysis of the clinical features for the BCR-FS task on the *full dataset*, the *learning set*, and the *holdout set*. The $p$-values are computed using the Mann-Whitney U test [1] for continuous clinical features (age and psa) and the $\chi^2$ test [2] for categorical features (clinical stage, global Gleason score, primary Gleason score, and secondary Gleason score) using `scipy` [3] Python library.

| | Full dataset | | | | Learning set | | | | Holdout set | | | |
|---|---|---|---|---|---|---|---|---|---|---|---|---|
| Feature | All | Negative | Positive | $p$-value | All | Negative | Positive | $p$-value | All | Negative | Positive | $p$-value |
| | $n = 289$ | $n = 129$ | $n = 160$ | | $n = 245$ | $n = 112$ | $n = 133$ | | $n = 44$ | $n = 17$ | $n = 27$ | |
| | 100.0 % | 44.6 % | 55.4 % | | 100.0 % | 45.7 % | 54.3 % | | 100.0 % | 38.6 % | 61.4 % | |
| Age [years] | | | | 0.1391 | | | | 0.1541 | | | | 0.7444 |
| Mean (Median) | 65.4 (66.0) | 66.1 (67.0) | 64.9 (65.0) | | 65.6 (66.0) | 66.2 (67.0) | 65.0 (65.0) | | 64.5 (66.0) | 65.1 (66.0) | 64.1 (66.0) | |
| Min - Max | 48.0 - 80.0 | 48.0 - 80.0 | 48.0 - 78.0 | | 48.0 - 80.0 | 48.0 - 80.0 | 48.0 - 78.0 | | 50.0 - 76.0 | 53.0 - 76.0 | 50.0 - 75.0 | |
| PSA [ng/ml] | | | | 0.0271 | | | | 0.0170 | | | | 0.885 |
| Mean (Median) | 11.0 (7.4) | 8.5 (7.0) | 13.0 (7.9) | | 11.6 (7.4) | 8.6 (7.0) | 14.0 (8.2) | | 7.9 (7.3) | 8.1 (7.7) | 7.8 (7.2) | |
| Min - Max | 1.1 - 155.3 | 1.1 - 48.0 | 1.4 - 155.3 | | 1.1 - 155.3 | 1.1 - 48.0 | 1.4 - 155.3 | | 1.1 - 17.7 | 1.1 - 17.7 | 2.4 - 16.7 | |
| Clinical stage | | | | 0.1518 | | | | 0.1921 | | | | 0.9320 |
| T1-T2 | 225 (87.9) | 100 (91.7) | 125 (85.0) | | 191 (87.6) | 86 (91.5) | 105 (84.7) | | 34 (89.5) | 14 (93.3) | 20 (87.0) | |
| T3a | 31 (12.1) | 9 (8.3) | 22 (15.0) | | 27 (12.4) | 8 (8.5) | 19 (15.3) | | 4 (10.5) | 1 (6.7) | 3 (13.0) | |
| Global Gleason | | | | 0.0395 | | | | 0.0291 | | | | 1.0000 |
| 8 | 184 (63.7) | 91 (70.5) | 93 (58.1) | | 156 (63.7) | 80 (71.4) | 76 (57.1) | | 28 (63.6) | 11 (64.7) | 17 (63.0) | |
| 9 | 105 (36.3) | 38 (29.5) | 67 (41.9) | | 89 (36.3) | 32 (28.6) | 57 (42.9) | | 16 (36.4) | 6 (35.3) | 10 (37.0) | |
| 10 | 0 (0.0) | 0 (0.0) | 0 (0.0) | | 0 (0.0) | 0 (0.0) | 0 (0.0) | | 0 (0.0) | 0 (0.0) | 0 (0.0) | |
| Primary Gleason | | | | 0.0944 | | | | 0.3045 | | | | 0.1702 |
| 3 | 16 (5.5) | 11 (8.5) | 5 (3.1) | | 14 (5.7) | 9 (8.0) | 5 (3.8) | | 2 (4.5) | 2 (11.8) | 0 (0.0) | |
| 4 | 263 (91.0) | 115 (89.1) | 148 (92.5) | | 225 (91.8) | 101 (90.2) | 124 (93.2) | | 38 (86.4) | 14 (82.4) | 24 (88.9) | |
| 5 | 10 (3.5) | 3 (2.3) | 7 (4.4) | | 6 (2.4) | 2 (1.8) | 4 (3.0) | | 4 (9.1) | 1 (5.9) | 3 (11.1) | |
| Secondary Gleason | | | | 0.2254 | | | | 0.2041 | | | | 0.4877 |
| 3 | 2 (0.7) | 0 (0.0) | 2 (1.2) | | 0 (0.0) | 0 (0.0) | 0 (0.0) | | 2 (4.5) | 0 (0.0) | 2 (7.4) | |
| 4 | 174 (60.2) | 83 (64.3) | 91 (56.9) | | 148 (60.4) | 73 (65.2) | 75 (56.4) | | 26 (59.1) | 10 (58.8) | 16 (59.3) | |
| 5 | 113 (39.1) | 46 (35.7) | 67 (41.9) | | 97 (39.6) | 39 (34.8) | 58 (43.6) | | 16 (36.4) | 7 (41.2) | 9 (33.3) | |

**Supplementary Table 3** Descriptive analysis of the clinical features for the MFS task on the *full dataset*, the *learning set*, and the *holdout set*. The $p$-values are computed using the Mann-Whitney U test [1] for continuous clinical features (age and psa) and the $\chi^2$ test [2] for categorical features (clinical stage, global Gleason score, primary Gleason score, and secondary Gleason score) using `scipy` [3] Python library.

| | Full dataset | | | | Learning set | | | | Holdout set | | | |
|---|---|---|---|---|---|---|---|---|---|---|---|---|
| Feature | All | Negative | Positive | $p$-value | All | Negative | Positive | $p$-value | All | Negative | Positive | $p$-value |
| | $n = 157$ | $n = 119$ | $n = 38$ | | $n = 131$ | $n = 99$ | $n = 32$ | | $n = 26$ | $n = 20$ | $n = 6$ | |
| | 100.0 % | 75.8 % | 24.2 % | | 100.0 % | 75.6 % | 24.4 % | | 100.0 % | 76.9 % | 23.1 % | |
| Age [years] | | | | 0.0312 | | | | 0.0357 | | | | 0.6038 |
| Mean (Median) | 64.7 (65.0) | 65.4 (66.0) | 62.8 (62.5) | | 64.7 (65.0) | 65.4 (66.0) | 62.7 (62.5) | | 64.7 (66.0) | 65.2 (66.5) | 63.3 (63.5) | |
| Min - Max | 48.0 - 78.0 | 48.0 - 78.0 | 48.0 - 77.0 | | 48.0 - 78.0 | 48.0 - 78.0 | 48.0 - 77.0 | | 53.0 - 75.0 | 53.0 - 75.0 | 54.0 - 73.0 | |
| PSA [ng/ml] | | | | 0.0255 | | | | 0.0277 | | | | 0.9272 |
| Mean (Median) | 12.5 (7.8) | 12.2 (7.4) | 13.4 (10.0) | | 13.4 (8.2) | 13.1 (7.5) | 14.4 (10.3) | | 7.8 (7.3) | 7.7 (7.3) | 8.0 (6.9) | |
| Min - Max | 1.1 - 155.3 | 1.1 - 155.3 | 2.6 - 85.3 | | 1.1 - 155.3 | 1.1 - 155.3 | 2.6 - 85.3 | | 3.0 - 16.7 | 3.0 - 16.7 | 5.3 - 13.3 | |
| Clinical stage | | | | 0.2874 | | | | 0.3531 | | | | 1.0000 |
| T1-T2 | 119 (83.8) | 93 (86.1) | 26 (76.5) | | 99 (83.2) | 77 (85.6) | 22 (75.9) | | 20 (87.0) | 16 (88.9) | 4 (80.0) | |
| T3a | 23 (16.2) | 15 (13.9) | 8 (23.5) | | 20 (16.8) | 13 (14.4) | 7 (24.1) | | 3 (13.0) | 2 (11.1) | 1 (20.0) | |
| Global Gleason | | | | 0.0020 | | | | 0.0076 | | | | 0.2540 |
| 8 | 101 (64.3) | 85 (71.4) | 16 (42.1) | | 85 (64.9) | 71 (71.7) | 14 (43.8) | | 16 (61.5) | 14 (70.0) | 2 (33.3) | |
| 9 | 56 (35.7) | 34 (28.6) | 22 (57.9) | | 46 (35.1) | 28 (28.3) | 18 (56.2) | | 10 (38.5) | 6 (30.0) | 4 (66.7) | |
| 10 | 0 (0.0) | 0 (0.0) | 0 (0.0) | | 0 (0.0) | 0 (0.0) | 0 (0.0) | | 0 (0.0) | 0 (0.0) | 0 (0.0) | |
| Primary Gleason | | | | 0.1414 | | | | 0.2097 | | | | 0.5656 |
| 3 | 7 (4.5) | 6 (5.0) | 1 (2.6) | | 6 (4.6) | 5 (5.1) | 1 (3.1) | | 1 (3.8) | 1 (5.0) | 0 (0.0) | |
| 4 | 145 (92.4) | 111 (93.3) | 34 (89.5) | | 122 (93.1) | 93 (93.9) | 29 (90.6) | | 23 (88.5) | 18 (90.0) | 5 (83.3) | |
| 5 | 5 (3.2) | 2 (1.7) | 3 (7.9) | | 3 (2.3) | 1 (1.0) | 2 (6.2) | | 2 (7.7) | 1 (5.0) | 1 (16.7) | |
| Secondary Gleason | | | | 0.0809 | | | | 0.0569 | | | | 0.7225 |
| 3 | 1 (0.6) | 1 (0.8) | 0 (0.0) | | 0 (0.0) | 0 (0.0) | 0 (0.0) | | 1 (3.8) | 1 (5.0) | 0 (0.0) | |
| 4 | 97 (61.8) | 79 (66.4) | 18 (47.4) | | 82 (62.6) | 67 (67.7) | 15 (46.9) | | 15 (57.7) | 12 (60.0) | 3 (50.0) | |
| 5 | 59 (37.6) | 39 (32.8) | 20 (52.6) | | 49 (37.4) | 32 (32.3) | 17 (53.1) | | 10 (38.5) | 7 (35.0) | 3 (50.0) | |

**Supplementary Table 4** Descriptive analysis of the clinical features for the dADT-FS task on the *full dataset*, the *learning set*, and the *holdout set*. The $p$-values are computed using the Mann-Whitney U test [1] for continuous clinical features (age and psa) and the $\chi^2$ test [2] for categorical features (clinical stage, global Gleason score, primary Gleason score, and secondary Gleason score) using `scipy` [3] Python library.

| | Full dataset | | | | Learning set | | | | Holdout set | | | |
|---|---|---|---|---|---|---|---|---|---|---|---|---|
| Feature | All<br>$n = 282$<br>100.0 % | Negative<br>$n = 210$<br>74.5 % | Positive<br>$n = 72$<br>25.5 % | $p$-value | All<br>$n = 239$<br>100.0 % | Negative<br>$n = 173$<br>72.4 % | Positive<br>$n = 66$<br>27.6 % | $p$-value | All<br>$n = 43$<br>100.0 % | Negative<br>$n = 37$<br>86.0 % | Positive<br>$n = 6$<br>14.0 % | $p$-value |
| Age [years] | | | | 0.8754 | | | | 0.9975 | | | | 0.4716 |
| Mean (Median) | 65.2 (65.0) | 65.3 (66.0) | 65.2 (65.0) | | 65.4 (65.0) | 65.4 (66.0) | 65.4 (65.0) | | 64.3 (65.0) | 64.6 (66.0) | 62.7 (62.5) | |
| Min - Max | 48.0 - 80.0 | 48.0 - 80.0 | 48.0 - 78.0 | | 48.0 - 80.0 | 48.0 - 80.0 | 48.0 - 78.0 | | 50.0 - 76.0 | 50.0 - 76.0 | 54.0 - 73.0 | |
| PSA [ng/ml] | | | | 0.0008 | | | | 0.0012 | | | | 0.5749 |
| Mean (Median) | 10.7 (7.3) | 9.4 (6.9) | 14.5 (9.0) | | 11.2 (7.3) | 9.7 (6.9) | 15.1 (9.2) | | 7.9 (7.4) | 7.8 (7.2) | 8.4 (8.1) | |
| Min - Max | 1.1 - 155.3 | 1.1 - 155.3 | 1.8 - 110.0 | | 1.1 - 155.3 | 1.1 - 155.3 | 1.8 - 110.0 | | 1.1 - 17.7 | 1.1 - 17.7 | 5.3 - 13.3 | |
| Clinical stage | | | | 0.0198 | | | | 0.0205 | | | | 1.0000 |
| T1-T2 | 221 (89.1) | 166 (92.2) | 55 (80.9) | | 189 (89.2) | 139 (92.7) | 50 (80.6) | | 32 (88.9) | 27 (90.0) | 5 (83.3) | |
| T3a | 27 (10.9) | 14 (7.8) | 13 (19.1) | | 23 (10.8) | 11 (7.3) | 12 (19.4) | | 4 (11.1) | 3 (10.0) | 1 (16.7) | |
| Global Gleason | | | | 0.0021 | | | | 0.0012 | | | | 0.6701 |
| 8 | 180 (63.8) | 146 (69.5) | 34 (47.2) | | 153 (64.0) | 122 (70.5) | 31 (47.0) | | 27 (62.8) | 24 (64.9) | 3 (50.0) | |
| 9 | 101 (35.8) | 63 (30.0) | 38 (52.8) | | 86 (36.0) | 51 (29.5) | 35 (53.0) | | 15 (34.9) | 12 (32.4) | 3 (50.0) | |
| 10 | 1 (0.4) | 1 (0.5) | 0 (0.0) | | 0 (0.0) | 0 (0.0) | 0 (0.0) | | 1 (2.3) | 1 (2.7) | 0 (0.0) | |
| Primary Gleason | | | | 0.1675 | | | | 0.2341 | | | | 0.1834 |
| 3 | 16 (5.7) | 13 (6.2) | 3 (4.2) | | 14 (5.9) | 11 (6.4) | 3 (4.5) | | 2 (4.7) | 2 (5.4) | 0 (0.0) | |
| 4 | 256 (90.8) | 192 (91.4) | 64 (88.9) | | 220 (92.1) | 160 (92.5) | 60 (90.9) | | 36 (83.7) | 32 (86.5) | 4 (66.7) | |
| 5 | 10 (3.5) | 5 (2.4) | 5 (6.9) | | 5 (2.1) | 2 (1.2) | 3 (4.5) | | 5 (11.6) | 3 (8.1) | 2 (33.3) | |
| Secondary Gleason | | | | 0.0336 | | | | 0.0145 | | | | 0.3211 |
| 3 | 2 (0.7) | 1 (0.5) | 1 (1.4) | | 0 (0.0) | 0 (0.0) | 0 (0.0) | | 2 (4.7) | 1 (2.7) | 1 (16.7) | |
| 4 | 169 (59.9) | 135 (64.3) | 34 (47.2) | | 144 (60.3) | 113 (65.3) | 31 (47.0) | | 25 (58.1) | 22 (59.5) | 3 (50.0) | |
| 5 | 111 (39.4) | 74 (35.2) | 37 (51.4) | | 95 (39.7) | 60 (34.7) | 35 (53.0) | | 16 (37.2) | 14 (37.8) | 2 (33.3) | |

**Supplementary Table 5** Descriptive analysis of the clinical features for the CRPC-FS task on the *full dataset*, the *learning set*, and the *holdout set*. The $p$-values are computed using the Mann-Whitney U test [1] for continuous clinical features (age and psa) and the $\chi^2$ test [2] for categorical features (clinical stage, global Gleason score, primary Gleason score, and secondary Gleason score) using `scipy` [3] Python library.

| | Full dataset | | | | Learning set | | | | Holdout set | | | |
|---|---|---|---|---|---|---|---|---|---|---|---|---|
| Feature | All<br>$n = 290$<br>100.0 % | Negative<br>$n = 267$<br>92.1 % | Positive<br>$n = 23$<br>7.9 % | $p$-value | All<br>$n = 246$<br>100.0 % | Negative<br>$n = 227$<br>92.3 % | Positive<br>$n = 19$<br>7.7 % | $p$-value | All<br>$n = 44$<br>100.0 % | Negative<br>$n = 33$<br>90.9 % | Positive<br>$n = 4$<br>9.1 % | $p$-value |
| Age [years] | | | | 0.0383 | | | | 0.0337 | | | | 0.7904 |
| Mean (Median) | 65.4 (66.0) | 65.6 (66.0) | 62.7 (62.0) | | 65.6 (66.0) | 65.8 (66.0) | 62.5 (62.0) | | 64.4 (66.0) | 64.5 (66.0) | 63.5 (63.5) | |
| Min - Max | 48.0 - 80.0 | 48.0 - 80.0 | 48.0 - 74.0 | | 48.0 - 80.0 | 48.0 - 80.0 | 48.0 - 74.0 | | 50.0 - 76.0 | 50.0 - 76.0 | 54.0 - 73.0 | |
| PSA [ng/ml] | | | | 0.0032 | | | | 0.0044 | | | | 0.4142 |
| Mean (Median) | 10.7 (7.4) | 10.3 (7.1) | 15.3 (10.4) | | 11.1 (7.4) | 10.7 (7.1) | 16.6 (11.0) | | 8.0 (7.3) | 7.9 (7.0) | 9.1 (8.7) | |
| Min - Max | 1.1 - 155.3 | 1.1 - 155.3 | 2.6 - 85.3 | | 1.1 - 155.3 | 1.1 - 155.3 | 2.6 - 85.3 | | 1.1 - 17.7 | 1.1 - 17.7 | 5.6 - 13.3 | |
| Clinical stage | | | | 0.1347 | | | | 0.2290 | | | | 0.9083 |
| T1-T2 | 228 (89.1) | 211 (90.2) | 17 (77.3) | | 195 (89.0) | 181 (90.0) | 14 (77.8) | | 33 (89.2) | 30 (90.9) | 3 (75.0) | |
| T3a | 28 (10.9) | 23 (9.8) | 5 (22.7) | | 24 (11.0) | 20 (10.0) | 4 (22.2) | | 4 (10.8) | 3 (9.1) | 1 (25.0) | |
| Global Gleason | | | | 0.4467 | | | | 0.7260 | | | | 0.2400 |
| 8 | 185 (63.8) | 173 (64.8) | 12 (52.2) | | 158 (64.2) | 147 (64.8) | 11 (57.9) | | 27 (61.4) | 26 (65.0) | 1 (25.0) | |
| 9 | 104 (35.9) | 93 (34.8) | 11 (47.8) | | 88 (35.8) | 80 (35.2) | 8 (42.1) | | 16 (36.4) | 13 (32.5) | 3 (75.0) | |
| 10 | 1 (0.3) | 1 (0.4) | 0 (0.0) | | 0 (0.0) | 0 (0.0) | 0 (0.0) | | 1 (2.3) | 1 (2.5) | 0 (0.0) | |
| Primary Gleason | | | | 0.2034 | | | | 0.3591 | | | | 0.4745 |
| 3 | 16 (5.5) | 14 (5.2) | 2 (8.7) | | 14 (5.7) | 12 (5.3) | 2 (10.5) | | 2 (4.5) | 2 (5.0) | 0 (0.0) | |
| 4 | 265 (91.4) | 246 (92.1) | 19 (82.6) | | 227 (92.3) | 211 (93.0) | 16 (84.2) | | 38 (86.4) | 35 (87.5) | 3 (75.0) | |
| 5 | 9 (3.1) | 7 (2.6) | 2 (8.7) | | 5 (2.0) | 4 (1.8) | 1 (5.3) | | 4 (9.1) | 3 (7.5) | 1 (25.0) | |
| Secondary Gleason | | | | 0.6625 | | | | 0.6223 | | | | 0.8570 |
| 3 | 1 (0.3) | 1 (0.4) | 0 (0.0) | | 0 (0.0) | 0 (0.0) | 0 (0.0) | | 1 (2.3) | 1 (2.5) | 0 (0.0) | |
| 4 | 175 (60.3) | 163 (61.0) | 12 (52.2) | | 149 (60.6) | 139 (61.2) | 10 (52.6) | | 26 (59.1) | 24 (60.0) | 2 (50.0) | |
| 5 | 114 (39.3) | 103 (38.6) | 11 (47.8) | | 97 (39.4) | 88 (38.8) | 9 (47.4) | | 17 (38.6) | 15 (37.5) | 2 (50.0) | |

**Supplementary Table 6** Descriptive analysis of the clinical features for the PCSS task on the *full dataset*, the *learning set*, and the *holdout set*. The $p$-values are computed using the Mann-Whitney U test [1] for continuous clinical features (age and psa) and the $\chi^2$ test [2] for categorical features (clinical stage, global Gleason score, primary Gleason score, and secondary Gleason score) using `scipy` [3] Python library.

| | Full dataset | | | | Learning set | | | | Holdout set | | | |
|---|---|---|---|---|---|---|---|---|---|---|---|---|
| Feature | All $n = 295$ 100.0 % | Negative $n = 284$ 96.3 % | Positive $n = 11$ 3.7 % | $p$-value | All $n = 250$ 100.0 % | Negative $n = 241$ 96.4 % | Positive $n = 9$ 3.6 % | $p$-value | All $n = 45$ 100.0 % | Negative $n = 43$ 95.6 % | Positive $n = 2$ 4.4 % | $p$-value |
| Age [years] | | | | 0.7197 | | | | 0.9719 | | | | 0.3774 |
| Mean (Median) | 65.4 (66.0) | 65.4 (66.0) | 64.3 (66.0) | | 65.6 (66.0) | 65.6 (66.0) | 65.2 (67.0) | | 64.4 (66.0) | 64.6 (66.0) | 60.0 (60.0) | |
| Min - Max | 48.0 - 80.0 | 48.0 - 80.0 | 48.0 - 77.0 | | 48.0 - 80.0 | 48.0 - 80.0 | 48.0 - 77.0 | | 50.0 - 76.0 | 50.0 - 76.0 | 54.0 - 66.0 | |
| PSA [ng/ml] | | | | 0.0494 | | | | 0.0879 | | | | 0.3082 |
| Mean (Median) | 11.0 (7.4) | 10.7 (7.2) | 17.3 (10.0) | | 11.5 (7.4) | 11.2 (7.3) | 18.9 (10.0) | | 8.0 (7.4) | 7.9 (7.2) | 10.4 (10.4) | |
| Min - Max | 1.1 - 155.3 | 1.1 - 155.3 | 2.6 - 85.3 | | 1.1 - 155.3 | 1.1 - 155.3 | 2.6 - 85.3 | | 1.1 - 17.7 | 1.1 - 17.7 | 7.4 - 13.3 | |
| Clinical stage | | | | 0.0367 | | | | 0.1413 | | | | 0.4932 |
| T1-T2 | 230 (88.1) | 223 (89.2) | 7 (63.6) | | 196 (87.9) | 190 (88.8) | 6 (66.7) | | 34 (89.5) | 33 (91.7) | 1 (50.0) | |
| T3a | 31 (11.9) | 27 (10.8) | 4 (36.4) | | 27 (12.1) | 24 (11.2) | 3 (33.3) | | 4 (10.5) | 3 (8.3) | 1 (50.0) | |
| Global Gleason | | | | 0.7873 | | | | 0.8541 | | | | 0.8960 |
| 8 | 188 (63.7) | 182 (64.1) | 6 (54.5) | | 160 (64.0) | 155 (64.3) | 5 (55.6) | | 28 (62.2) | 27 (62.8) | 1 (50.0) | |
| 9 | 106 (35.9) | 101 (35.6) | 5 (45.5) | | 90 (36.0) | 86 (35.7) | 4 (44.4) | | 16 (35.6) | 15 (34.9) | 1 (50.0) | |
| 10 | 1 (0.3) | 1 (0.4) | 0 (0.0) | | 0 (0.0) | 0 (0.0) | 0 (0.0) | | 1 (2.2) | 1 (2.3) | 0 (0.0) | |
| Primary Gleason | | | | 0.5308 | | | | 0.6906 | | | | 0.1983 |
| 3 | 16 (5.4) | 15 (5.3) | 1 (9.1) | | 14 (5.6) | 13 (5.4) | 1 (11.1) | | 2 (4.4) | 2 (4.7) | 0 (0.0) | |
| 4 | 268 (90.8) | 259 (91.2) | 9 (81.8) | | 230 (92.0) | 222 (92.1) | 8 (88.9) | | 38 (84.4) | 37 (86.0) | 1 (50.0) | |
| 5 | 11 (3.7) | 10 (3.5) | 1 (9.1) | | 6 (2.4) | 6 (2.5) | 0 (0.0) | | 5 (11.1) | 4 (9.3) | 1 (50.0) | |
| Secondary Gleason | | | | 0.8764 | | | | 0.4991 | | | | 0.4654 |
| 3 | 2 (0.7) | 2 (0.7) | 0 (0.0) | | 0 (0.0) | 0 (0.0) | 0 (0.0) | | 2 (4.4) | 2 (4.7) | 0 (0.0) | |
| 4 | 178 (60.3) | 172 (60.6) | 6 (54.5) | | 152 (60.8) | 148 (61.4) | 4 (44.4) | | 26 (57.8) | 24 (55.8) | 2 (100.0) | |
| 5 | 115 (39.0) | 110 (38.7) | 5 (45.5) | | 98 (39.2) | 93 (38.6) | 5 (55.6) | | 17 (37.8) | 17 (39.5) | 0 (0.0) | |

#### 1.2 Descriptive analysis of outcomes

**Supplementary Table 7** Survival time analysis for each task on the *full dataset*, the *learning set*, and the *holdout set*, with all durations measured in months from the date of radical prostatectomy (RP). Survival time, also known as failure time, refers to the duration of time between RP and the particular event (task) of interest. For instance, the median survival time is obtained by calculating the median of the observed times of patients who did experience failure.

| | Full dataset | | | Learning set | | | Holdout set | | |
|---|---|---|---|---|---|---|---|---|---|
| Task | Mean (Median) | Min-Max | Std | Mean (Median) | Min-Max | Std | Mean (Median) | Min-Max | Std |
| BCR-FS | 20.39 (9.05) | 0.89 - 106.02 | 25.18 | 19.52 (7.59) | 0.89 - 101.89 | 24.87 | 24.71 (14.88) | 1.94 - 106.02 | 26.72 |
| MFS | 35.19 (26.68) | 0.03 - 101.85 | 27.49 | 34.83 (26.68) | 0.03 - 101.85 | 26.90 | 37.05 (23.23) | 5.32 - 92.06 | 33.13 |
| dADT-FS | 31.06 (18.69) | 2.50 - 103.70 | 28.51 | 31.22 (19.30) | 2.50 - 103.70 | 27.99 | 29.31 (11.19) | 3.84 - 95.74 | 36.80 |
| CRPC-FS | 36.93 (34.79) | 11.27 - 87.03 | 21.77 | 39.06 (35.68) | 12.39 - 87.03 | 21.46 | 26.79 (17.17) | 11.27 - 61.57 | 23.35 |
| PCSS | 60.51 (57.30) | 23.43 - 102.57 | 26.40 | 61.09 (57.29) | 23.43 - 102.57 | 28.96 | 57.92 (57.92) | 46.85 - 68.99 | 15.66 |

**Supplementary Table 8** Median follow-up time in the *full dataset*, the *learning set*, and the *holdout set*, with all durations measured in months from the date of radical prostatectomy (RP). Three methods are used to calculate the median follow-up time [4–6]: considering all patients in the study, regardless of censoring or failure ($T_{\text{obs}}$); using only patients who did not experience failure ($T_{\text{cens}}$); or generating a reverse Kaplan-Meier curve, where censoring and failure are swapped, and using the point at 50% of this curve as the median follow-up ($T_{\text{R-KM}}$).

| | Full dataset | | | Learning set | | | Holdout set | | |
|---|---|---|---|---|---|---|---|---|---|
| Task | $T_{\text{obs}}$ | $T_{\text{cens}}$ | $T_{\text{R-KM}}$ | $T_{\text{obs}}$ | $T_{\text{cens}}$ | $T_{\text{R-KM}}$ | $T_{\text{obs}}$ | $T_{\text{cens}}$ | $T_{\text{R-KM}}$ |
| BCR-FS | 20.60 | 38.11 | 66.86 | 20.60 | 41.05 | 66.86 | 20.42 | 24.15 | 55.29 |
| MFS | 55.33 | 84.80 | 87.29 | 56.57 | 86.64 | 88.71 | 47.28 | 66.53 | 82.83 |
| dADT-FS | 49.31 | 60.29 | 73.49 | 49.53 | 66.86 | 75.96 | 43.70 | 54.80 | 55.29 |
| CRPC-FS | 60.39 | 66.86 | 69.42 | 62.72 | 69.42 | 70.74 | 54.80 | 55.05 | 55.29 |
| PCSS | 68.24 | 69.32 | 69.85 | 69.73 | 70.74 | 71.92 | 56.18 | 56.18 | 60.06 |

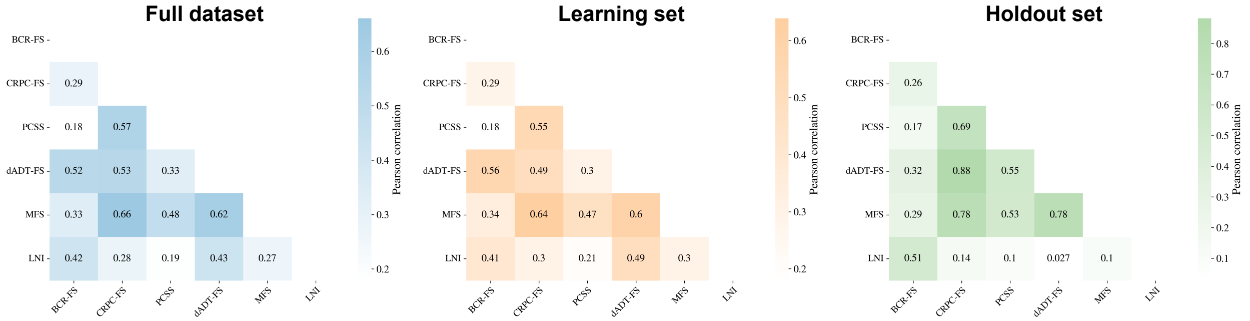


**Supplementary Fig. 1** Pearson correlation between each pair of tasks for the *full dataset*, the *learning set*, and the *holdout set*. The distributions of class labels and event indicators are respectively used to compute the correlation of classification and survival tasks.

##### 1.2.1 Biochemical recurrence-free survival (BCR-FS)

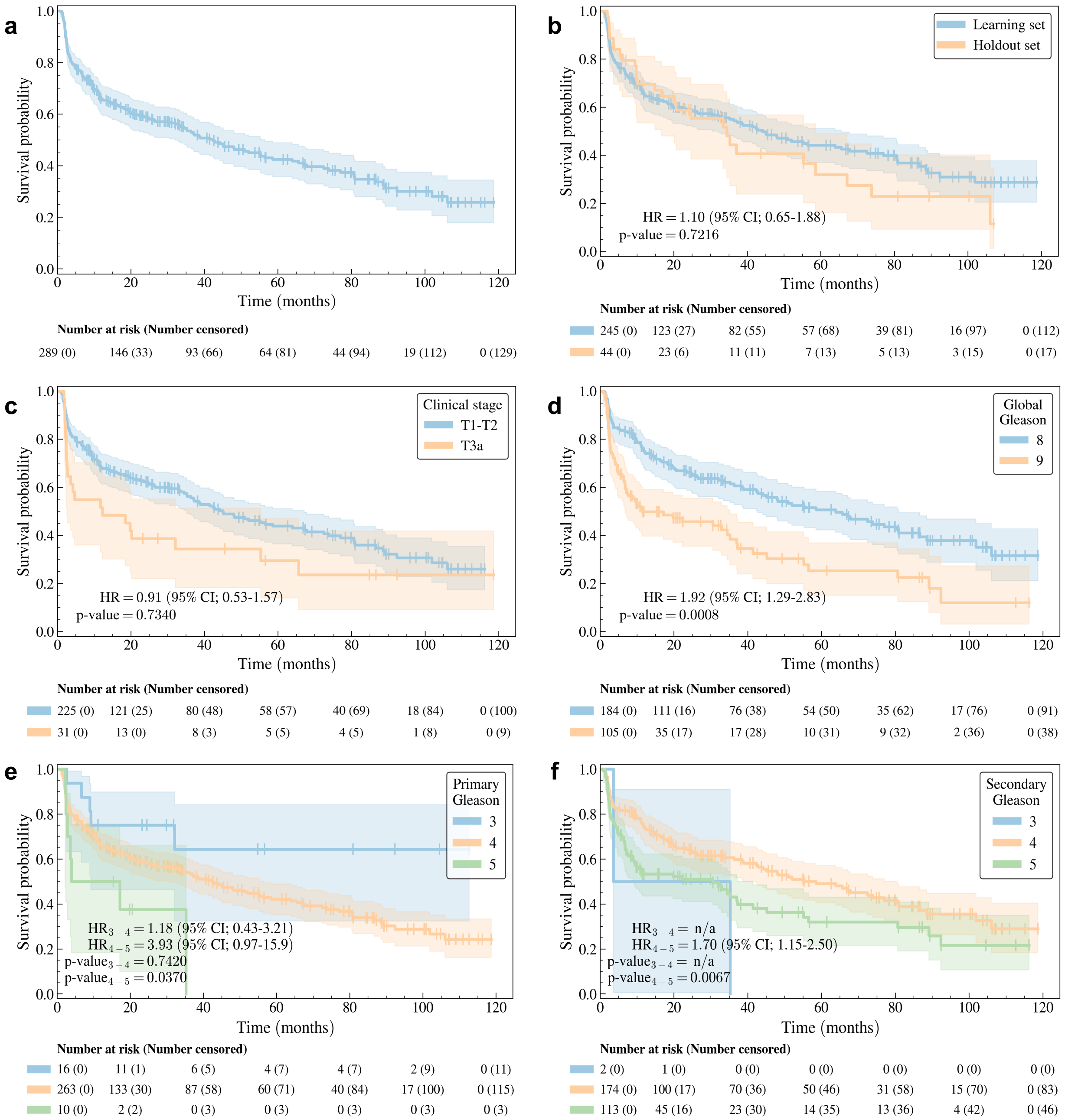


**Supplementary Fig. 2** Kaplan-Meier curve [7] of the *full dataset* for the BCR-FS task using (a) no stratification and stratification based on (b) datasets, (c) clinical stage, (d) global Gleason score, (e) primary Gleason score, and (f) secondary Gleason score. The 95% confidence interval (shade) of the Kaplan-Meier curve (line) is estimated using the log hazard [8]. The *p*-value is computed using a log-rank test [9, 10], which also provides statistics to calculate the hazard ratio (HR) and its 95% confidence interval (95% CI) [11]. The `scikit-survival` [12] Python Library is used to generate the Kaplan-Meier curves and perform the tests.

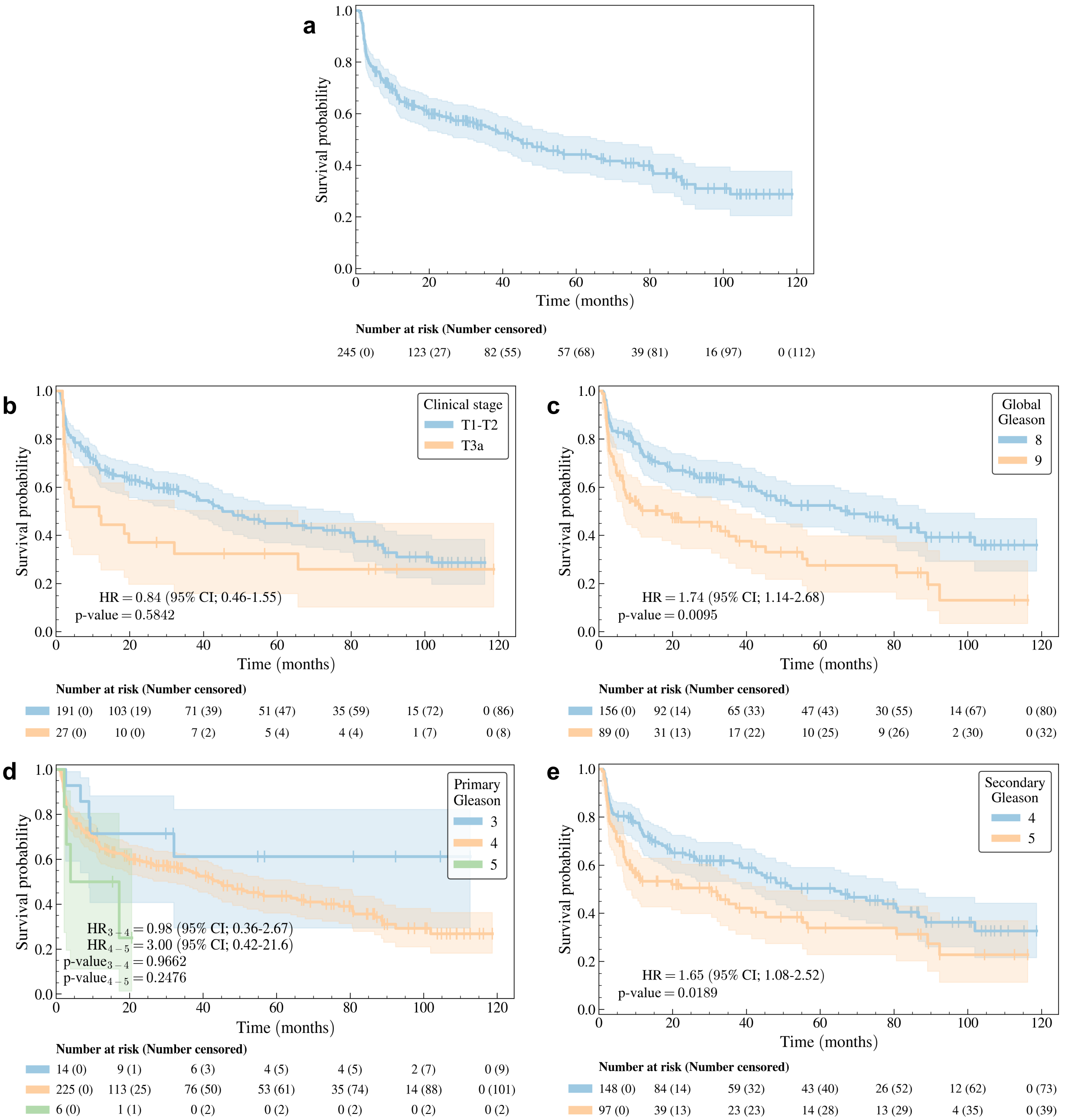


**Supplementary Fig. 3** Kaplan-Meier curve [7] of the *learning set* for the BCR-FS task using (a) no stratification and stratification based on (b) clinical stage, (c) global Gleason score, (d) primary Gleason score, and (e) secondary Gleason score. The 95% confidence interval (shade) of the Kaplan-Meier curve (line) is estimated using the log hazard [8]. The $p$-value is computed using a log-rank test [9, 10], which also provides statistics to calculate the hazard ratio (HR) and its 95% confidence interval (95% CI) [11]. The `scikit-survival` [12] Python Library is used to generate the Kaplan-Meier curves and perform the tests.

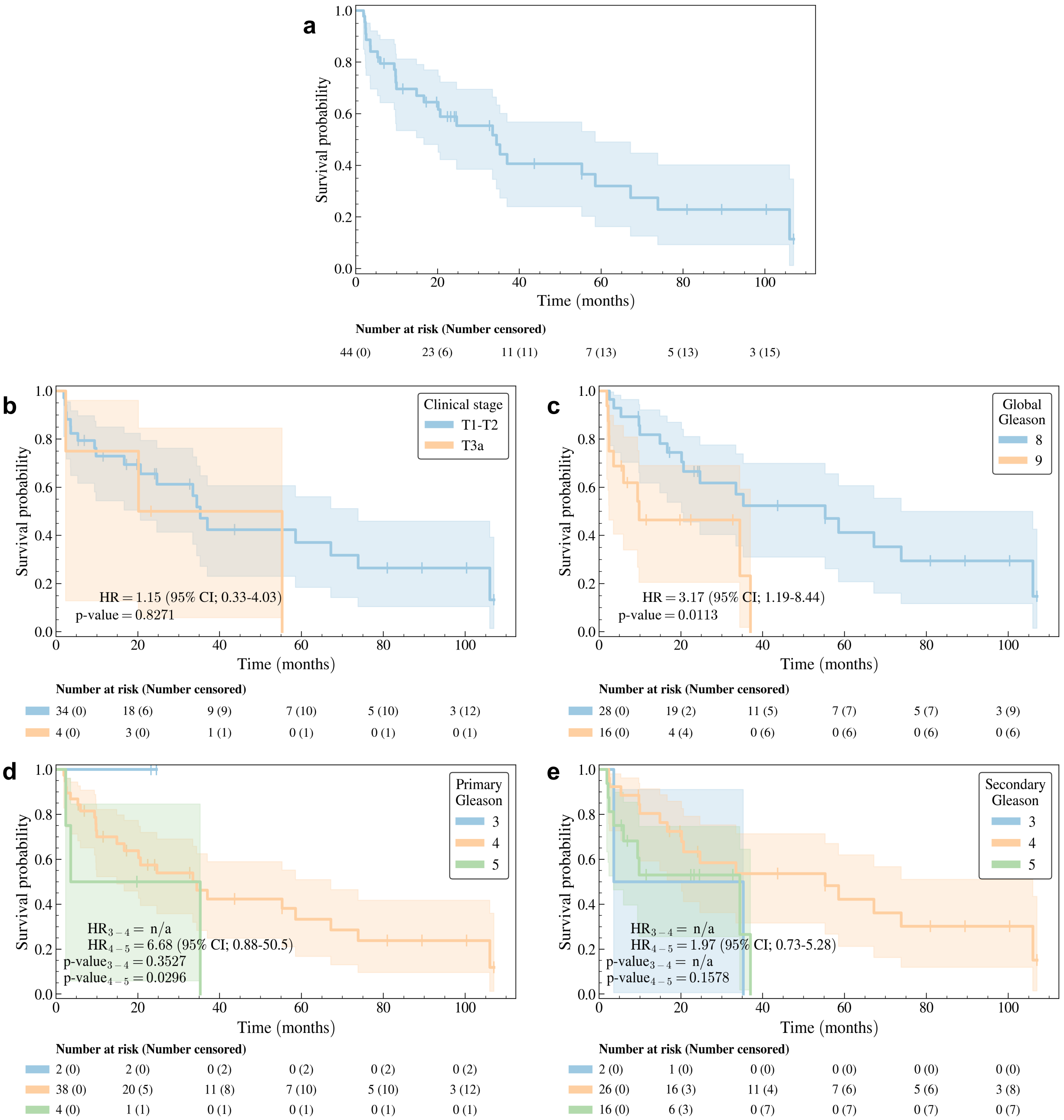


**Supplementary Fig. 4** Kaplan-Meier curve [7] of the *holdout set* for the BCR-FS task using (a) no stratification and stratification based on (b) clinical stage, (c) global Gleason score, (d) primary Gleason score, and (e) secondary Gleason score. The 95% confidence interval (shade) of the Kaplan-Meier curve (line) is estimated using the log hazard [8]. The $p$-value is computed using a log-rank test [9, 10], which also provides statistics to calculate the hazard ratio (HR) and its 95% confidence interval (95% CI) [11]. The `scikit-survival` [12] Python Library is used to generate the Kaplan-Meier curves and perform the tests.

##### 1.2.2 Metastasis-free survival (MFS)

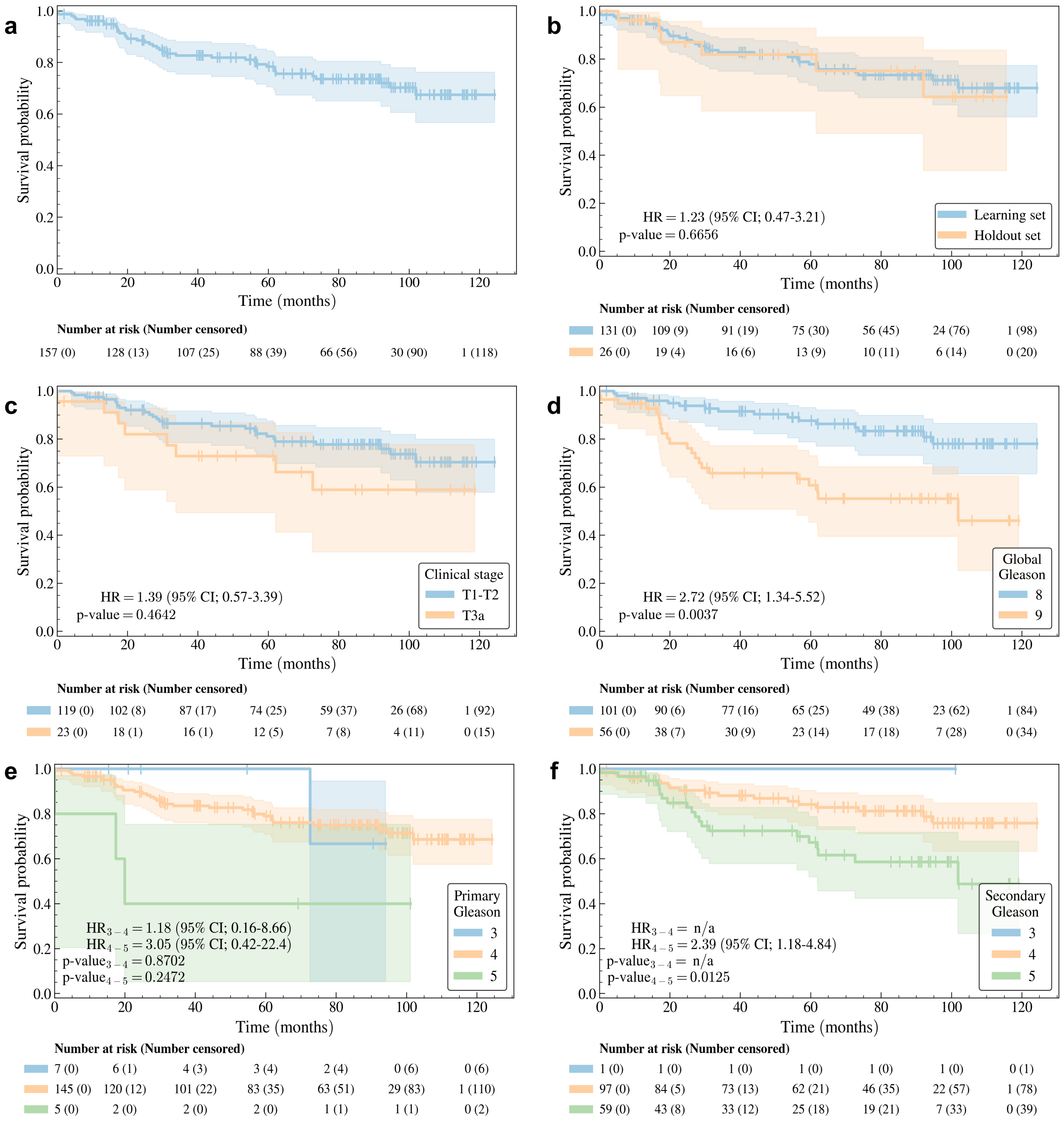


**Supplementary Fig. 5** Kaplan-Meier curve [7] of the *full dataset* for the MFS task using (a) no stratification and stratification based on (b) datasets, (c) clinical stage, (d) global Gleason score, (e) primary Gleason score, and (f) secondary Gleason score. The 95% confidence interval (shade) of the Kaplan-Meier curve (line) is estimated using the log hazard [8]. The $p$-value is computed using a log-rank test [9, 10], which also provides statistics to calculate the hazard ratio (HR) and its 95% confidence interval (95% CI) [11]. The `scikit-survival` [12] Python Library is used to generate the Kaplan-Meier curves and perform the tests.

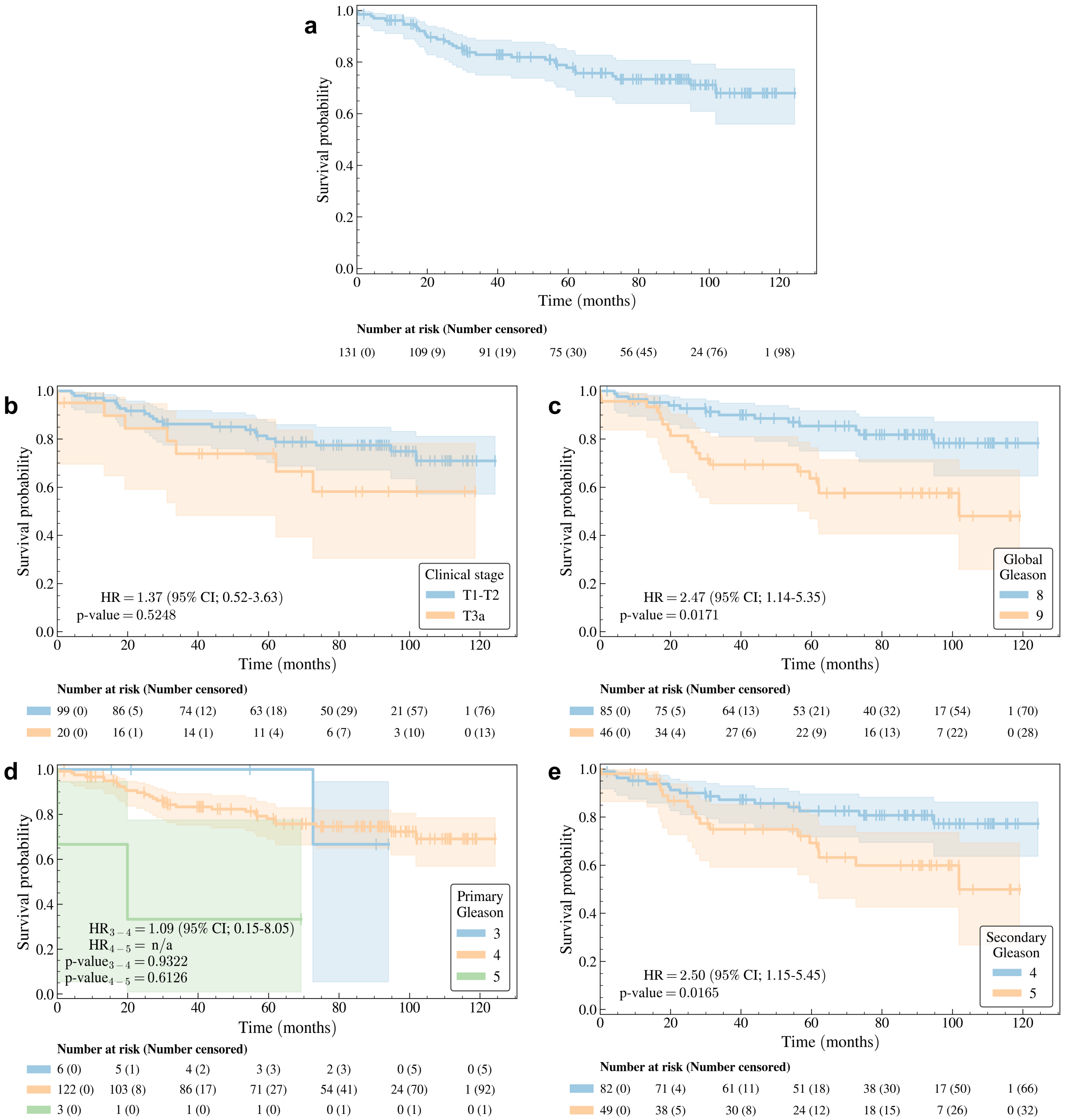


**Supplementary Fig. 6** Kaplan-Meier curve [7] of the *learning set* for the MFS task using (a) no stratification and stratification based on (b) clinical stage, (c) global Gleason score, (d) primary Gleason score, and (e) secondary Gleason score. The 95% confidence interval (shade) of the Kaplan-Meier curve (line) is estimated using the log hazard [8]. The $p$-value is computed using a log-rank test [9, 10], which also provides statistics to calculate the hazard ratio (HR) and its 95% confidence interval (95% CI) [11]. The `scikit-survival` [12] Python Library is used to generate the Kaplan-Meier curves and perform the tests.

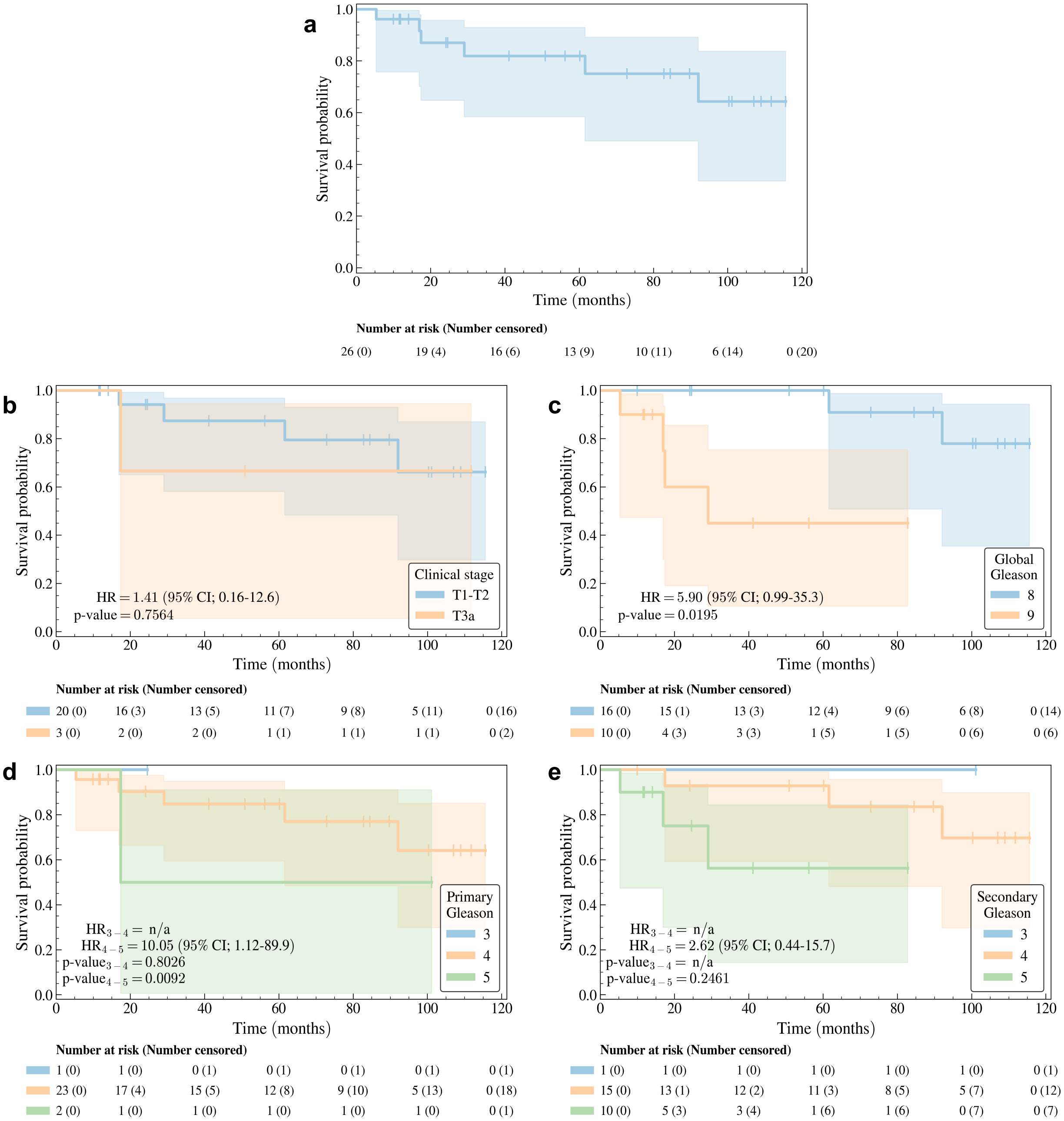


**Supplementary Fig. 7** Kaplan-Meier curve [7] of the *holdout set* for the MFS task using (a) no stratification and stratification based on (b) clinical stage, (c) global Gleason score, (d) primary Gleason score, and (e) secondary Gleason score. The 95% confidence interval (shade) of the Kaplan-Meier curve (line) is estimated using the log hazard [8]. The $p$-value is computed using a log-rank test [9, 10], which also provides statistics to calculate the hazard ratio (HR) and its 95% confidence interval (95% CI) [11]. The `scikit-survival` [12] Python Library is used to generate the Kaplan-Meier curves and perform the tests.

##### 1.2.3 Definitive androgen deprivation therapy-free (dADT-FS)

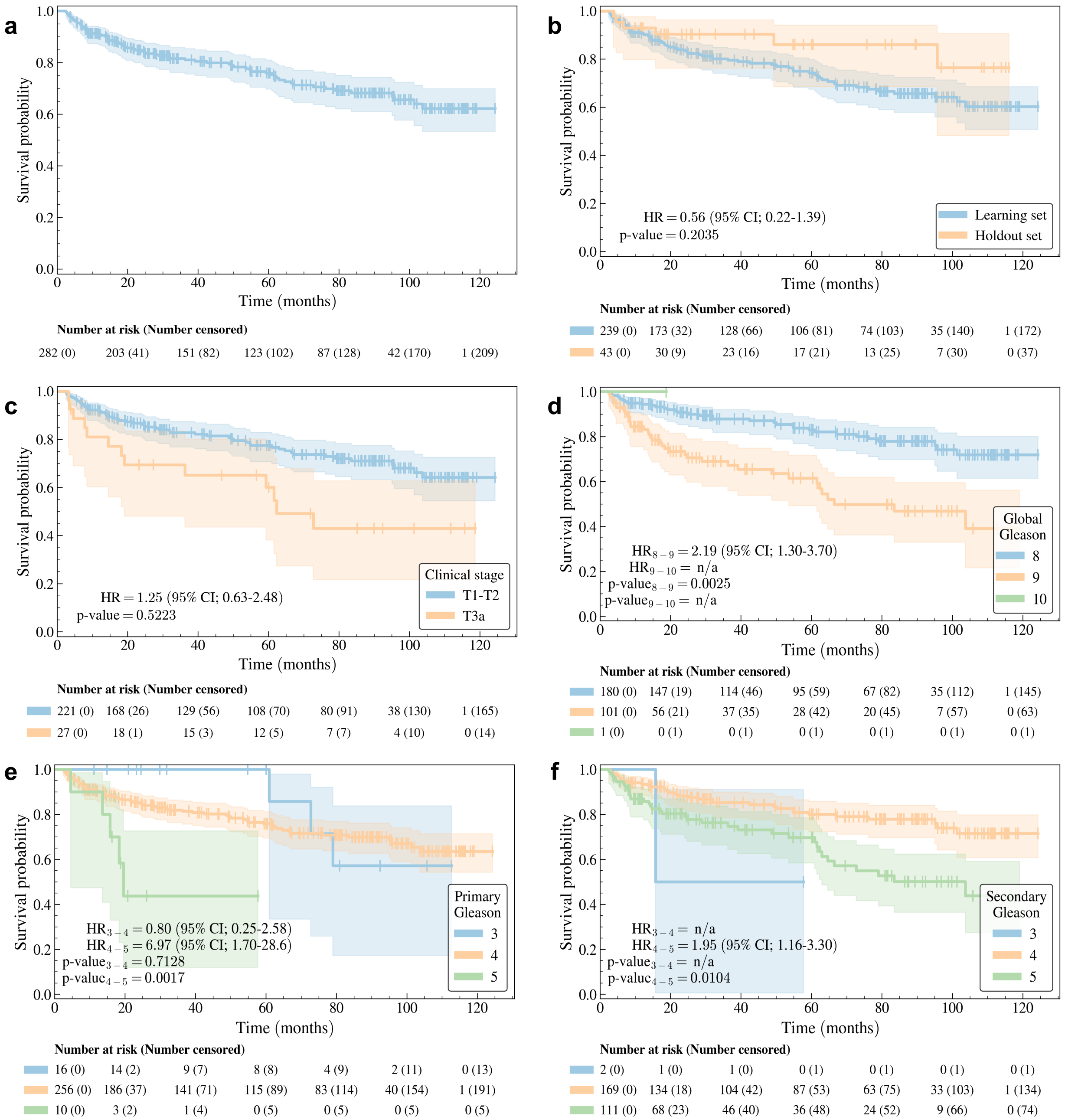


**Supplementary Fig. 8** Kaplan-Meier curve [7] of the *full dataset* for the dADT-FS task using (a) no stratification and stratification based on (b) datasets, (c) clinical stage, (d) global Gleason score, (e) primary Gleason score, and (f) secondary Gleason score. The 95% confidence interval (shade) of the Kaplan-Meier curve (line) is estimated using the log hazard [8]. The *p*-value is computed using a log-rank test [9, 10], which also provides statistics to calculate the hazard ratio (HR) and its 95% confidence interval (95% CI) [11]. The `scikit-survival` [12] Python Library is used to generate the Kaplan-Meier curves and perform the tests.

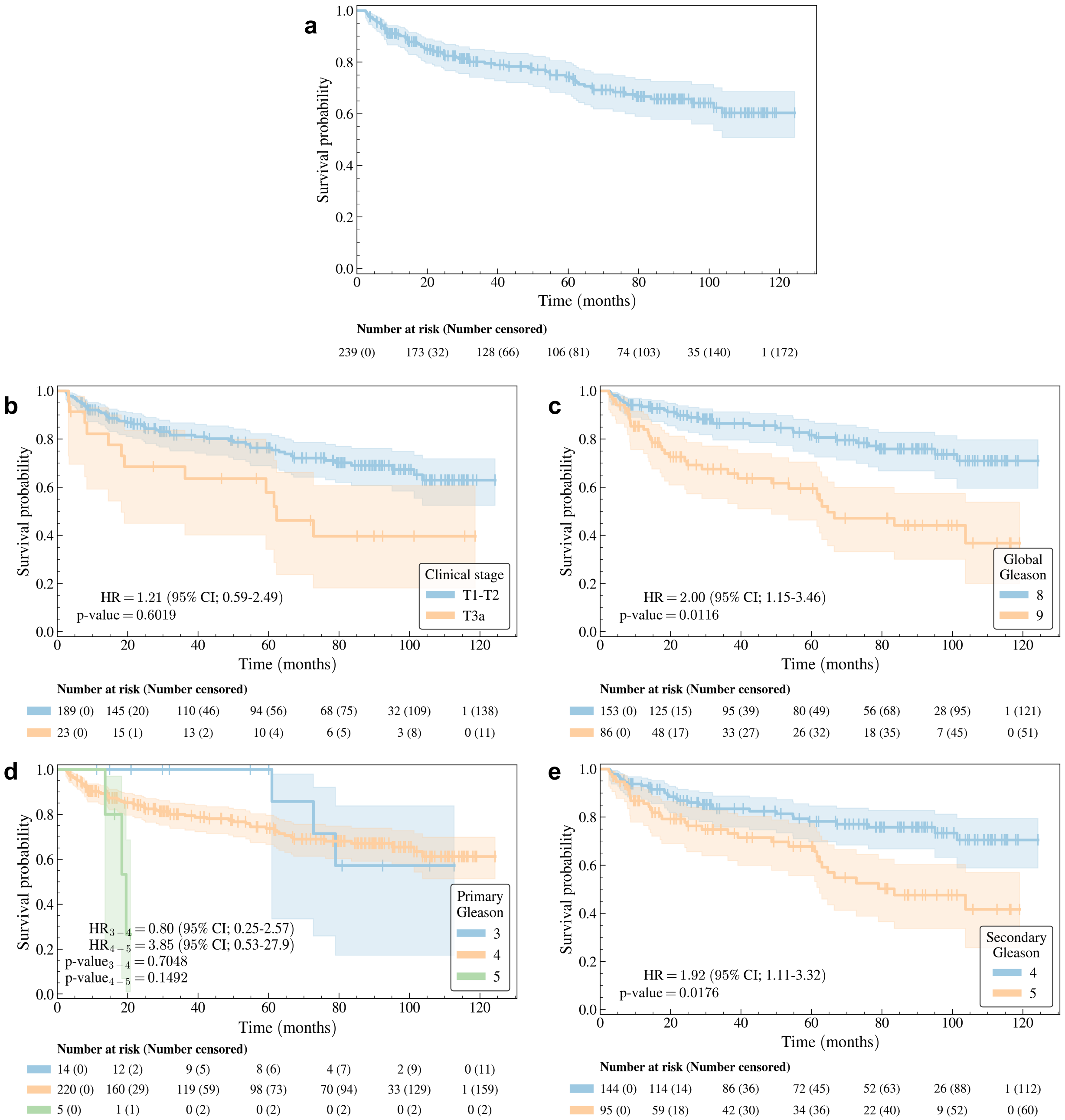


**Supplementary Fig. 9** Kaplan-Meier curve [7] of the *learning set* for the dADT-FS task using (a) no stratification and stratification based on (b) clinical stage, (c) global Gleason score, (d) primary Gleason score, and (e) secondary Gleason score. The 95% confidence interval (shade) of the Kaplan-Meier curve (line) is estimated using the log hazard [8]. The $p$-value is computed using a log-rank test [9, 10], which also provides statistics to calculate the hazard ratio (HR) and its 95% confidence interval (95% CI) [11]. The `scikit-survival` [12] Python Library is used to generate the Kaplan-Meier curves and perform the tests.

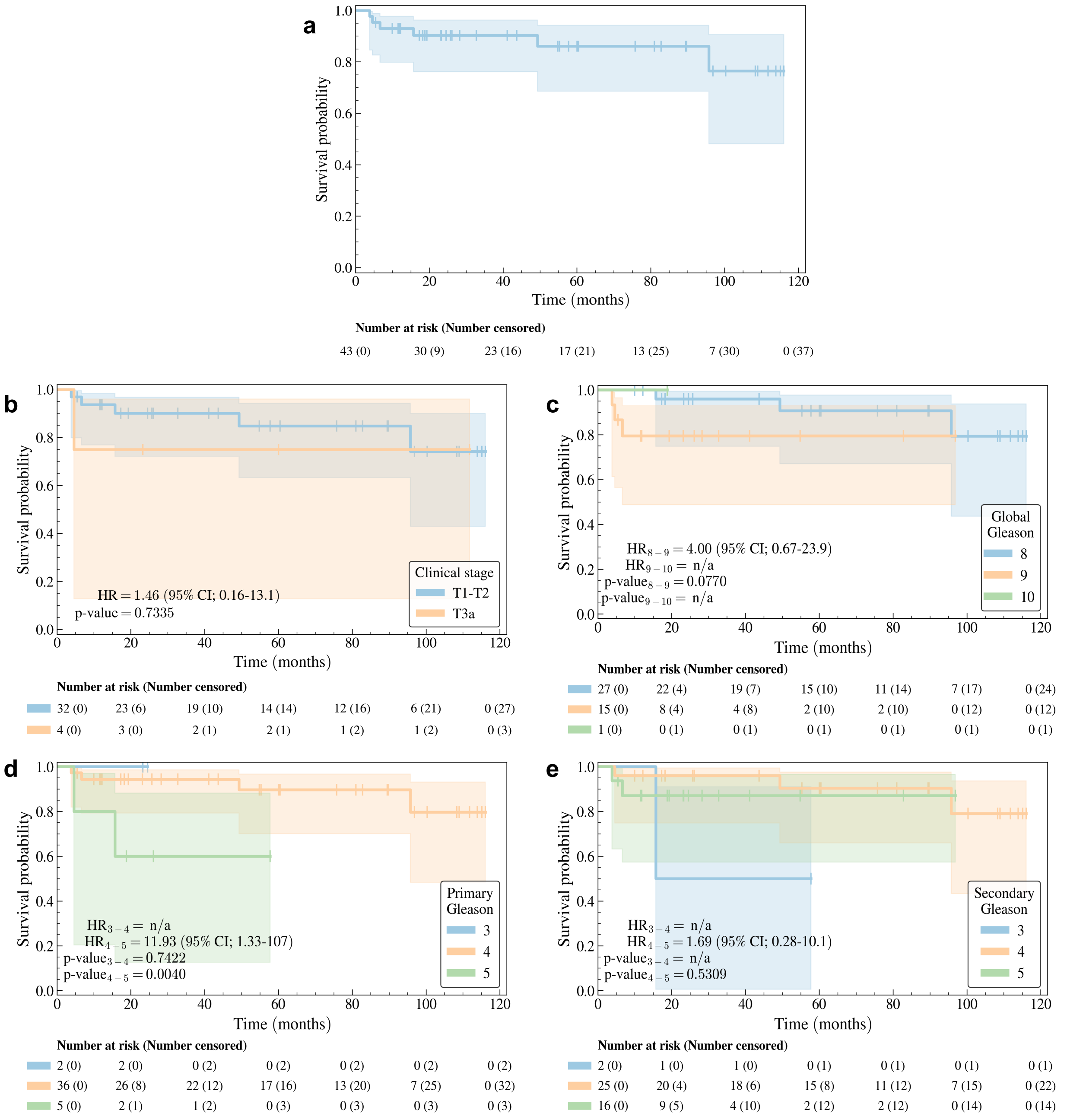


**Supplementary Fig. 10** Kaplan-Meier curve [7] of the *holdout set* for the dADT-FS task using (a) no stratification and stratification based on (b) clinical stage, (c) global Gleason score, (d) primary Gleason score, and (e) secondary Gleason score. The 95% confidence interval (shade) of the Kaplan-Meier curve (line) is estimated using the log hazard [8]. The $p$-value is computed using a log-rank test [9, 10], which also provides statistics to calculate the hazard ratio (HR) and its 95% confidence interval (95% CI) [11]. The `scikit-survival` [12] Python Library is used to generate the Kaplan-Meier curves and perform the tests.

##### 1.2.4 Castration-resistant prostate cancer free survival (CRPC-FS)

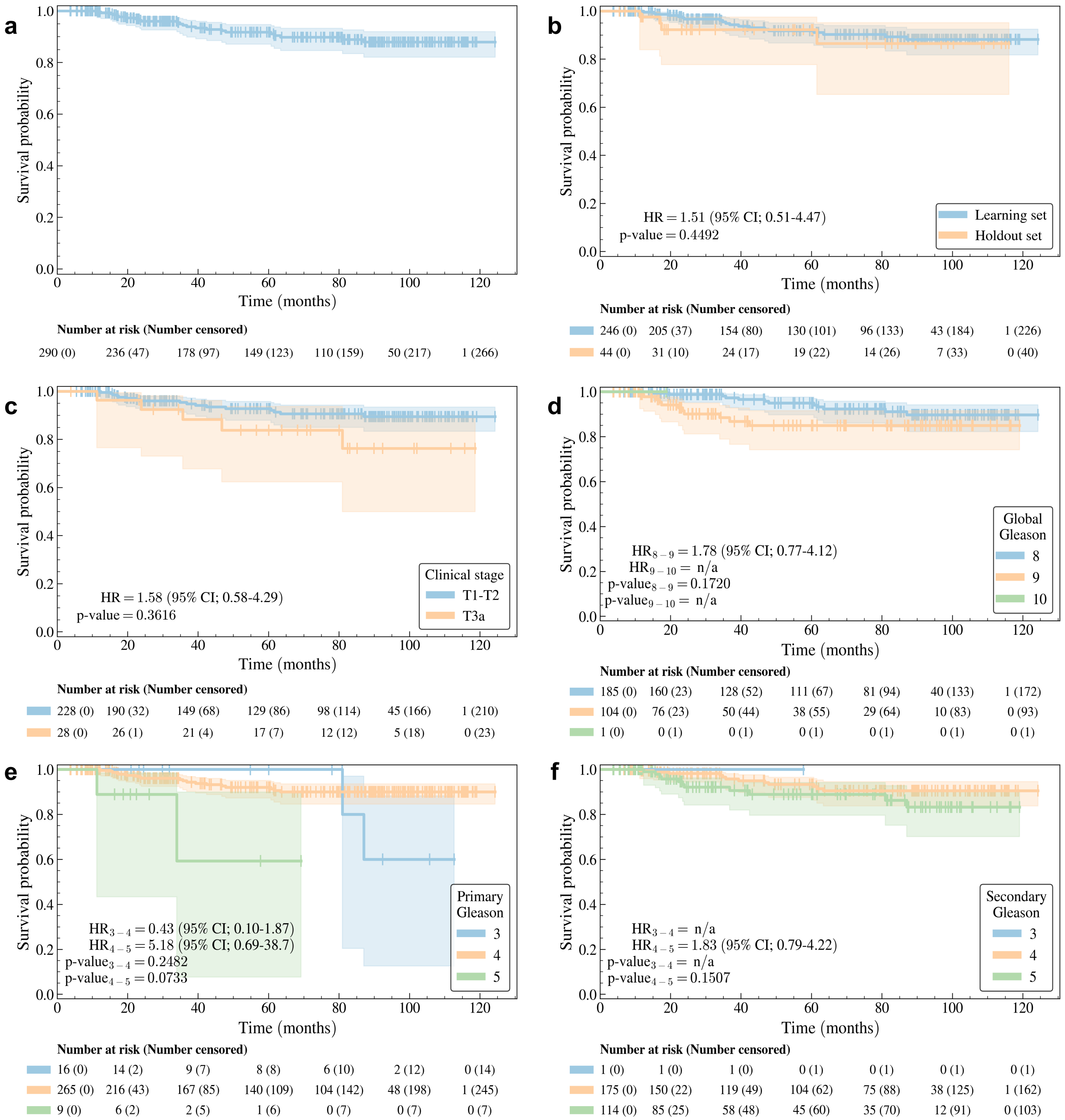


**Supplementary Fig. 11** Kaplan-Meier curve [7] of the *full dataset* for the CRPC-FS task using (a) no stratification and stratification based on (b) datasets, (c) clinical stage, (d) global Gleason score, (e) primary Gleason score, and (f) secondary Gleason score. The 95% confidence interval (shade) of the Kaplan-Meier curve (line) is estimated using the log hazard [8]. The $p$-value is computed using a log-rank test [9, 10], which also provides statistics to calculate the hazard ratio (HR) and its 95% confidence interval (95% CI) [11]. The `scikit-survival` [12] Python Library is used to generate the Kaplan-Meier curves and perform the tests.

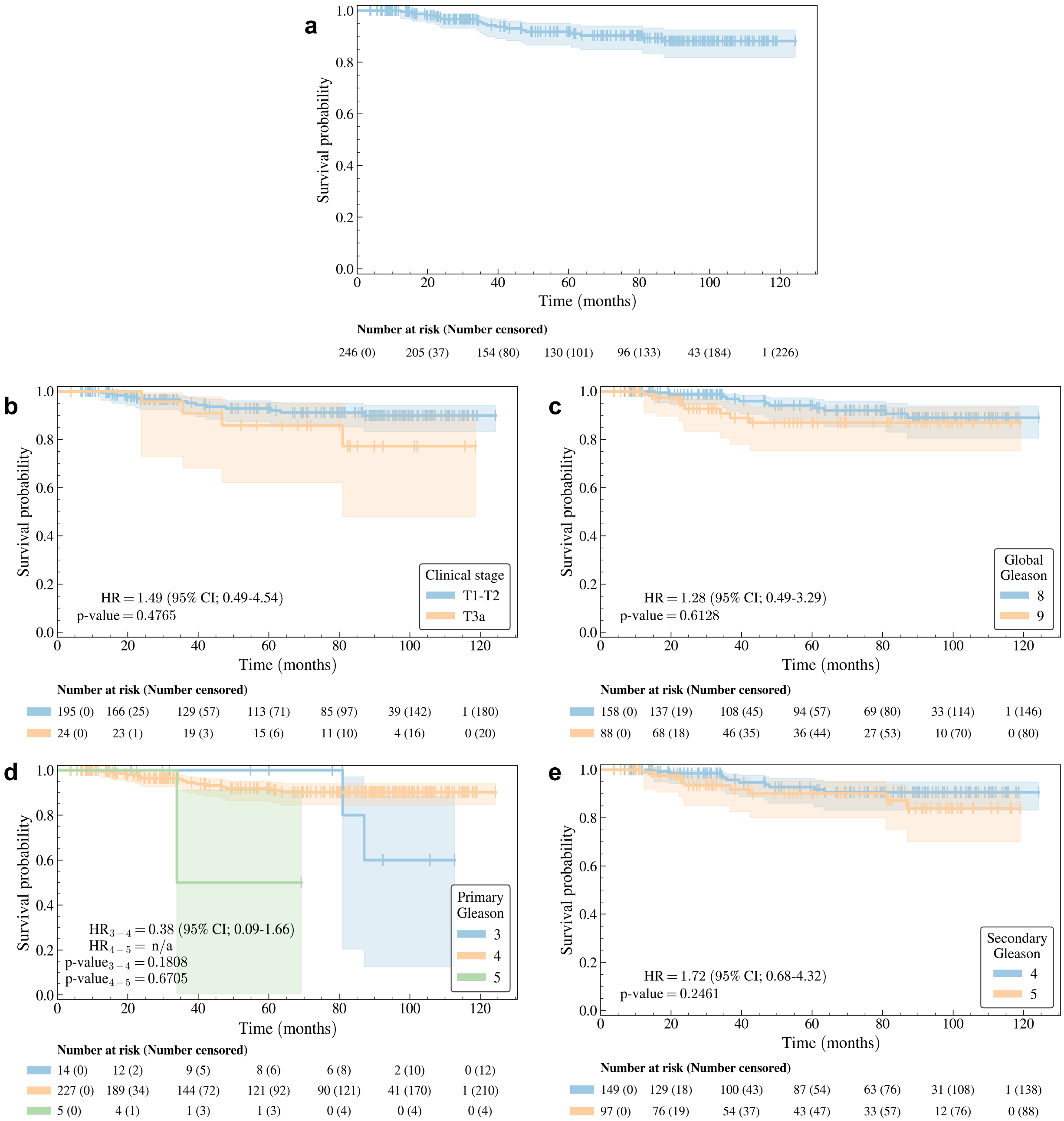


**Supplementary Fig. 12** Kaplan-Meier curve [7] of the *learning set* for the CRPC-FS task using (a) no stratification and stratification based on (b) clinical stage, (c) global Gleason score, (d) primary Gleason score, and (e) secondary Gleason score. The 95% confidence interval (shade) of the Kaplan-Meier curve (line) is estimated using the log hazard [8]. The $p$-value is computed using a log-rank test [9, 10], which also provides statistics to calculate the hazard ratio (HR) and its 95% confidence interval (95% CI) [11]. The `scikit-survival` [12] Python Library is used to generate the Kaplan-Meier curves and perform the tests.

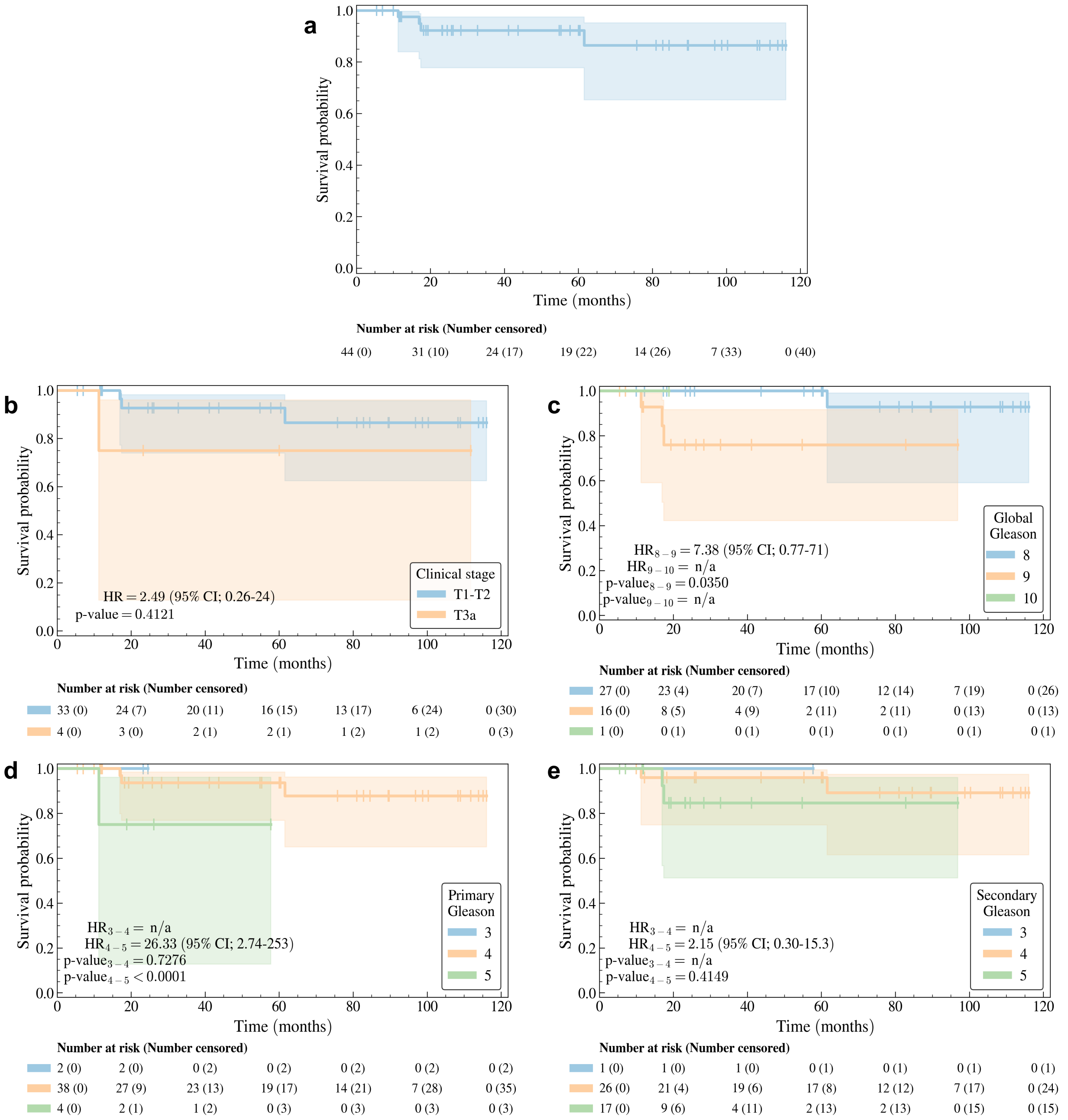


**Supplementary Fig. 13** Kaplan-Meier curve [7] of the *holdout set* for the CRPC-FS task using (a) no stratification and stratification based on (b) clinical stage, (c) global Gleason score, (d) primary Gleason score, and (e) secondary Gleason score. The 95% confidence interval (shade) of the Kaplan-Meier curve (line) is estimated using the log hazard [8]. The $p$-value is computed using a log-rank test [9, 10], which also provides statistics to calculate the hazard ratio (HR) and its 95% confidence interval (95% CI) [11]. The `scikit-survival` [12] Python Library is used to generate the Kaplan-Meier curves and perform the tests.

##### 1.2.5 Prostate cancer-specific survival (PCSS)

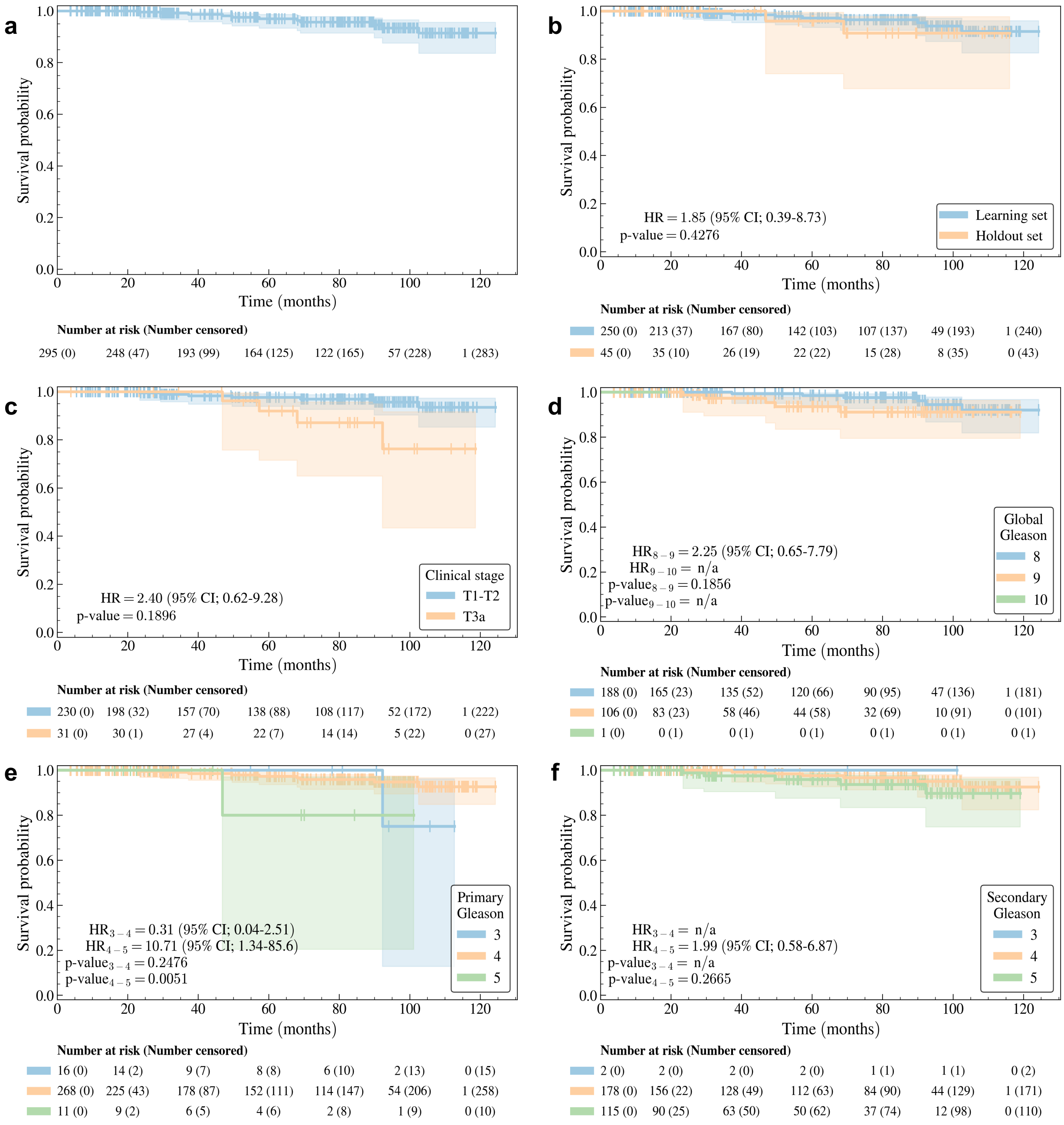


**Supplementary Fig. 14** Kaplan-Meier curve [7] of the *full dataset* for the PCSS task using (a) no stratification and stratification based on (b) datasets, (c) clinical stage, (d) global Gleason score, (e) primary Gleason score, and (f) secondary Gleason score. The 95% confidence interval (shade) of the Kaplan-Meier curve (line) is estimated using the log hazard [8]. The *p*-value is computed using a log-rank test [9, 10], which also provides statistics to calculate the hazard ratio (HR) and its 95% confidence interval (95% CI) [11]. The `scikit-survival` [12] Python Library is used to generate the Kaplan-Meier curves and perform the tests.

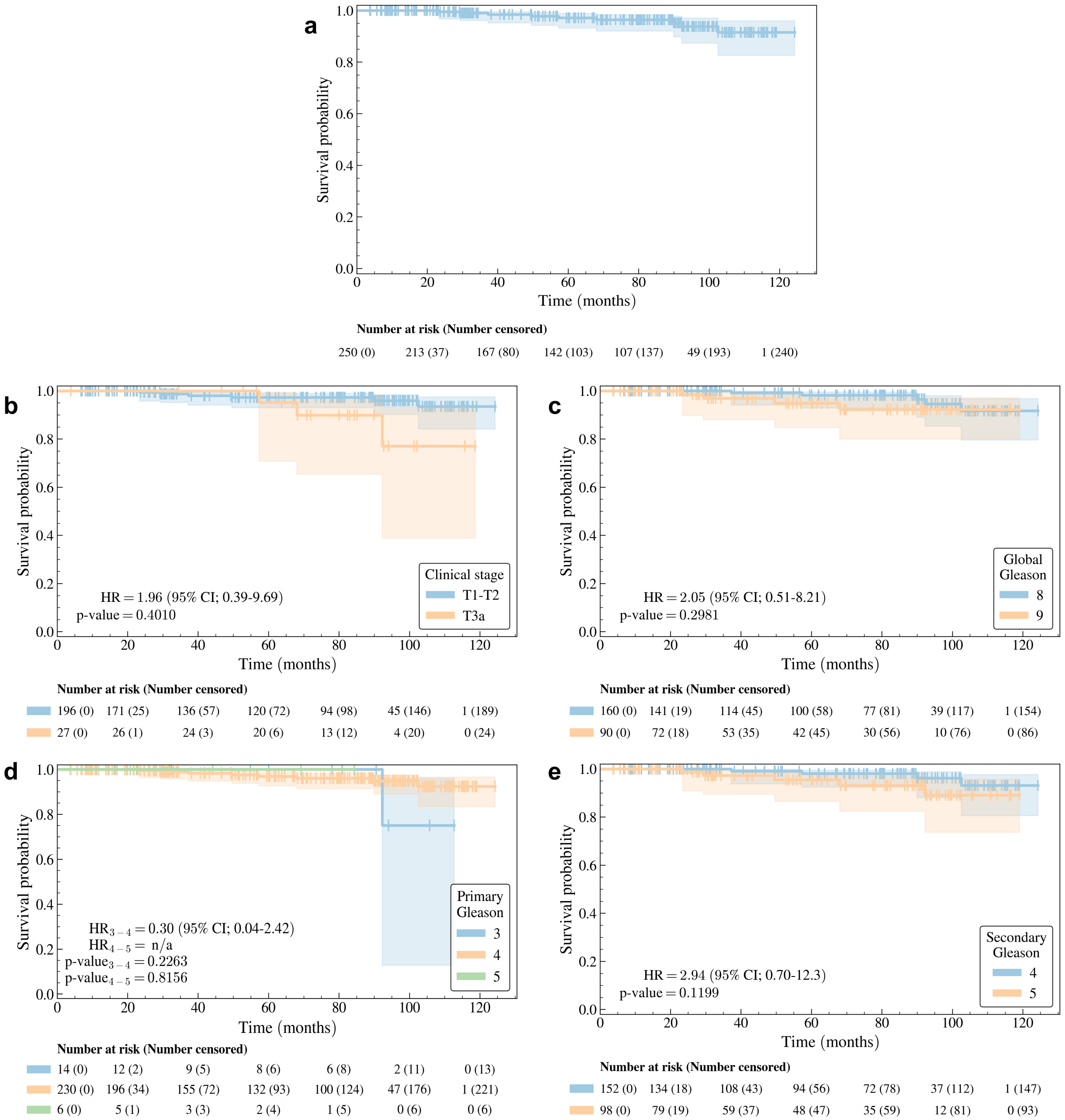


**Supplementary Fig. 15** Kaplan-Meier curve [7] of the *learning set* for the PCSS task using (a) no stratification and stratification based on (b) clinical stage, (c) global Gleason score, (d) primary Gleason score, and (e) secondary Gleason score. The 95% confidence interval (shade) of the Kaplan-Meier curve (line) is estimated using the log hazard [8]. The $p$-value is computed using a log-rank test [9, 10], which also provides statistics to calculate the hazard ratio (HR) and its 95% confidence interval (95% CI) [11]. The `scikit-survival` [12] Python Library is used to generate the Kaplan-Meier curves and perform the tests.

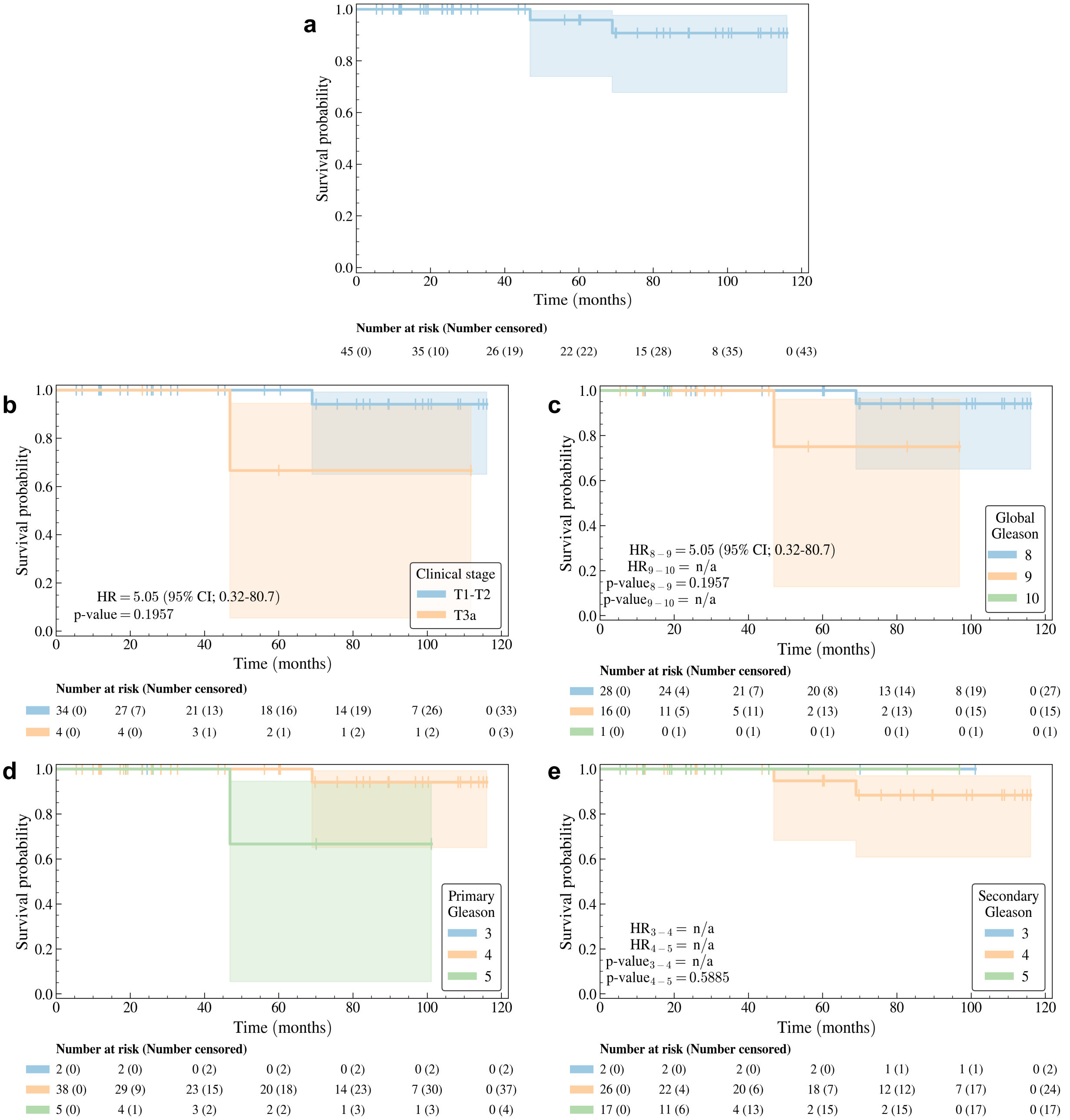


**Supplementary Fig. 16** Kaplan-Meier curve [7] of the *holdout set* for the PCSS task using (a) no stratification and stratification based on (b) clinical stage, (c) global Gleason score, (d) primary Gleason score, and (e) secondary Gleason score. The 95% confidence interval (shade) of the Kaplan-Meier curve (line) is estimated using the log hazard [8]. The $p$-value is computed using a log-rank test [9, 10], which also provides statistics to calculate the hazard ratio (HR) and its 95% confidence interval (95% CI) [11]. The `scikit-survival` [12] Python Library is used to generate the Kaplan-Meier curves and perform the tests.

#### 1.3 Visual comparison between the *learning set* and the *holdout set*

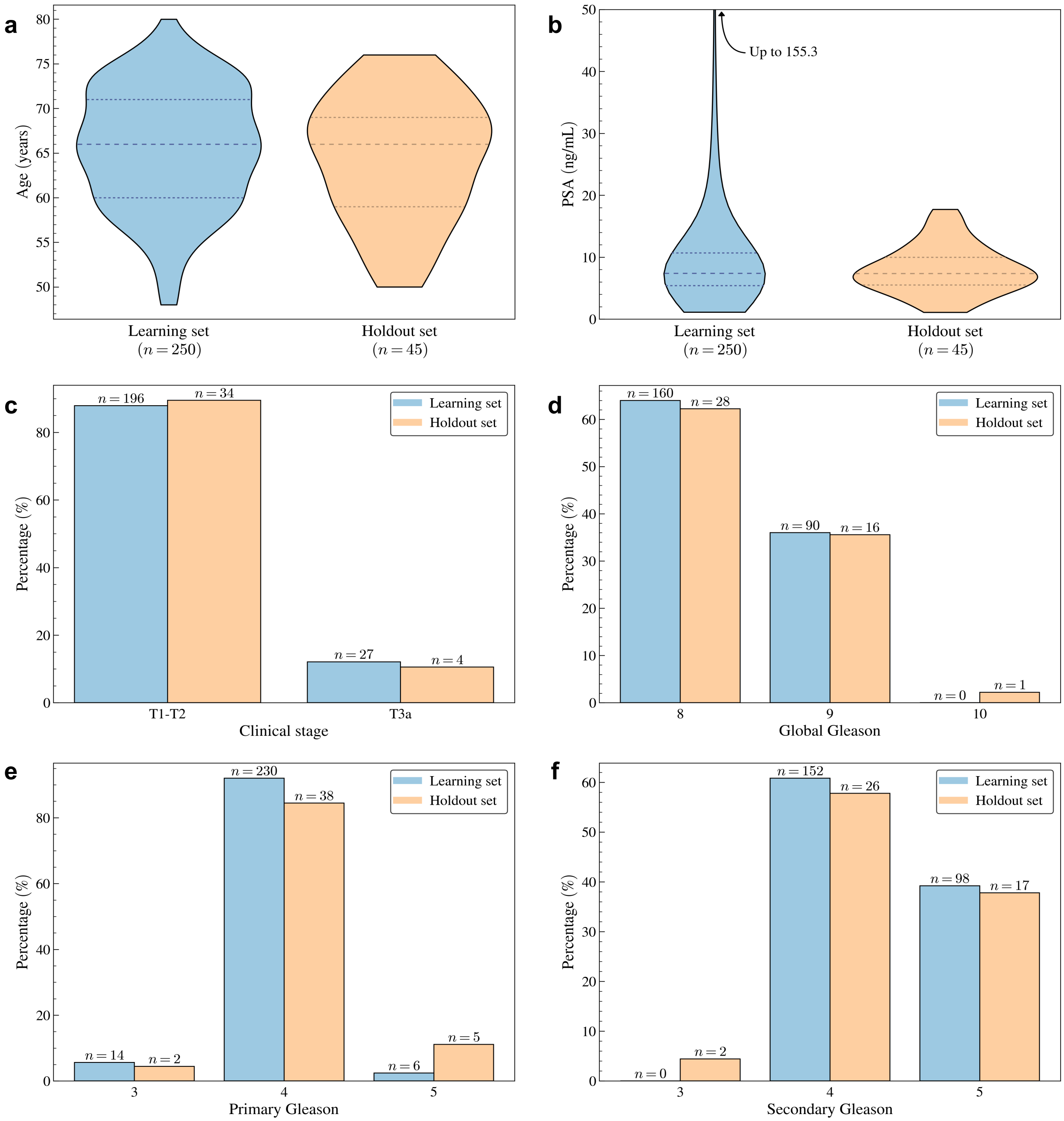


**Supplementary Fig. 17** Comparison of the distribution of (a) age, (b) prostate-specific antigen (PSA), (c) clinical stage, (d) global Gleason score, (e) primary Gleason score, and (f) secondary Gleason score between the *learning set* and the *holdout set*. Note that there is 1 patient with a missing PSA and 34 patients with missing clinical stage in the *full dataset*.

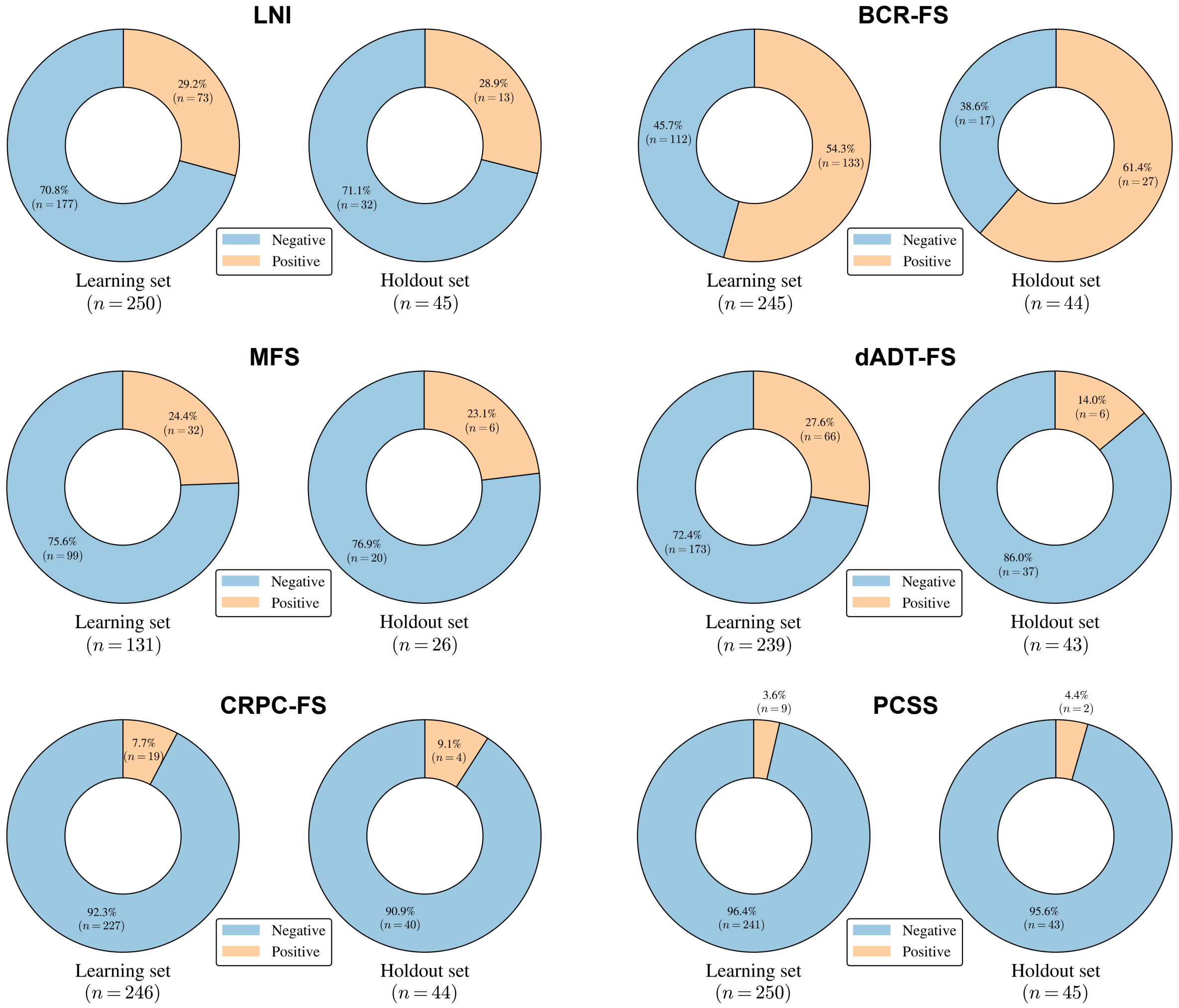


**Supplementary Fig. 18** Comparison of the distribution of class labels and event indicators between the *learning set* and the *holdout set* for the classification and survival tasks, respectively.

##### 1.4 Statistical analysis between the *learning set* and the *holdout set*

**Supplementary Table 9** Statistical analysis comparing the distribution of each clinical feature between the *learning set* and the *holdout set* using the Mann-Whitney U test [1] and the $\chi^2$ test [2].

| Feature | Test | $p$-value |
|---|---|---|
| Age | Mann–Whitney U | $4 \times 10^{-1}$ |
| PSA | Mann–Whitney U | $6 \times 10^{-1}$ |
| Clinical stage | $\chi^2$ | $1 \times 10^{0}$ |
| Global Gleason | $\chi^2$ | $6 \times 10^{-2}$ |
| Primary Gleason | $\chi^2$ | $2 \times 10^{-2}$ |
| Secondary Gleason | $\chi^2$ | $4 \times 10^{-3}$ |

**Supplementary Table 10** Statistical analysis comparing the distribution of class labels and event indicators between the *learning set* and the *holdout set* using the $\chi^2$ test [2] and the log-rank test [9, 10] for classification and survival tasks, respectively.

| Task | Test | $p$-value |
|---|---|---|
| LNI | $\chi^2$ | $1 \times 10^{0}$ |
| BCR-FS | Log-rank | $7 \times 10^{-1}$ |
| MFS | Log-rank | $7 \times 10^{-1}$ |
| dADT-FS | Log-rank | $2 \times 10^{-1}$ |
| CRPC-FS | Log-rank | $4 \times 10^{-1}$ |
| PCSS | Log-rank | $4 \times 10^{-1}$ |

#### 2 Model hyperparameters

##### 2.1 Hyperparameter search space

**Supplementary Table 11** Fixed hyperparameters common to all models.

| Hyperparameter | Value |
|---|---|
| Activation | PReLU |
| Batch size | 16 |
| Epochs | 100 |
| Learning rate scheduler | Exponential |
| Normalization | Instance |
| Optimizer | Adam |
| Patience | 20 |
| Regularization | L2 |

**Supplementary Table 12** Search space for hyperparameters of both standalone MLPs and those integrated within the SN.

| Hyperparameter | Type | Search space |
|---|---|---|
| Dropout | Float | $[0.05, 0.25]$ |
| Learning rate | Float[1] | $[0.0001, 0.01]$ |
| Layers | Integer | $\{0, 1, 2, 3\}$ |
| Neurons | Integer | $\{5, 10, 15, 20\}$ |
| Weight decay | Float[1] | $[0.0001, 0.01]$ |
| $\alpha$ | Float[1] | $[0.00001, 0.01]$ |
| $\gamma$ | Fixed | 0.99 |

[1]The value is sampled from the range in the log domain.

**Supplementary Table 13** Search space for hyperparameters of the Bayesian MLPs integrated in the BSN.

| Hyperparameter | Type | Search space |
|---|---|---|
| Dropout | Float | $[0, 0.25]$ |
| Learning rate | Float[1] | $[0.0001, 0.01]$ |
| Layers | Integer | $\{0, 1, 2, 3\}$ |
| Neurons | Integer | $\{5, 10, 15, 20\}$ |
| Weight decay | Float[1] | $[0.0001, 0.01]$ |
| $\alpha$ | Float[1] | $[0.00001, 0.01]$ |
| $\gamma$ | Fixed | 0.99 |
| Temperature | Float[1] | $[0.0001, 0.1]$ |

[1]The value is sampled from the range in the log domain.

**Supplementary Table 14** Search space for hyperparameters of the U-Net.

| Hyperparameter | Type | Search space |
|---|---|---|
| Channels | Fixed | (64, 128, 256, 512, 1024) |
| Dropout | Float | $[0, 0.3]$ |
| Kernel size | Fixed | 3 |
| Learning rate | Float[1] | [0.0005, 0.005] |
| Residual units | Fixed | 3 |
| Weight decay | Fixed | 0.01 |
| $\alpha$ | Fixed | 0 |
| $\gamma$ | Fixed | 0.99 |

[1]The value is sampled from the range in the log domain.

**Supplementary Table 15** Search space for hyperparameters of the Bayesian U-Net.

| Hyperparameter | Type | Search space |
|---|---|---|
| Channels | Fixed | (64, 128, 256, 512, 1024) |
| Dropout | Float | $[0, 0.1]$ |
| Kernel size | Fixed | 3 |
| Learning rate | Float[1] | [0.0005, 0.005] |
| Residual units | Fixed | 3 |
| Temperature | Fixed | 0.0001 |
| Weight decay | Fixed | 0.01 |
| $\alpha$ | Fixed | 0 |
| $\gamma$ | Fixed | 0.99 |

[1]The value is sampled from the range in the log domain.

**Supplementary Table 16** Search space for hyperparameters of the U-NEXtractor.

| Hyperparameter | Type | Search space |
|---|---|---|
| Channels | Fixed | (64, 128, 256, 512, 1024) |
| Dropout (CNN) | Float | $[0.2, 0.8]$ |
| Dropout (FNN) | Float | $[0.1, 0.4]$ |
| Kernel size | Fixed | 3 |
| Learning rate | Float[1] | $[0.0001, 0.001]$ |
| Loss weights ($\omega_{\text{prognosis}}$, $\omega_{\text{segmentation}}$) | Categorical | $\{(0.25, 0.75), (0.33, 0.67), (0.5, 0.5), (0.67, 0.33), (0.75, 0.25)\}$ |
| Residual units | Fixed | 2 |
| Weight decay | Float[1] | $[0.001, 0.1]$ |
| $\alpha$ | Float[1] | $[0.0001, 0.01]$ |
| $\gamma$ | Fixed | 0.95 |

[1]The value is sampled from the range in the log domain.

**Supplementary Table 17** Search space for hyperparameters of the Bayesian U-NEXtractor.

| Hyperparameter | Type | Search space |
|---|---|---|
| Channels | Fixed | (64, 128, 256, 512, 1024) |
| Dropout (CNN) | Float | $[0, 0.6]$ |
| Dropout (FNN) | Float | $[0, 0.3]$ |
| Kernel size | Fixed | 3 |
| Learning rate | Float[1] | $[0.0001, 0.001]$ |
| Loss weights ($\omega_{\text{prognosis}}$, $\omega_{\text{segmentation}}$) | Categorical | $\{(0.25, 0.75), (0.33, 0.67), (0.5, 0.5), (0.67, 0.33), (0.75, 0.25)\}$ |
| Residual units | Fixed | 2 |
| Temperature (Prognosis task) | Float[1] | $[0.0001, 0.1]$ |
| Temperature (Segmentation task) | Fixed | 0.0001 |
| Weight decay | Float[1] | $[0.001, 0.1]$ |
| $\alpha$ | Float[1] | $[0.0001, 0.01]$ |
| $\gamma$ | Fixed | 0.95 |

[1]The value is sampled from the range in the log domain.

#### 2.2 Selected hyperparameter values

**Supplementary Table 18** Hyperparameters of the Bayesian MLPs integrated in the BSN trained with the *learning set*.

| Hyperparameter | LNI | BCR-FS | MFS | dADT-FS | CRPC-FS | PCSS |
|---|---|---|---|---|---|---|
| Dropout | 0.10 | 0.10 | 0.05 | 0.15 | 0.05 | 0.05 |
| Learning rate | 0.002 | 0.002 | 0.001 | 0.002 | 0.002 | 0.002 |
| Layers | 2 | 1 | 0 | 2 | 1 | 2 |
| Neurons | 10 | 15 | 0 | 10 | 10 | 10 |
| Weight decay | 0.001 | 0.001 | 0.001 | 0.001 | 0.005 | 0.001 |
| $\alpha$ | 0.001 | 0.001 | 0.001 | 0.005 | 0.0005 | 0.001 |
| $\gamma$ | 0.99 | 0.99 | 0.99 | 0.99 | 0.99 | 0.99 |
| Temperature | 0.0001 | 0.001 | 0.0001 | 0.0001 | 0.001 | 0.001 |

**Supplementary Table 19** Hyperparameters of the Bayesian U-NEXtractor integrated in the BSN trained with the *learning set*.

| Hyperparameter | Value |
|---|---|
| Channels | (64, 128, 256, 512, 1024) |
| Dropout (CNN) | 0.4 |
| Dropout (FNN) | 0.1 |
| Kernel size | 3 |
| Learning rate | 0.0001 |
| Loss weights ($\omega_{\text{prognosis}}, \omega_{\text{segmentation}}$) | $(0.5, 0.5)$ |
| Residual units | 2 |
| Temperature (BCR-FS task) | 0.01 |
| Temperature (Segmentation task) | 0.0001 |
| Weight decay | 0.01 |
| $\alpha$ | $[0.0001, 0.01]$ |
| $\gamma$ | 0.95 |

**Supplementary Table 20** Hyperparameters of the Bayesian U-Net integrated in the BSN trained with the *learning set*.

| Hyperparameter | Value |
|---|---|
| Channels | (64, 128, 256, 512, 1024) |
| Dropout | 0 |
| Kernel size | 3 |
| Learning rate | 0.001 |
| Residual units | 3 |
| Temperature | 0.0001 |
| Weight decay | 0.01 |
| $\alpha$ | 0 |
| $\gamma$ | 0.99 |

### 3 Handcrafted radiomics extraction parameters

**Supplementary Table 21** Parameters for the extraction of handcrafted radiomic features on the CT image using the `pyradiomics` [13] Python library. Description of the parameters is available on the web page of the `pyradiomics` package at https://pyradiomics.readthedocs.io/en/latest/features.html.

| Parameter | Value |
|---|---|
| Bin width | 25 |
| Interpolator | B-spline |
| Sigma | $[1, 2, 3, 4, 5]$ |
| Resegment range | $[-500, \infty]$ |
| Image type | Original |
| Feature class | Shape, first-order, glcm, glrlm, glszm, gldm, ngtdm |

**Supplementary Table 22** Parameters for the extraction of handcrafted radiomic features on the PET image using the `pyradiomics` [13] Python library. Description of the parameters is available on the web page of the `pyradiomics` package at https://pyradiomics.readthedocs.io/en/latest/features.html.

| Parameter | Value |
|---|---|
| Bin width | 1 |
| Interpolator | B-spline |
| Sigma | $[1, 2, 3, 4, 5]$ |
| Resegment range | $[0, 25]$ |
| Image type | Original |
| Feature class | Shape, first-order, glrlm, glszm, gldm, ngtdm |

### 4 Experiments results

#### 4.1 Selected handcrafted radiomic features

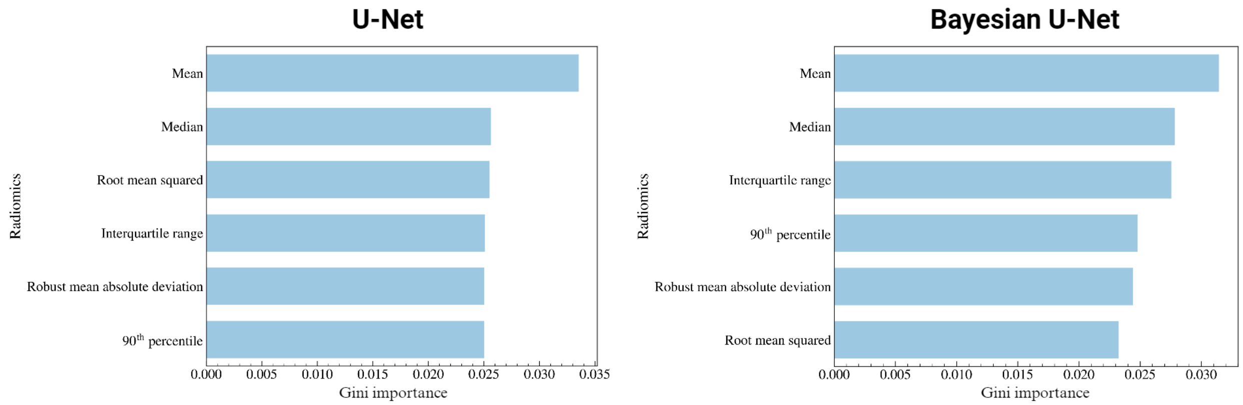


**Supplementary Fig. 19** Gini importance [14] of the 6 most important handcrafted radiomic features extracted from the PET image in the region defined by the prostate segmentation map generated by the U-Net and the Bayesian U-Net. The Gini importance is obtained from a random forest classifier with 10,000 trees implemented with the `scikit-learn` [15] Python library and trained to predict LNI using 200 radiomic features extracted on both CT and PET images. The 6 most important features all come from the PET image. Both the segmentation networks and the random forest classifier are trained using the *learning set*.

#### 4.2 Model performance

**Supplementary Table 23** Performance of the models for the LNI task. See Table 1 for the full caption.

| | | Model | Data | LNI AUC | LNI BA | LNI TPR | LNI TNR |
|---|---|---|---|---|---|---|---|
| Test sets | (A) | MSKCC | CD | 70±7 | 65±5 | 67±17 | 62±21 |
| | | CAPRA | CD | 62±4 | 52±5 | 30±21 | 74±28 |
| | (B) | MLP | CD | 69±7 | 64±6 | 66±8 | 62±14 |
| | | MLP | CD+HCR | **72±5** | **67±3** | 60±11 | 74±6 |
| | | MLP | CD+DLR | 54±7 | 52±5 | 61±25 | 42±27 |
| | (C) | SN | (B)'s best[1] | **72±5** | **67±3** | 60±11 | 74±6 |
| | | BSN | (B)'s best[1] | 71±4 | 67±4 | 58±14 | 76±10 |
| | (D) | $BSN_{t=12}$ | (B)'s best[1] | 71±4 | 67±4 | 58±14 | 76±10 |
| | | $BSN_{t=24}$ | (B)'s best[1] | 71±4 | 67±4 | 58±14 | 76±10 |
| | | $BSN_{t=60}$ | (B)'s best[1] | 71±4 | 67±4 | 58±14 | 76±10 |
| Holdout set | (E) | MSKCC | CD | 60.6 | 52.2 | 23.1 | 81.3 |
| | | CAPRA | CD | 57.7 | 50.0 | 0.0 | 100.0 |
| | (F) | SN | (B)'s best[1] | 65.4 | 65.1 | 61.5 | 68.8 |
| | | BSN | (B)'s best[1] | **66.3** | **66.7** | 61.5 | 71.9 |
| | (G) | $BSN_{t=12}$ | (B)'s best[1] | 66.3 | 66.7 | 61.5 | 71.9 |
| | | $BSN_{t=24}$ | (B)'s best[1] | 66.3 | 66.7 | 61.5 | 71.9 |
| | | $BSN_{t=60}$ | (B)'s best[1] | 66.3 | 66.7 | 61.5 | 71.9 |

[1] Section (B)'s best data is CD+HCR for LNI, CD+DLR for BCR-FS and CD for MFS, dADT-FS, CRPC-FS and PCSS.

**Supplementary Table 24** Results of intermediate experiments on the *test sets* showing pairwise comparison of the performance of different models. The prostate segmentation maps generated by the U-Net, which are used to compute automatic handcrafted radiomic features (AHCR), achieved an average Dice similarity coefficient (DSC) [16] of 0.842± 0.004 compared to manual contours, which are used to compute the manual handcrafted radiomic features (MHCR).

| | | | Task | | | | | | | | | | | | | | | | | | |
|---|---|---|---|---|---|---|---|---|---|---|---|---|---|---|---|---|---|---|---|---|---|
| | | | LNI | | | | BCR-FS | | | MFS | | | dADT-FS | | | CRPC-FS | | | PCSS | | |
| | Model | Data | AUC | BA | TPR | TNR | CI | CICW | CDA | CI | CICW | CDA | CI | CICW | CDA | CI | CICW | CDA | CI | CICW | CDA |
| A | LR | CD | 68±7 | 64±6 | 67±8 | 62±14 | 62±7 | 63±5 | 68±8 | 66±4 | 63±9 | 70±9 | 68±5 | 67±7 | 70±8 | 65±13 | 56±14 | 72±13 | 64±11 | 59±21 | 61±13 |
| A | MLP | CD | 69±7 | 64±6 | 66±8 | 62±14 | 63±7 | 63±5 | 66±6 | 67±6 | 64±12 | 74±8 | 68±6 | 67±5 | 68±8 | 71±10 | 69±7 | 72±9 | 70±16 | 73±23 | 58±34 |
| B | MLP | MHCR | 61±6 | 57±2 | 53±11 | 62±11 | 50±5 | 52±5 | 48±10 | 40±14 | 44±15 | 40±19 | 55±9 | 54±11 | 53±11 | 56±13 | 57±15 | 55±14 | 55±25 | 63±27 | 51±26 |
| B | MLP | AHCR | 58±10 | 57±7 | 31±22 | 82±10 | 48±5 | 49±4 | 45±2 | 47±9 | 43±10 | 51±14 | 54±5 | 53±5 | 56±6 | 61±18 | 60±18 | 62±19 | 49±28 | 50±30 | 51±30 |
| C | MLP | CD+MHCR | 73±4 | 67±4 | 63±11 | 72±11 | 59±9 | 59±8 | 62±9 | 57±7 | 54±6 | 58±9 | 61±10 | 61±8 | 62±7 | 68±9 | 70±9 | 67±8 | 68±9 | 70±9 | 67±8 |
| C | MLP | CD+AHCR | 72±5 | 67±3 | 60±11 | 74±6 | 58±7 | 58±4 | 58±9 | 58±4 | 53±9 | 62±6 | 64±12 | 62±10 | 66±11 | 67±10 | 65±10 | 69±17 | 70±10 | 64±20 | 74±15 |
| D | CNN | CT+PET | 55±5 | 52±2 | 50±43 | 54±45 | 53±4 | 52±5 | 58±4 | 53±9 | 54±9 | 53±16 | 54±8 | 54±9 | 54±10 | 45±20 | 50±17 | 42±23 | 50±26 | 45±24 | 52±33 |
| D | U-NEX | CT+PET | 50±3 | 50±2 | 31±37 | 69±39 | 55±3 | 56±3 | 60±11 | 61±12 | 63±13 | 61±17 | 50±4 | 53±45 | 54±9 | 49±10 | 54±14 | 46±11 | 32±13 | 44±20 | 30±20 |

**Supplementary Table 25** Dice similarity coefficient (DSC) [16] on the *training set*, the *validation set*, and the *holdout set* of segmentation models trained with the *learning set*.

| | Training set | | | Validation set | | | Holdout set | | |
|---|---|---|---|---|---|---|---|---|---|
| Model | Mean (Median) | Min-Max | Std | Mean (Median) | Min-Max | Std | Mean (Median) | Min-Max | Std |
| U-Net | 0.947 (0.954) | 0.778 - 0.972 | 0.025 | 0.843 (0.849) | 0.578 - 0.925 | 0.064 | 0.845 (0.869) | 0.298 - 0.931 | 0.099 |
| Bayesian U-Net | 0.935 (0.941) | 0.802 - 0.965 | 0.024 | 0.838 (0.844) | 0.612 - 0.931 | 0.059 | 0.834 (0.866) | 0.246 - 0.917 | 0.109 |
| U-NEXtractor | 0.249 (0.237) | 0.095 - 0.608 | 0.086 | 0.259 (0.251) | 0.117 - 0.499 | 0.088 | 0.259 (0.255) | 0.092 - 0.460 | 0.080 |
| Bayesian U-NEXtractor | 0.044 (0.039) | 0.016 - 0.169 | 0.020 | 0.046 (0.042) | 0.019 - 0.106 | 0.019 | 0.046 (0.044) | 0.014 - 0.114 | 0.018 |

#### 4.3 Statistical analysis comparing model performance

**Supplementary Table 26** Statistical analysis comparing the performance of the BSN against the MSKCC nomogram, the CAPRA score and the SN on the *test sets* and the *holdout set*. AUC $p$-values are determined using the fast implementation of DeLong test [17, 18], while CI $p$-values are calculated with the U-statistics-based C estimator [19]. All other $p$-values are obtained by bootstrap [20, 21] with 10 000 repetitions. The $p$-value shown on *test sets* is the median of the $p$-values calculated on the 5 *test sets*. Color code: increase (cyan), decrease (red) and no significant change (black) of the performance of the BSN compared with the reference model.

| | | | Task | | | | | | | | | | | | | | | | |
|---|---|---|---|---|---|---|---|---|---|---|---|---|---|---|---|---|---|---|---|
| | | | LNI | | BCR-FS | | | MFS | | | dADT-FS | | | CRPC-FS | | | PCSS | | |
| | Model | Data | AUC | BA | CI | CICW | CDA | CI | CICW | CDA | CI | CICW | CDA | CI | CICW | CDA | CI | CICW | CDA |
| Test sets | MSKCC | CD | $1\times10^{-1}$ | $3\times10^{-1}$ | $8\times10^{-1}$ | $5\times10^{-1}$ | $1\times10^{-1}$ | — | — | — | — | — | — | — | — | — | $5\times10^{-3}$ | $3\times10^{-2}$ | $3\times10^{-2}$ |
| Test sets | CAPRA | CD | $4\times10^{-2}$ | $2\times10^{-1}$ | $3\times10^{-1}$ | $5\times10^{-1}$ | $8\times10^{-2}$ | $5\times10^{-2}$ | $8\times10^{-2}$ | $6\times10^{-2}$ | $2\times10^{-2}$ | $5\times10^{-2}$ | $2\times10^{-1}$ | $2\times10^{-3}$ | $2\times10^{-2}$ | $1\times10^{-2}$ | $1\times10^{-2}$ | $9\times10^{-2}$ | $3\times10^{-2}$ |
| Test sets | SN | Best | $1\times10^{-1}$ | $3\times10^{-1}$ | $1\times10^{-1}$ | $2\times10^{-1}$ | $3\times10^{-2}$ | $3\times10^{-1}$ | $4\times10^{-1}$ | $3\times10^{-1}$ | $2\times10^{-1}$ | $6\times10^{-1}$ | $3\times10^{-1}$ | $2\times10^{-1}$ | $5\times10^{-1}$ | $3\times10^{-1}$ | $1\times10^{-1}$ | $3\times10^{-1}$ | $3\times10^{-1}$ |
| Holdout set | MSKCC | CD | $5\times10^{-1}$ | $1\times10^{-1}$ | $3\times10^{-1}$ | $6\times10^{-1}$ | $1\times10^{0}$ | — | — | — | — | — | — | — | — | — | $3\times10^{-1}$ | $6\times10^{-1}$ | $3\times10^{-2}$ |
| Holdout set | CAPRA | CD | $4\times10^{-1}$ | $1\times10^{-1}$ | $3\times10^{-1}$ | $2\times10^{-1}$ | $3\times10^{-1}$ | $1\times10^{-1}$ | $4\times10^{-1}$ | $2\times10^{-2}$ | $7\times10^{-1}$ | $6\times10^{-1}$ | $7\times10^{-1}$ | $9\times10^{-1}$ | $6\times10^{-1}$ | $3\times10^{-1}$ | $6\times10^{-3}$ | $2\times10^{-1}$ | $5\times10^{-1}$ |
| Holdout set | SN | Best | $6\times10^{-1}$ | $1\times10^{0}$ | $6\times10^{-1}$ | $9\times10^{-1}$ | $6\times10^{-1}$ | $2\times10^{-1}$ | $3\times10^{-1}$ | $4\times10^{-1}$ | $8\times10^{-1}$ | $9\times10^{-1}$ | $6\times10^{-1}$ | $3\times10^{-1}$ | $3\times10^{-1}$ | $4\times10^{-1}$ | $1\times10^{0}$ | $1\times10^{0}$ | $1\times10^{0}$ |

**Supplementary Table 27** Statistical analysis comparing the performance of the MLP with the best data (CD+HCR for LNI, CD+DR for BCR-FS and CD for MFS, dADT-FS, CRPC-FS and PCSS) against the MLP with CD alone, and the SN with the best data. The models are evaluated on the *test sets*. AUC $p$-values are determined using the fast implementation of DeLong test [17, 18], while CI $p$-values are calculated with the U-statistics-based C estimator [19]. All other $p$-values are obtained by bootstrap [20, 21] with 10 000 repetitions. The $p$-value shown is the median of the $p$-values calculated on the 5 *test sets*. Color code: increase (cyan), decrease (red), and no significant changes (black) of the performance of the MLP with the best data compared with the reference model.

| | | Task | | | | | | | | | | | | | | | | |
|---|---|---|---|---|---|---|---|---|---|---|---|---|---|---|---|---|---|---|
| Model | Data | LNI | | BCR-FS | | | MFS | | | dADT-FS | | | CRPC-FS | | | PCSS | | |
| | | AUC | BA | CI | CICW | CDA | CI | CICW | CDA | CI | CICW | CDA | CI | CICW | CDA | CI | CICW | CDA |
| MLP | CD | $5\times10^{-1}$ | $2\times10^{-1}$ | $3\times10^{-1}$ | $5\times10^{-1}$ | $2\times10^{-1}$ | — | — | — | — | — | — | — | — | — | — | — | — |
| SN | Best | $1\times10^{0}$ | $1\times10^{0}$ | $8\times10^{-1}$ | $5\times10^{-1}$ | $8\times10^{-1}$ | $3\times10^{-1}$ | $5\times10^{-1}$ | $5\times10^{-1}$ | $3\times10^{-1}$ | $5\times10^{-1}$ | $1\times10^{-1}$ | $5\times10^{-1}$ | $7\times10^{-1}$ | $2\times10^{-1}$ | $4\times10^{-1}$ | $6\times10^{-1}$ | $3\times10^{-1}$ |

### 5 Illustration of clinical application

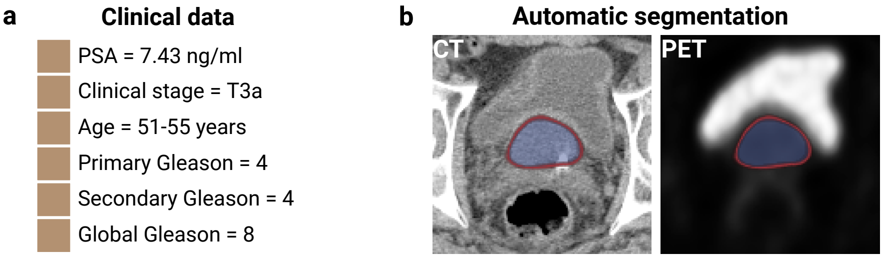


**Model prediction**

c

| Task | Positive class probability | Survival probability at 2 years | Survival probability at 5 years | Risk group |
|---|---|---|---|---|
| LNI | $0.73 \pm 0.07$ | — | — | High risk |
| BCR-FS | — | $0.48 \pm 0.04$ | $0.31 \pm 0.04$ | High risk |
| MFS | — | $0.86 \pm 0.02$ | $0.70 \pm 0.04$ | High risk |
| dADT-FS | — | $0.81 \pm 0.02$ | $0.72 \pm 0.03$ | High risk |
| CRPC-FS | — | $0.95 \pm 0.01$ | $0.87 \pm 0.03$ | High risk |
| PCSS | — | $0.998 \pm 0.001$ | $0.990 \pm 0.005$ | Low risk |

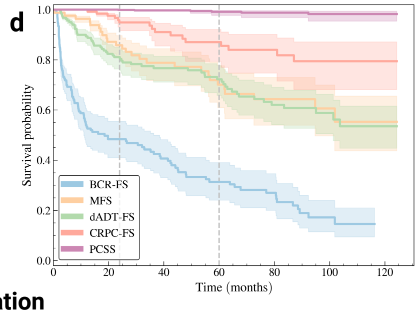


**Model interpretation**

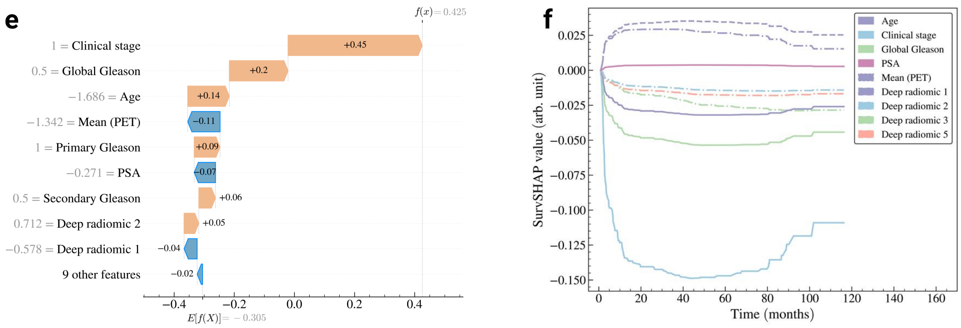


**Ground truth**

g

| Task | Target | Event time or censoring time (months) |
|---|---|---|
| LNI | 1 | — |
| BCR-FS | 1 | 2.4 |
| MFS | 1 | 17.4 |
| dADT-FS | 1 | 4.6 |
| CRPC-FS | 1 | 11.3 |
| PCSS | 1 | 46.9 |

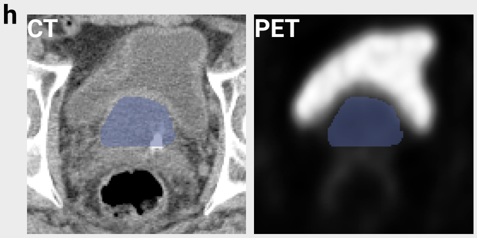


**Supplementary Fig. 20** Prognosis of an arbitrarily selected patient from the *holdout set*. (a) Clinical features of the patient. (b) Segmentation map of the prostate obtained from the Bayesian U-Net trained on the *learning set*. The segmentation map is overlaid on the CT and PET images to illustrate that the region of high FDG uptake by the bladder lies outside the boundaries of the segmentation map. The segmentation map is used to extract handcrafted radiomic features. The DSC between the automatic and the manual segmentation (ground truth) is 0.872. Color code: average prostate segmentation map (blue) and standard deviation (red) over 100 inferences. See Supplementary Fig. Xb for the segmentation map obtained by the Bayesian U-NEXtractor (c) Average prediction and standard deviation of the model over 100 inferences. (d) Average survival curves predicted by the model (line) and 95% confidence interval (shade) over 100 inferences. (e) Shapley additive explanation (SHAP) [22] of the predicted risk of BCR-FS. (f) Time-dependent SHAP (SurvSHAP($t$)) [23] of the predicted risk of BCR-FS. (g) Ground truth progression of the patient's cancer. Time represents the survival time when the target value is 1 and acts as a censoring time otherwise. (h) Ground truth prostate segmentation map obtained from manual contouring by a physician.

**a** **Clinical data**

- PSA = 5.10 ng/ml
- Clinical stage = T1-T2
- Age = 56-60 years
- Primary Gleason = 4
- Secondary Gleason = 4
- Global Gleason = 8

**b** **Automatic segmentation**

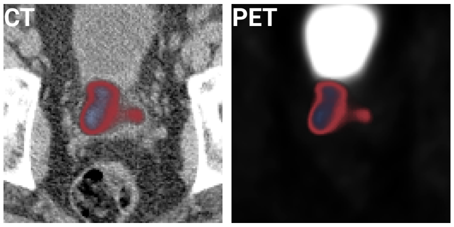


**Model prediction**

**c**

| Task | Positive class probability | Survival probability at 2 years | Survival probability at 5 years | Risk group |
|---|---|---|---|---|
| LNI | $0.31 \pm 0.05$ | — | — | Low risk |
| BCR-FS | — | $0.62 \pm 0.02$ | $0.47 \pm 0.03$ | Low risk |
| MFS | — | $0.924 \pm 0.008$ | $0.83 \pm 0.02$ | Low risk |
| dADT-FS | — | $0.85 \pm 0.01$ | $0.77 \pm 0.02$ | Low risk |
| CRPC-FS | — | $0.976 \pm 0.005$ | $0.94 \pm 0.01$ | Low risk |
| PCSS | — | $1.000 \pm 0.001$ | $0.999 \pm 0.001$ | Low risk |

**d**

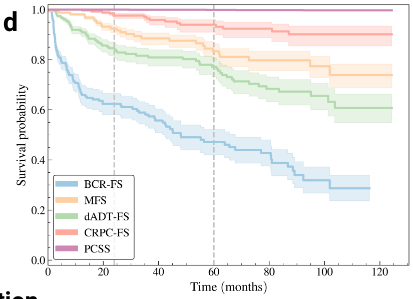


**Model interpretation**

**e**

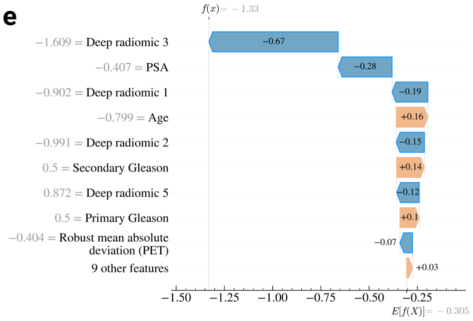


**f**

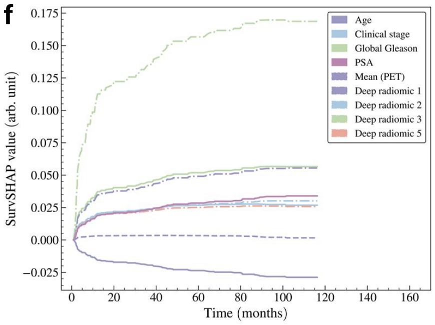


**Ground truth**

**g**

| Task | Target | Event time or censoring time (months) |
|---|---|---|
| LNI | 0 | — |
| BCR-FS | 0 | 100.4 |
| MFS | 0 | 100.4 |
| dADT-FS | 0 | 100.4 |
| CRPC-FS | 0 | 100.4 |
| PCSS | 0 | 100.4 |

**h**

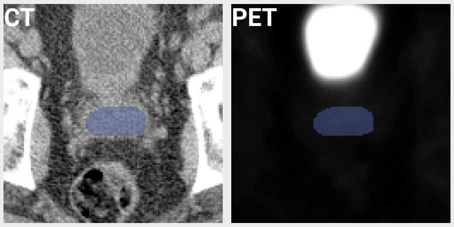


**Supplementary Fig. 21** Prognosis of an arbitrarily selected patient from the *holdout set*. (a) Clinical features of the patient. (b) Segmentation map of the prostate obtained from the Bayesian U-Net trained on the *learning set*. The segmentation map is overlaid on the CT and PET images to illustrate that the region of high FDG uptake by the bladder lies outside the boundaries of the segmentation map. The segmentation map is used to extract handcrafted radiomic features. The DSC between the automatic and the manual segmentation (ground truth) is 0.893. Color code: average prostate segmentation map (blue) and standard deviation (red) over 100 inferences. See Supplementary Fig. Xc for the segmentation map obtained by the Bayesian U-NEXtractor.(c) Average prediction and standard deviation of the model over 100 inferences. (d) Average survival curves predicted by the model (line) and 95% confidence interval (shade) over 100 inferences. (e) Shapley additive explanation (SHAP) [22] of the predicted risk of BCR-FS. (f) Time-dependent SHAP (SurvSHAP($t$)) [23] of the predicted risk of BCR-FS. (g) Ground truth progression of the patient's cancer. Time represents the survival time when the target value is 1 and acts as a censoring time otherwise. (h) Ground truth prostate segmentation map obtained from manual contouring by a physician.

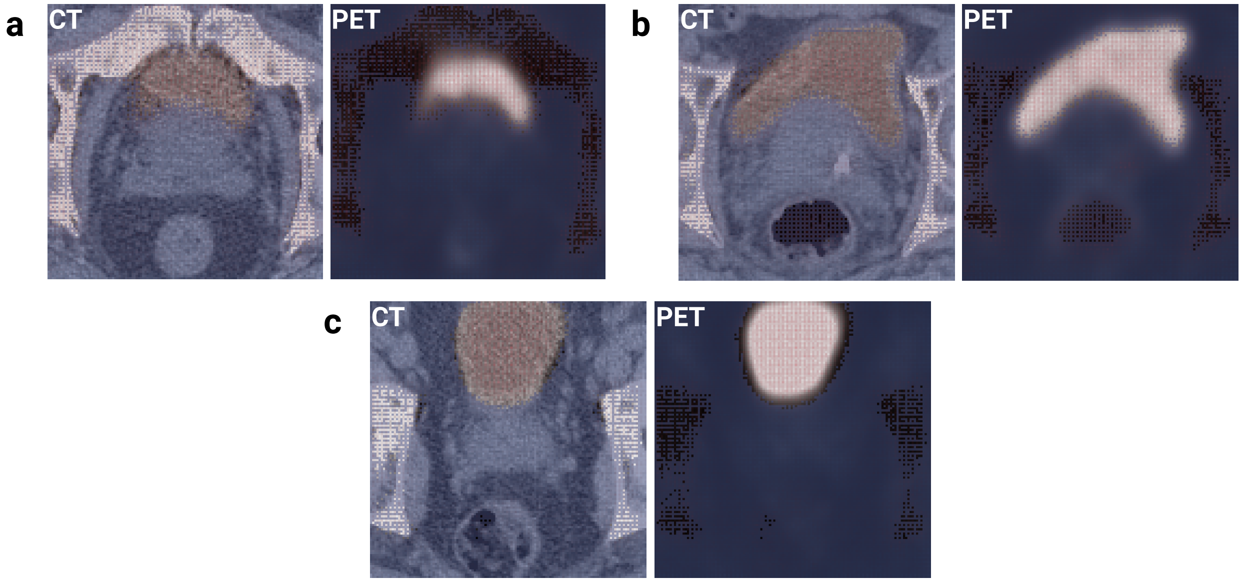


**Supplementary Fig. 22** Average prostate segmentation map (blue) and standard deviation (red) over 100 inferences obtained from the Bayesian U-NEXtractor for the patient shown in (a) Fig. 1, (b) Supplementary Fig. 20, and (c) Supplementary Fig. 21. The segmentation map overlaid on the PET image reveals that the Bayesian U-NEXtractor avoids bones and the region of high FDG uptake by the bladder, and segments everything else.
